# Clinical Pharmaceutical Services for People Living with HIV: A Scoping Review

**DOI:** 10.1101/2025.10.24.25338755

**Authors:** Marlessa Danielle Assis Vidal Rosa, Patricia Danielle Oliveira de Almeida, Ivana Andrade Vieira Neves, Zeni Silva Jucá Bessa, Rebeka Caribe Badin, Ana Cyra dos Santos Lucas

## Abstract

**Background:** Pharmacist-led clinical services play a crucial role in the care of people living with HIV (PLWH), contributing to adherence, therapeutic optimization, and improved health outcomes. This scoping review aimed to map and describe pharmacist-led services for PLWH, including service types, locations, outcomes, and geographic distribution.

**Methods:** A comprehensive literature search was conducted in Medline/PubMed, Scopus, Web of Science, Lilacs, ProQuest, and LA Referencia, covering studies from 1986 onward, with no language restrictions. Eligible studies addressed PLWH on antiretroviral therapy receiving pharmacist-led services in outpatient, hospital, or home-based settings. The review followed the Joanna Briggs Institute (JBI) methodology for scoping reviews.

**Results:** Of the 8,864 records identified, 146 studies met the inclusion criteria. Most were conducted in North America (51.9%) and Europe (21.8%). Study designs included observational (48.9%), interventional (42.8%), and review articles (9.6%). Research output increased over the past two decades, with publication peaks in 2009, 2014, and 2018. Outpatient clinics were the most frequent setting (n=98), followed by hospitals (n=41) and home care (n=2). Most studies occurred in urban areas (n=129). Common services included medication therapy review (68.0%), health education (61.9%), and pharmacotherapeutic follow-up (61.2%). Less frequent services included medication reconciliation, chronic disease management, dispensing, screening, and therapeutic drug monitoring. Services were primarily delivered by pharmacists alone, with some collaboration with physicians or nurses.

**Conclusion:** This review highlights the growing interest in pharmacist-led services for PLWH and offers a foundation for future studies by identifying established areas and research gaps.

## INTRODUCTION

Acquired Immunodeficiency Syndrome (AIDS), caused by the Human Immunodeficiency Virus (HIV), remains a significant global public health challenge ^1^. Since the first documented cases in the late 1970s, advances in antiretroviral therapy (ART) have transformed HIV into a chronic, manageable condition, enabling the social reintegration of people living with HIV/AIDS (PLWH). However, the success of ART hinges on strict adherence and comprehensive care, underscoring the critical role of pharmaceutical services in optimizing health outcomes for these patients.

Currently, patient-facing pharmaceutical services have become essential, being recommended by international organizations both to improve patients’ clinical outcomes and to promote resource savings and quality of life ^2^, as they have demonstrated efficacy in reducing hospital and emergency readmissions and a potential decrease in the incidence of drug-related adverse events and even an improvement in adherence ^3,4^, and have contributed to the economy through improved patient health management and cost reduction for the health system ^5,6^.

A wide range of clinical pharmacy can be implemented ^7^. Pharmaceutical services aimed at the patient, the family and the community are Clinical Pharmacy (or Pharmaceutical Care), Integrative and Complementary Practices and Aesthetics ^8^.

In this manuscript, we use the term “clinical pharmaceutical services” to denote pharmaceutical care activities delivered directly to patients, their families, and the community, including management of self-limiting conditions, dispensing, medication reconciliation, health screening, health education, therapeutic drug monitoring, pharmacotherapy review, disease-state management, and pharmacotherapeutic follow-up.

These services can be performed in different places of practice, including community pharmacy, hospital bed, hospital pharmacy, urgent and emergency services, primary health care services, outpatient clinic, patient’s home, long-term care institutions, among others ^9^.

The preliminary search for evidence on clinical pharmaceutical services for PLWH revealed that, although several scoping reviews have examined the role of such services in other conditions, including diabetes, tuberculosis, sensory loss, and intellectual disability ^10, 11,12,13^, none has focused on PLWH. We did identify systematic reviews that focused on specific components of pharmaceutical care for HIV treatment, such as the clinical impact of antiretroviral therapy ^14^, economic evaluations ^15^, adherence and viral suppression ^16^, medication-related problem management ^17^, and the effectiveness of teleconsultations ^18^.

All these studies are characterized by their focus on specific outcomes, however, in order to obtain a comprehensive view of the various clinical pharmaceutical services applied in the treatment PLWH, the best approach would be a scoping review, as they have a broad interest and the objective of knowing and mapping the available evidence on a given topic ^19,20^, in this case, the clinical pharmaceutical services directly related to the patient, family and community. Initial searches in Open Science Framework (OSF), PROSPERO, and PubMed found no prior comprehensive review on this topic.

The present review aims to identify current practices, knowledge gaps, and guide future research, ultimately supporting healthcare professionals in optimizing pharmaceutical care for PLWH. To our knowledge, this is the first scoping review to map global clinical pharmacy services specifically for PLWH.

## METHODOLOGY

This scoping review adhered to the Joanna Briggs Institute (JBI) methodology ^21^. The protocol was registered in the *Open Science Framework* (OSF) (https://doi.org/10.17605/OSF.IO/D8BHF). The review was documented using the PRISMA extension for scoping reviews (PRISMA-ScR)^22^.

### Research Question

Guided by the PCC framework (Population, Concept, Context), we defined: **Population**: People living with HIV (PLWH); **Concept**: Clinical pharmaceutical services, including the services management of self-limited health problems, dispensing, medication reconciliation, health screening, health education, therapeutic monitoring of medications, review of pharmacotherapy, management of health condition and pharmacotherapeutic follow-up. **Context**: Outpatient, hospital, and home-based care settings.

### Search Strategy

A systematic search was conducted, with no language restrictions, across Medline/PubMed, Lilacs, Web of Science, and Scopus. In addition, the Pro-Quest and LA Referencia databases were used as a source for unpublished studies or grey literature, and reference lists of included studies were consulted to identify any additional relevant studies. The search spanned studies published from 1986 (post-AZT approval), using tailored Boolean syntaxes combining MeSH/DeCS terms for HIV and pharmaceutical care, and they were adapted according to the particularity of each data source (see Appendix 1 – Supplementary Material). The searches were conducted on January 29, 2024 and updated on October 18, 2024, through the alerts, defined in the searches or by a new direct search in the bases where it was not possible to leave alerts.

### Study Selection

Two independent reviewers screened titles/abstracts and full texts using EndNoteWeb ®, Rayyan® and Zotero®. Discrepancies were resolved via consensus or a third reviewer.

Elegibility criteria: **Study designs**: Experimental studies, descriptive and analytical observational studies, and review studies were included. Texts and opinion papers, congress abstracts, qualitative studies focusing on qualitative data, covering projects such as phenomenology, grounded theory, ethnography, qualitative description, action research, and feminist research, were excluded. **Concept:** Studies focused on clinical pharmaceutical services targeting health outcomes in people living with HIV/AIDS through pharmacist-patient interventions. We excluded studies on the development of new drugs, studies related to access and logistics of drug distribution, and studies limited to epidemiological analyse. **Context:** Studies on clinical pharmaceutical services for outpatient, hospital, and home treatment of people living with HIV were included. We excluded studies where clinical pharmaceutical services were provided without the involvement of pharmacists

To ensure clarity and consistency in applying the eligibility criteria, a pilot test was conducted with six sources independently screened by two reviewers for alignment with the inclusion criteria. Following this calibration, study selection proceeded in two phases. First, two reviewers (MV and AL) independently screened titles and abstracts. Discrepancies were resolved by consensus or consultation with a third reviewer (PA). Then, full texts of potentially eligible studies were retrieved from original sources, journals, or institutional repositories. When unavailable, corresponding authors were contacted; studies remained excluded if no response was received after three attempts. The list of non-retrieved studies is presented in Appendix 2 (Supplementary Material). Full-text screening was conducted by two independent reviewers (MV and IV), with disagreements resolved through discussion or consultation with a third reviewer (AL). Excluded studies were documented with reasons in Appendix 3. The entire selection process was depicted in a PRISMA flow diagram (Appendix 4).

### Data Extraction & Synthesis

Data reliability was ensured by verifying 20% of the studies during the data extraction phase, conducted by two independent reviewers who demonstrated a high level of agreement. This process confirmed the quality of the remaining data extracted by a single reviewer, in accordance with recommendations^23^.Data were extracted using a data extraction guide developed by the review team (Appendix 5), and all information was recorded in Microsoft Excel®. A pilot test involving six sources of evidence was performed to ensure clarity and consistency in the extraction process. The extracted data included: title, author, year of publication, study design, country, continent, funding, study objective, study setting, sample size, year of the study, study duration, age group, participant characteristics, description of the clinical pharmaceutical service, type of service, follow-up, provider of the intervention, outcomes, study limit. Additionally, for each included review study, the research question, outcomes assessed, key findings, and the evaluation of quality or risk of bias were also extracted from the study reports.

To characterize and summarize the results, the diagrammatic and tabular form was used. In alignment with the aim of this study, we provide an overview in conceptual categories covering types of clinical pharmaceutical services, sample characteristics, study setting, study objectives, study design, setting, outcomes, key findings, and gaps identified in the current literature.

## RESULTS

### Selection of sources of evidence

The searches in the databases resulted in 9,125 references as shown in the PRISMA flowchart (Appendix 3). After removing 2,768 records in the EndNoteWeb software, and 1,072 studies in the Rayyan ® software, 5,285 were analyzed through titles and abstracts and 4,876 studies were excluded for not meeting the eligibility criteria. In the second stage of selection process, full text of 409 studies were analyzed, and 272 studies were excluded. Thus, 136 studies were included in the review after applying the eligibility criteria. In addition, 10 relevant studies were identified in the reference lists of the included studies, resulting in a final total of 146 studies included in the review.

### Characteristics of the included studies

The general characteristics of the 132 primary studies included are presented in Table 1, and Table 2 presents the characterization of the review studies included (n = 14).

**Table 1.**
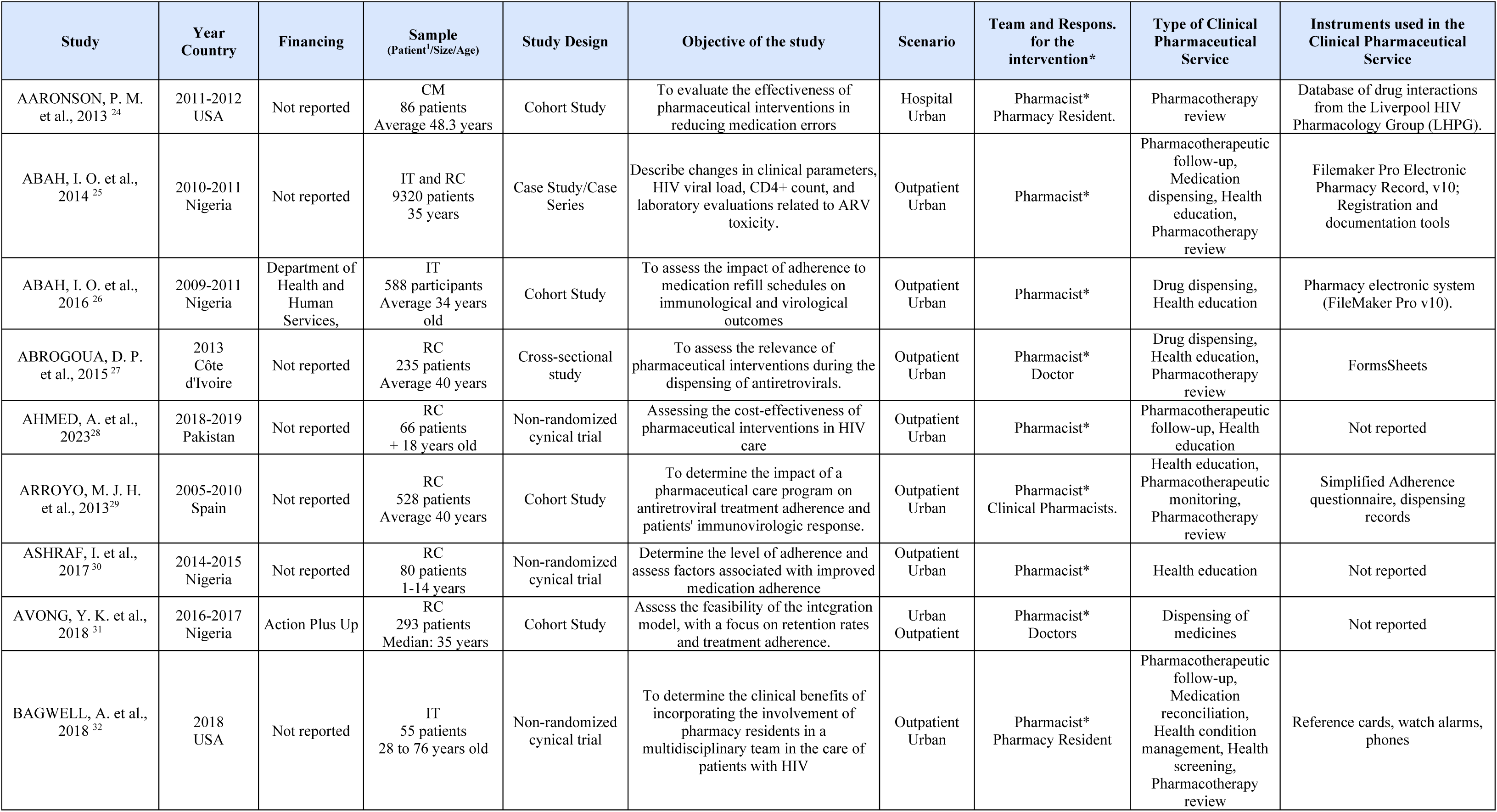
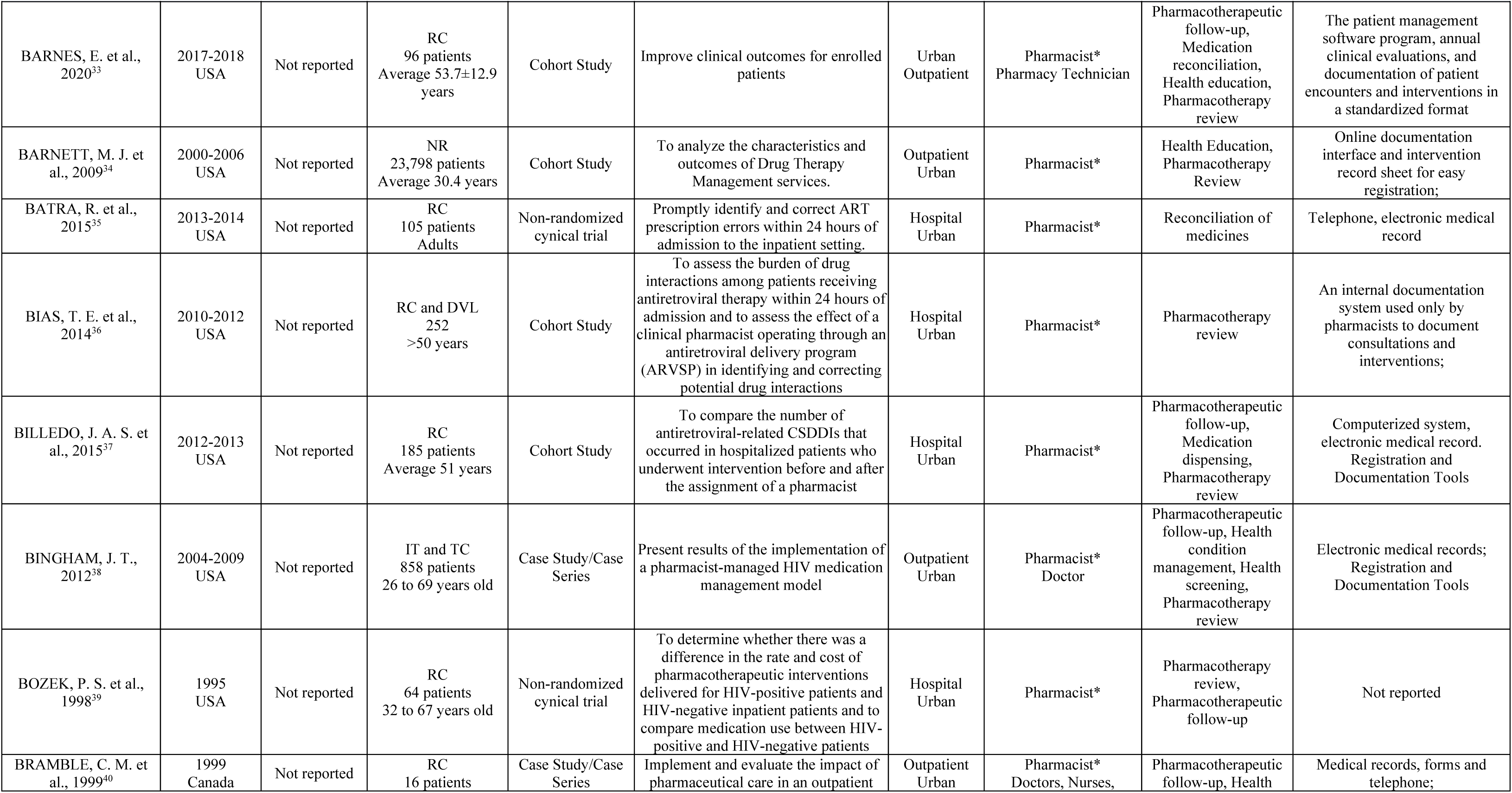
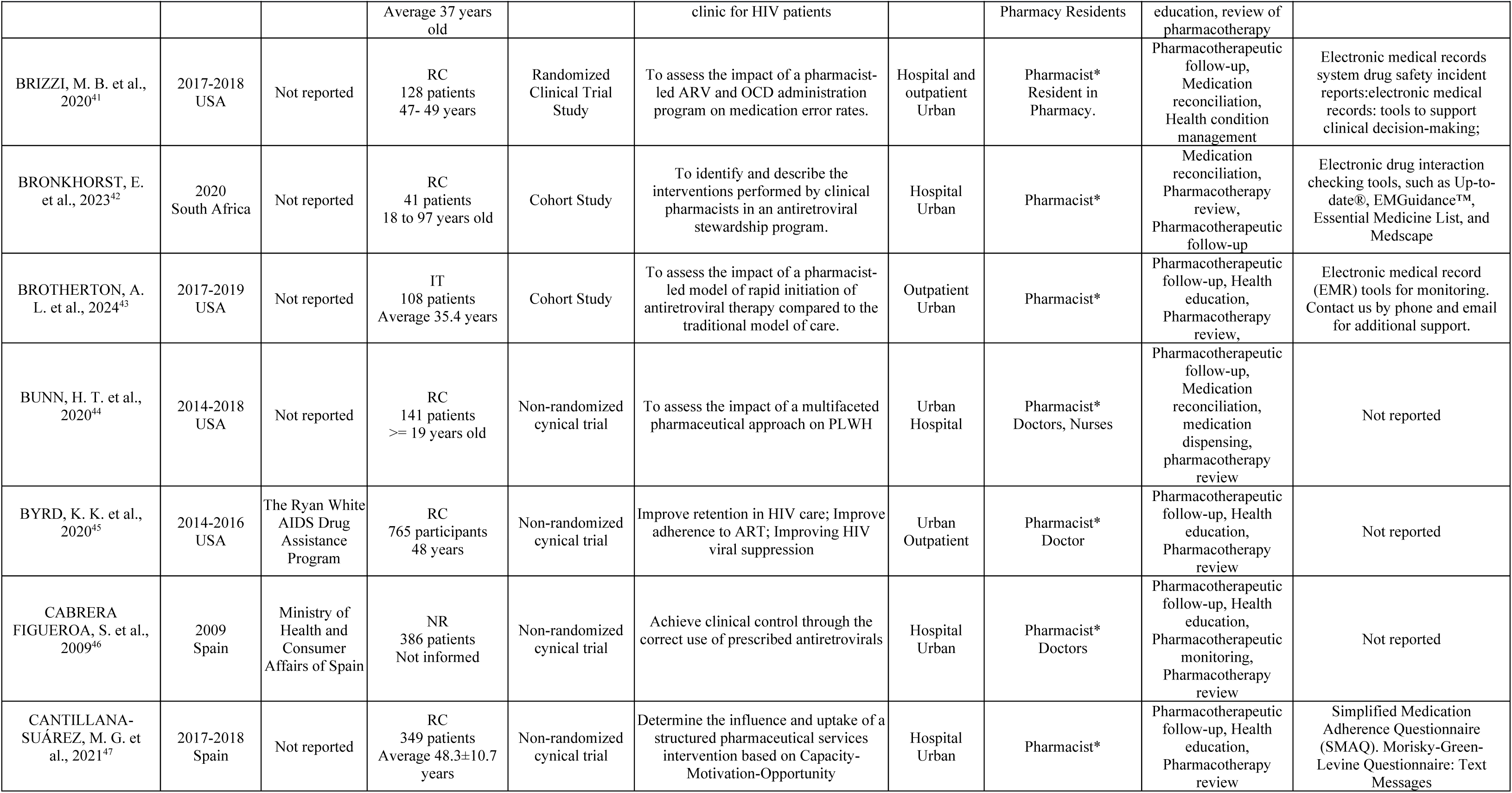
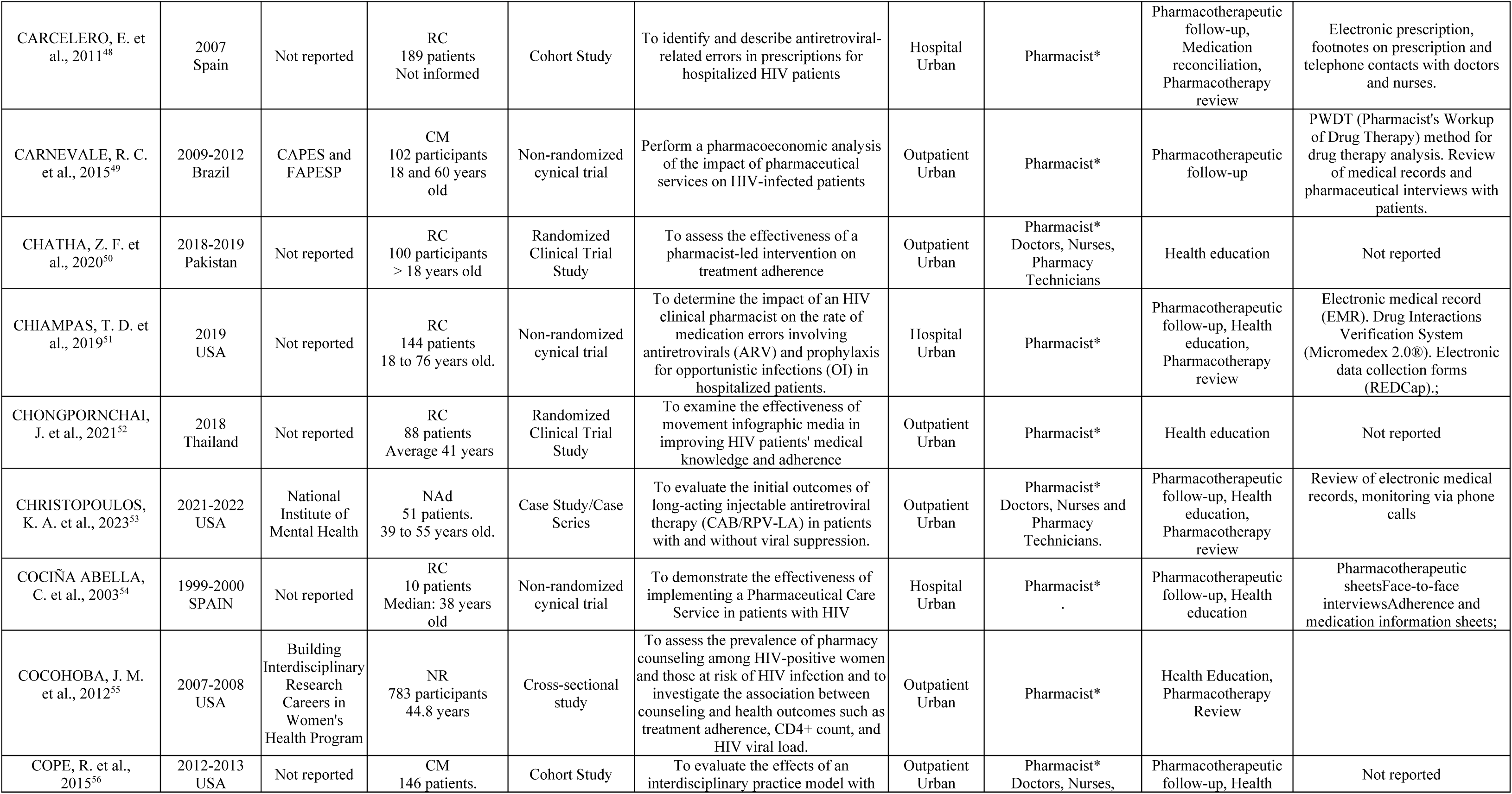
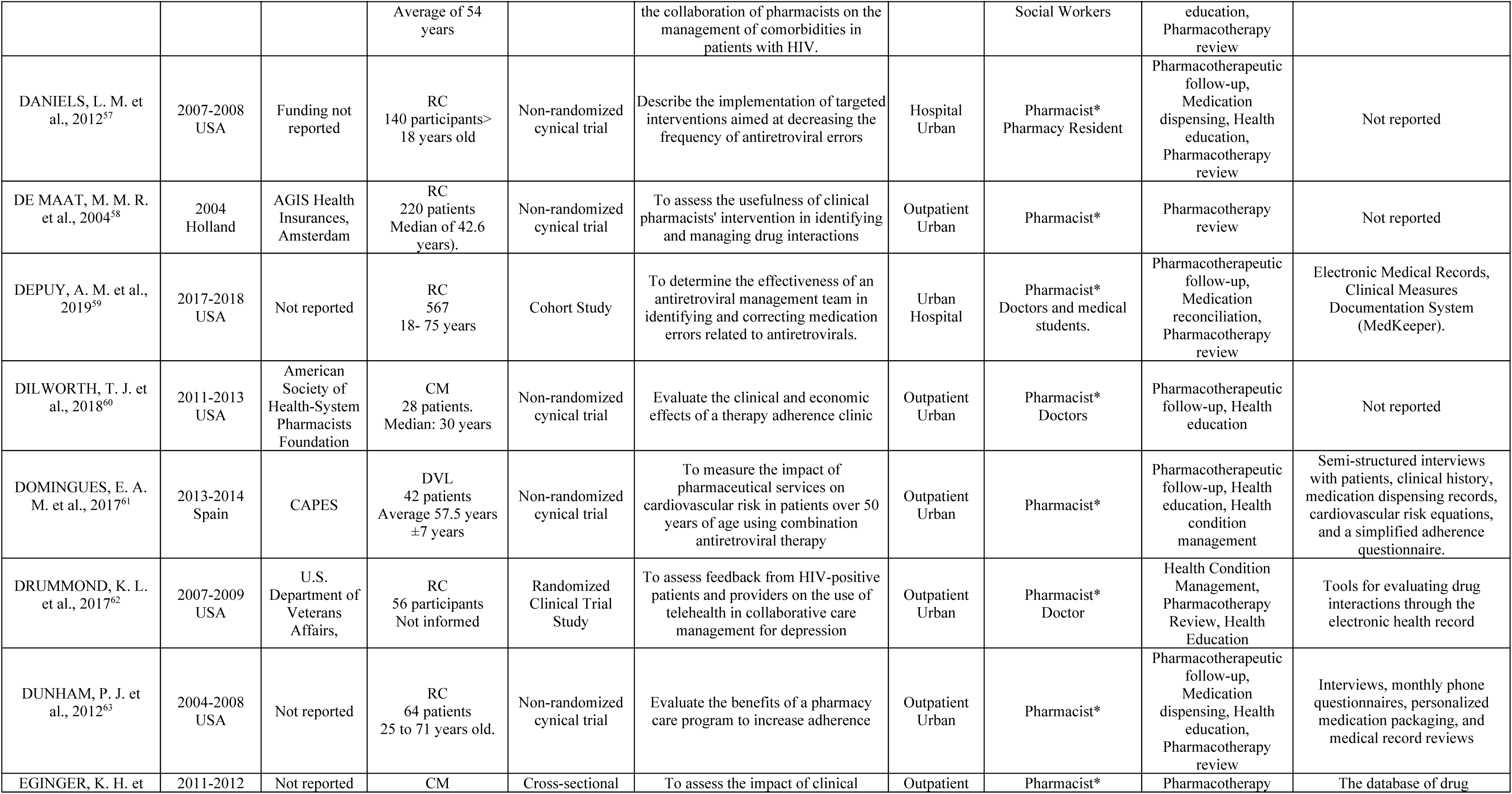
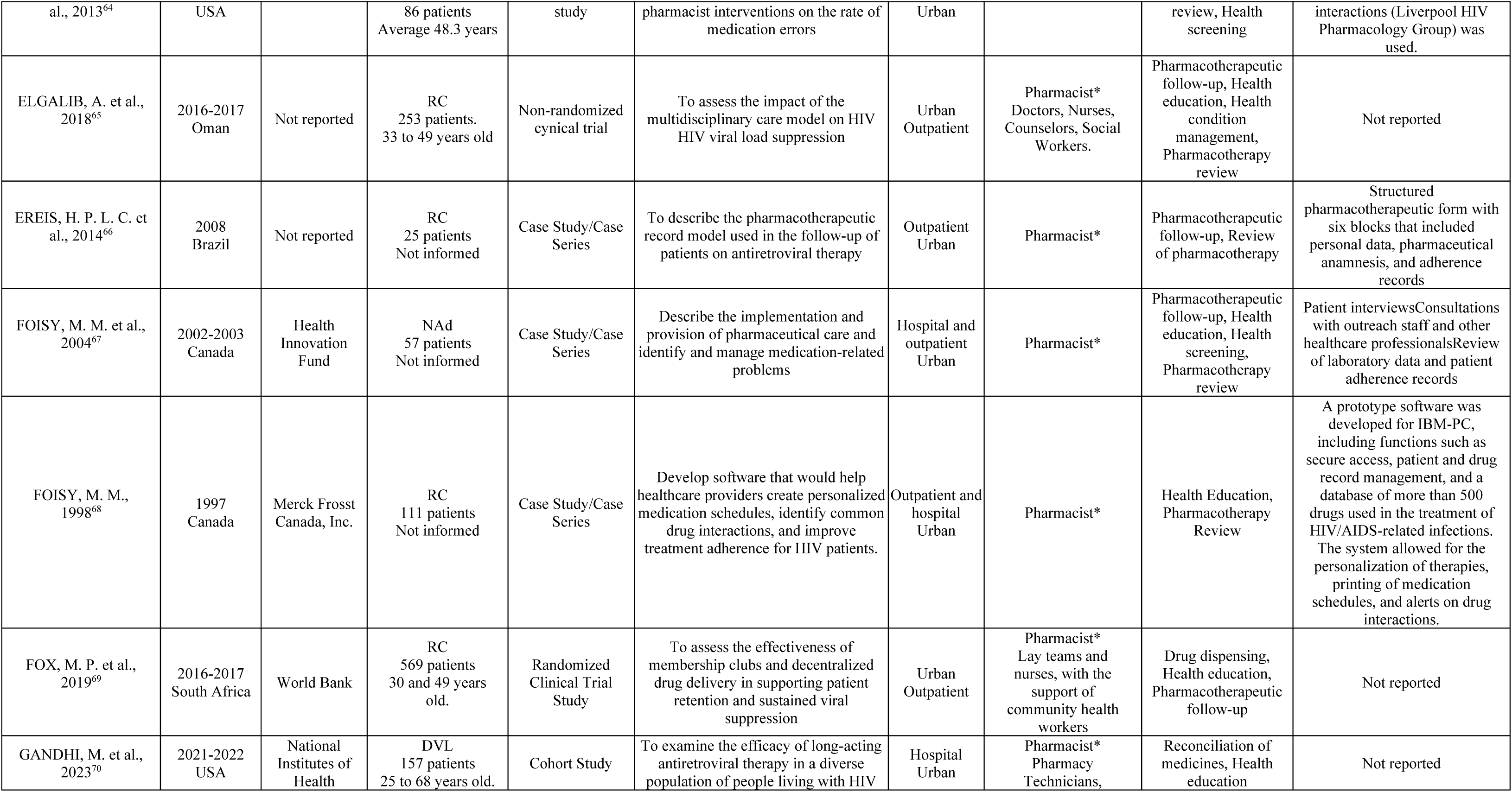
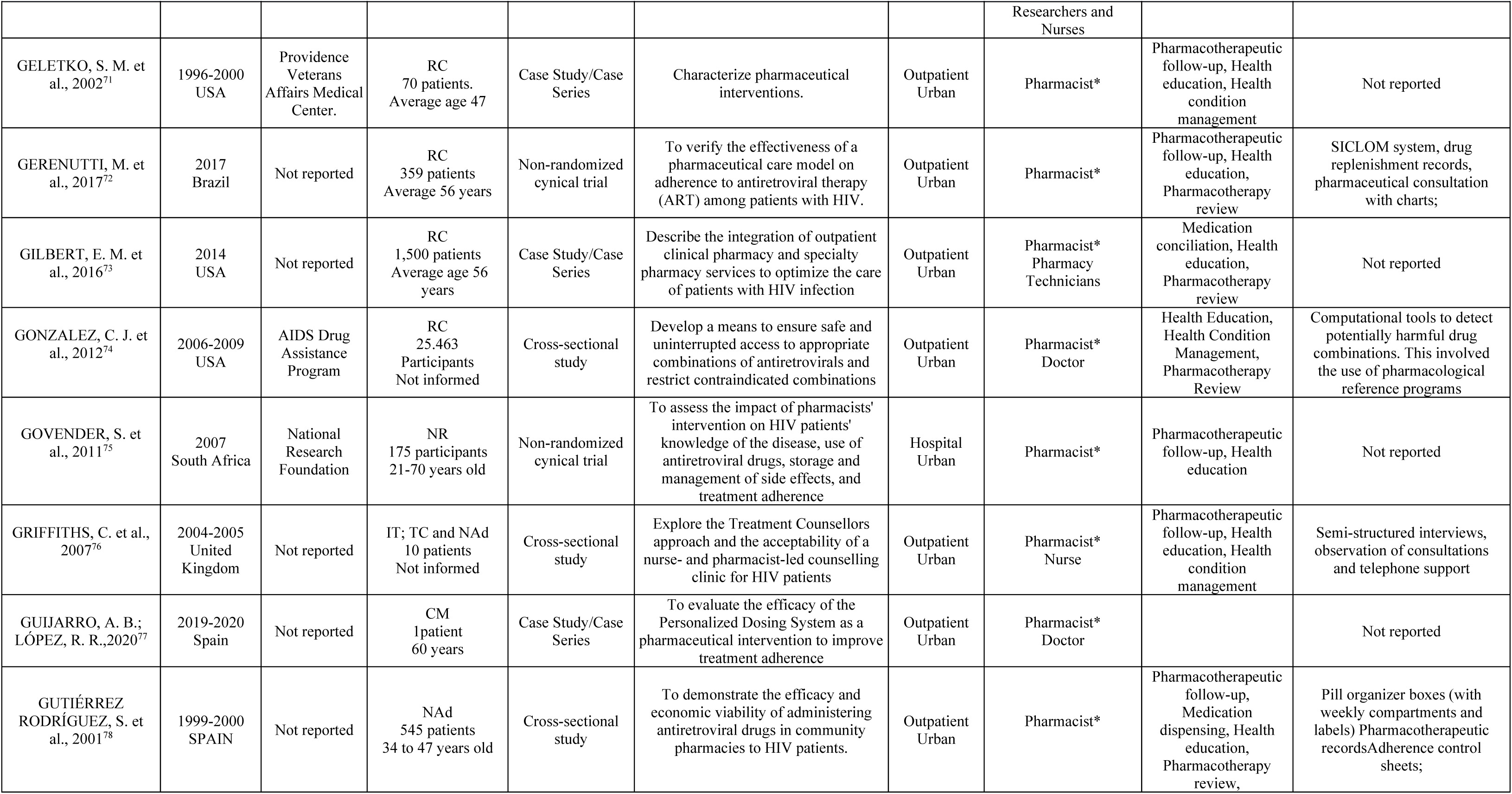
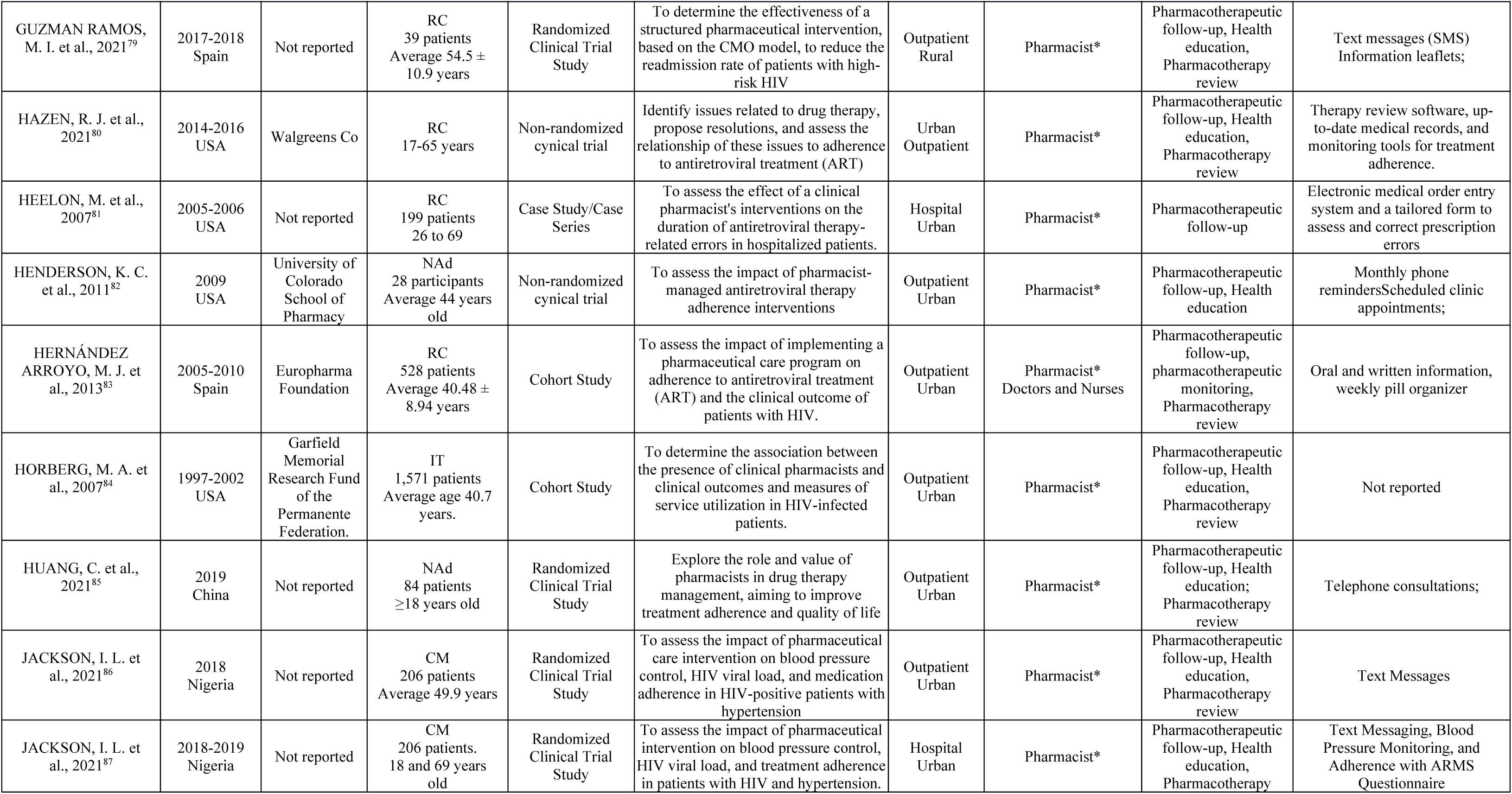
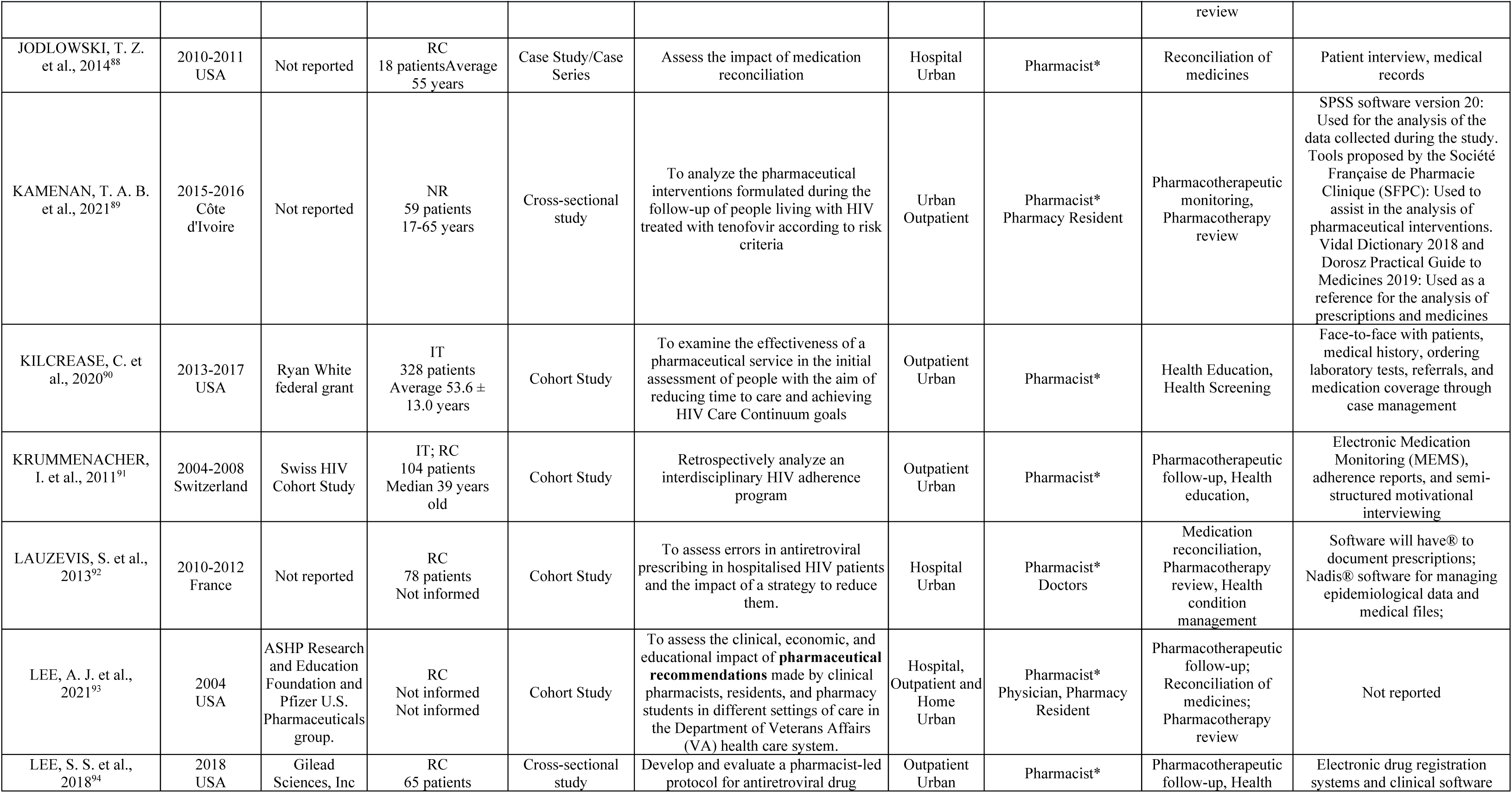
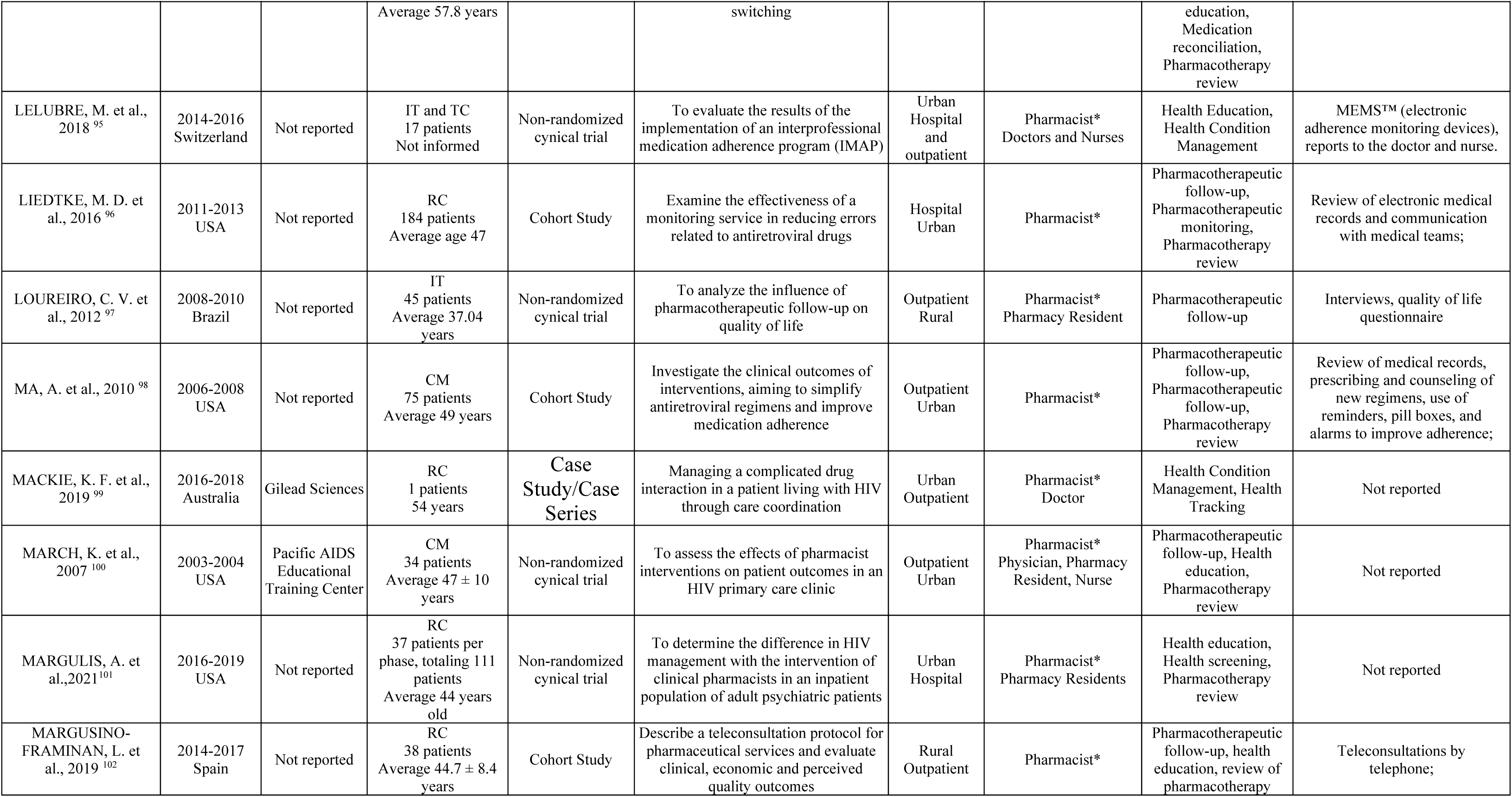
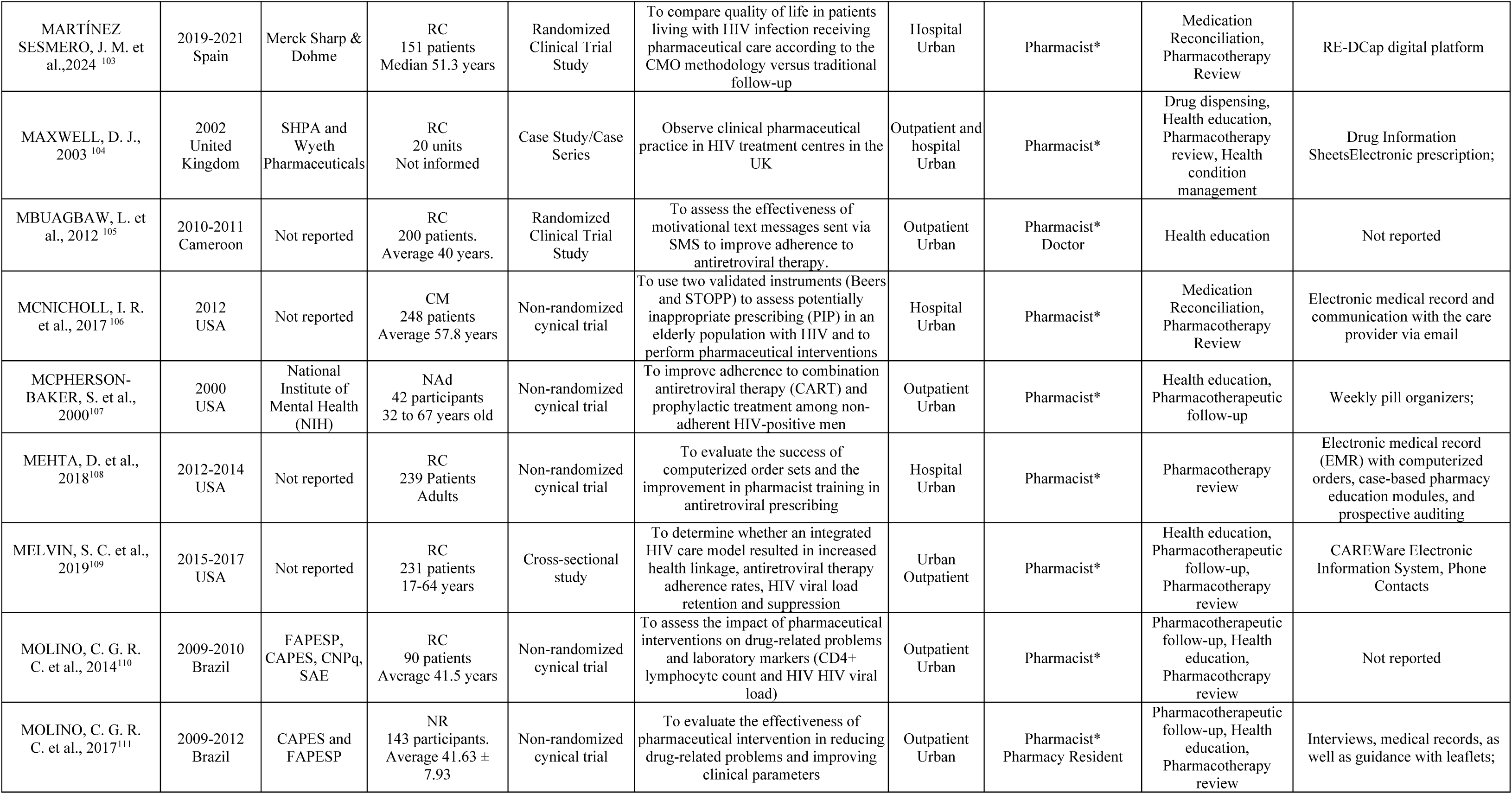
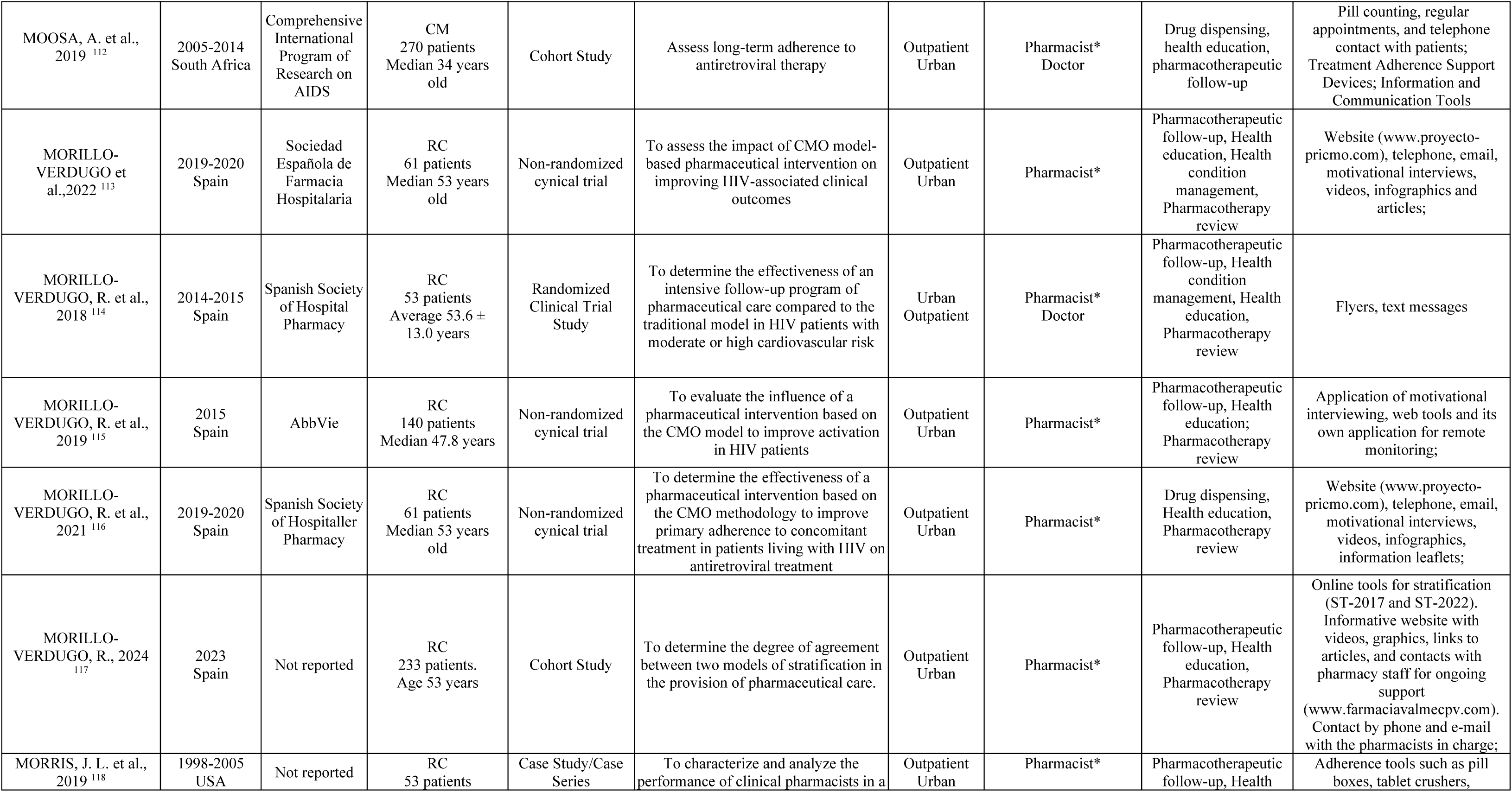
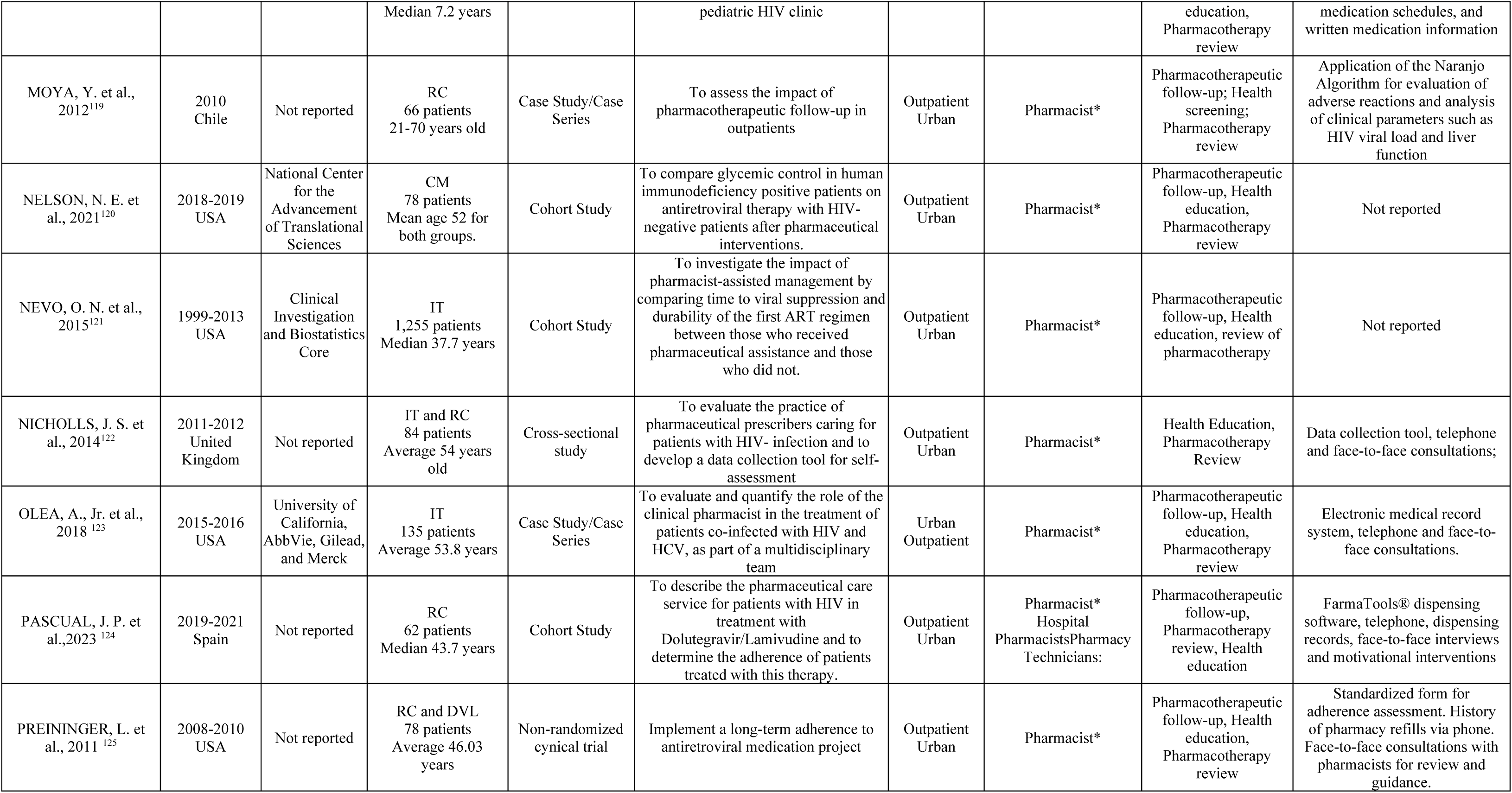
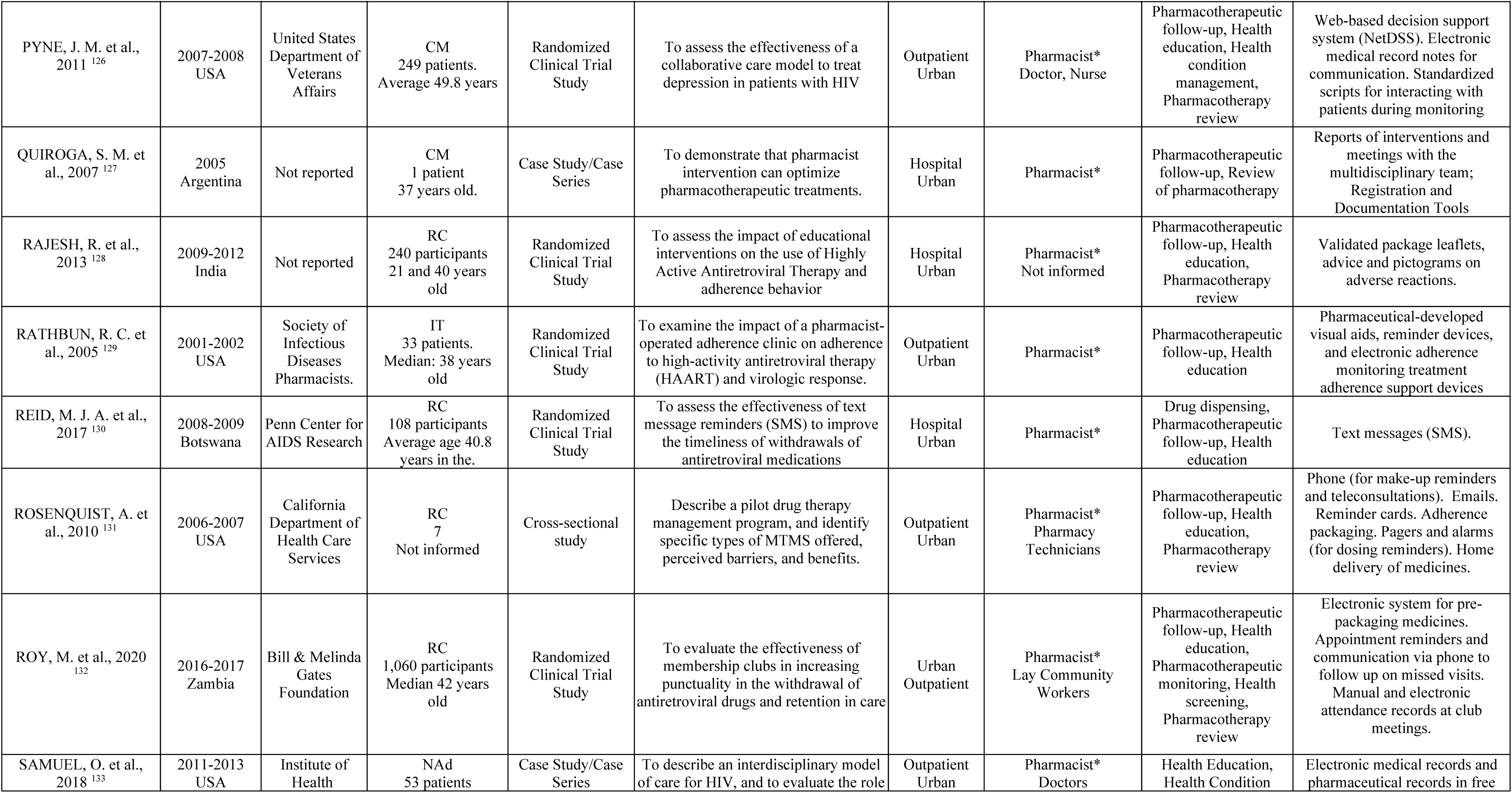
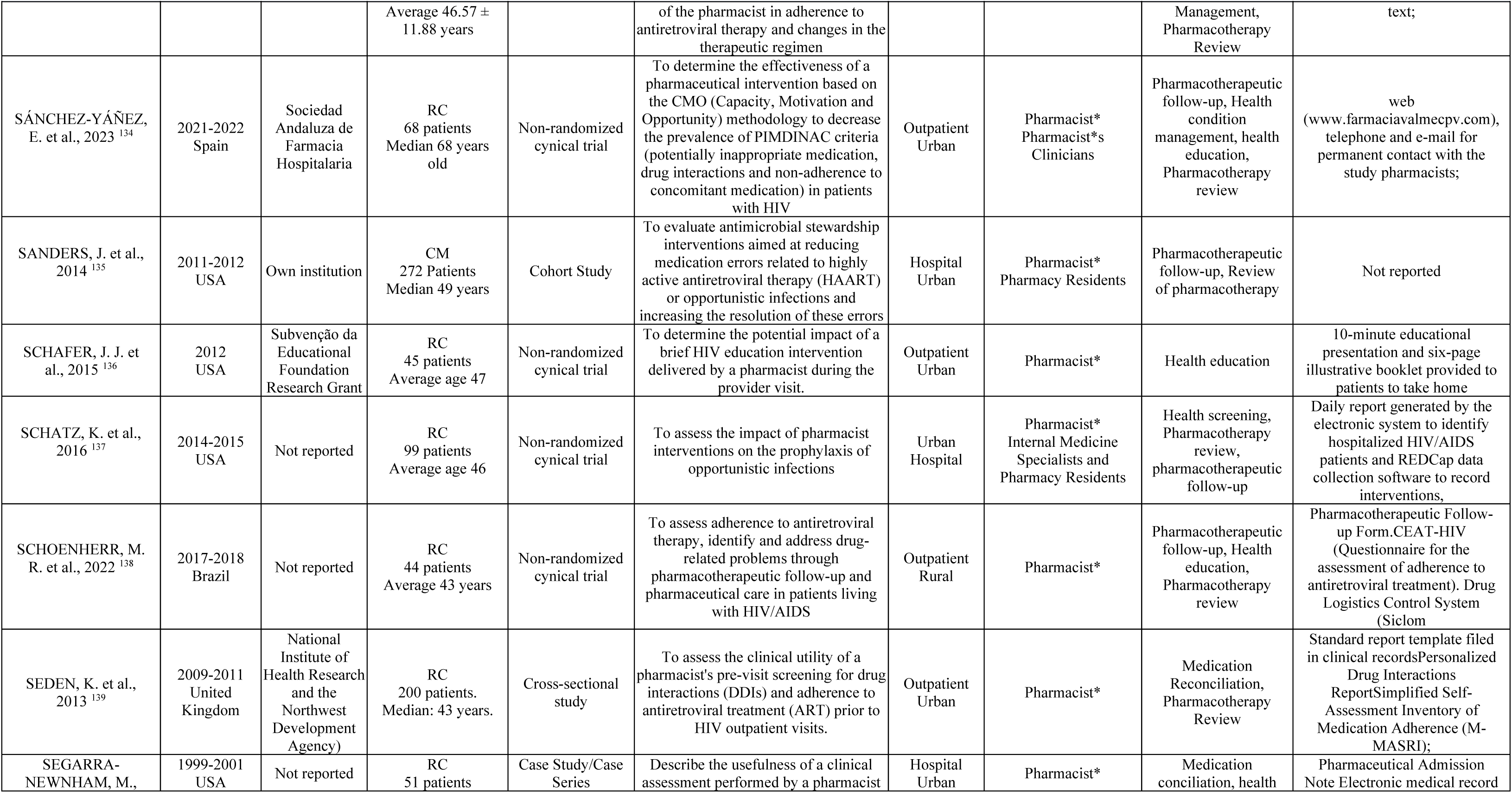
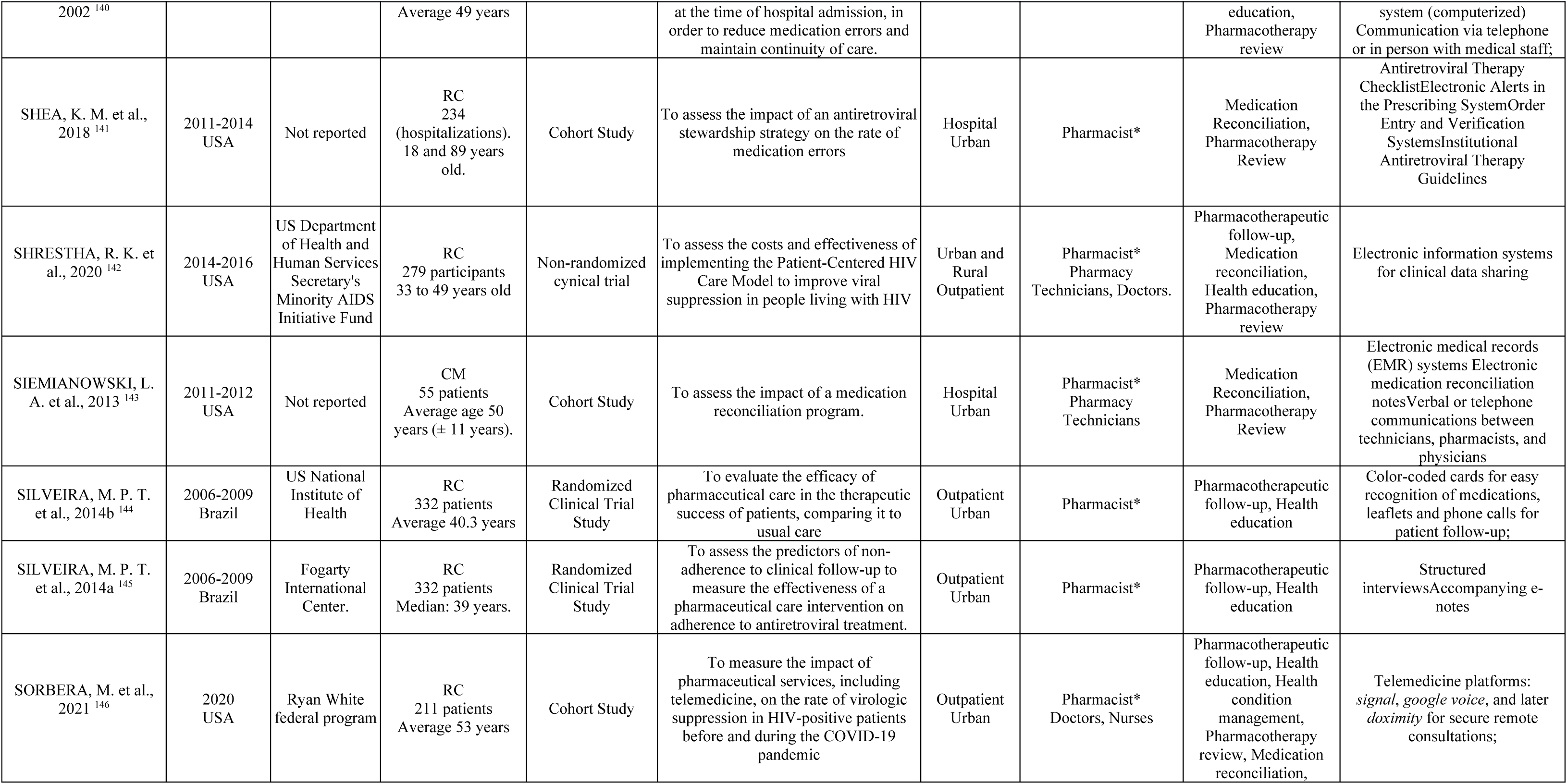
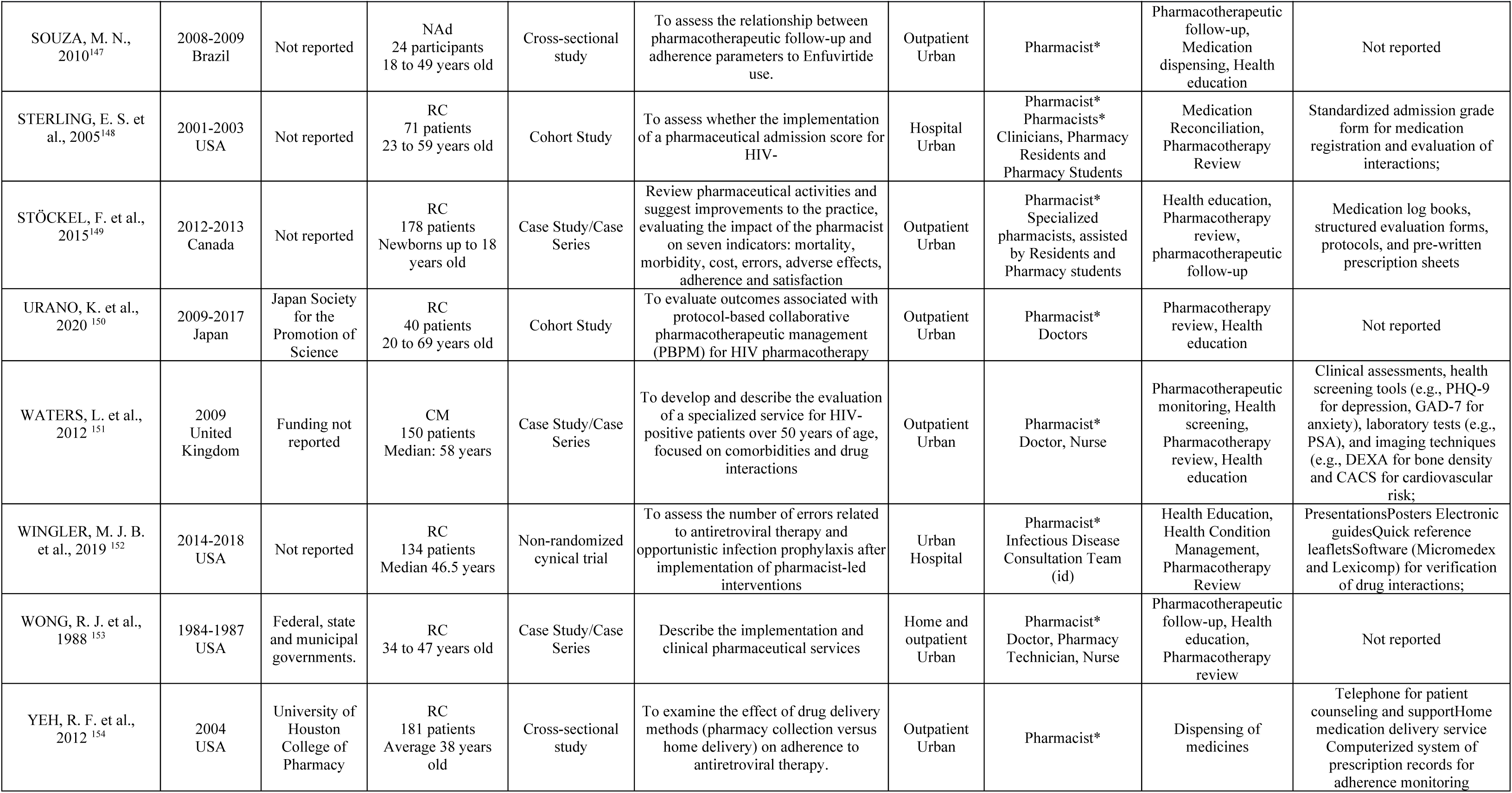
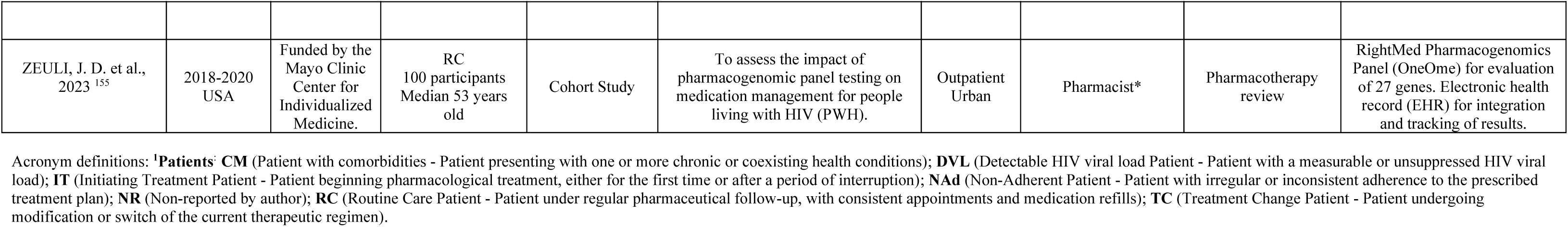
Characteristics of the primary studies included (n = 132)

**Table 2.** Characterization of Included Review Studies (n = 14)

| Study | Financing | Study Design (N studies) | Research Question | Type of Clinical Pharmaceutical Service Studied | Outcomes studied | Key Findings | Assessment of Quality or Risk of Bias |
| --- | --- | --- | --- | --- | --- | --- | --- |
| AHMED, A. et. al.,2021<sup>17</sup> | Not reported | Systematic Review 4 | What is the economic impact of pharmacy care services for HIV Long-Term Survivors? | Health Education, Health Tracking; Pharmacotherapy Review | Improvement in Quality of Life; Findings Economic | Pharmacist-managed services show cost-effectiveness to improve health outcomes for patients living with HIV/AIDS | CHEERS |
| AHMED, A. et. al.,2022<sup>15</sup> | Not reported | Systematic Review 25 | What is the effect of pharmaceutical care on adherence to antiretroviral therapy, suppression of HIV viral load and increase in CD4-t lymphocytes in HIV Long-Term Survivors? | Pharmacotherapeutic Follow-up; Health Education; Pharmacotherapy review | CD4+ count; HIV viral load; Adherence | Pharmaceutical interventions improve adherence, HIV viral load suppression, and cd4-t count in HIV Long-Term Survivors, but future high-quality studies are necessary to confirm these results | GRADE System |
| ALBUQUERQUE, A.S.A.; MARIE L. A, 2022<sup>156</sup> | Not reported | Integrative Review 11 | What is the importance of pharmaceutical assistance in the treatment with antiretrovirals? | Pharmacotherapeutic Follow-up; Health Education | Improvement in Quality of Life; Adherence | Pharmaceutical assistance is essential to support patients in adhering to antiretroviral treatment, improving their quality of life and encouraging correct treatment follow-up. |  |
| HIGGINS, N. et. al.,2009<sup>157</sup> | Ministère De La Santé Et Des Services Sociaux Du Québec. | Narrative Review 65 | - | Pharmacotherapeutic Monitoring, Pharmacotherapeutic Monitoring | CD4+ count; HIV viral load; DRP Resolution | Therapeutic drug monitoring (TDM) services have been essential to optimize the efficacy and safety of antiretroviral treatment in people living with HIV, especially in vulnerable groups. By allowing the personalization of doses based on individual characteristics, the program contributed to reducing variability between patients and improving clinical outcomes. | Not reported |
| HILL, L.A.et.al.,2019<sup>158</sup> | Not reported | Narrative Review 30 | What is the role of the clinical pharmacist in the management of people living with HIV in the modern era of antiretroviral therapy? | Health Education, Health Condition Management, Pharmacotherapy Review, Medication Reconciliation | CD4+ count; HIV viral load | The role of the clinical pharmacist is fundamental in the modern era of antiretroviral therapy, with positive impacts on HIV management, treatment adherence and control of the comorbidities. | Not reported |
| KUO, A.P. et. al.,2022<sup>159</sup> | Not reported | Scoping Review 28 | What is the effectiveness, feasibility and acceptability of HIV service delivery in private pharmacies in sub-Saharan Africa? | Health Education | Other | Private pharmacies are a promising alternative for HIV service delivery, especially in urban areas, and can offer convenience and stigma reduction compared to public clinics. | (Rob2) For Randomized Trials and Robins-I For Studies Non-randomized. |

| Study | Financing | Study Design (n studies) | Research Question | Type of Clinical Pharmaceutical Service Studied | Outcomes studied | Key Findings | Assessment of Quality or Risk of Bias |
| --- | --- | --- | --- | --- | --- | --- | --- |
| NEWMAN, T.V. et. al.,2020 <sup>160</sup> | Not reported | Overview 15 | What is the impact of community pharmacist-led interventions on chronic disease management on clinical, service utilization, and economic outcomes? | Pharmacotherapeutic Follow-up; Health Education; Review Pharmacotherapy | Economic Results |  | AMSTAR |
| ROCHA, B.et. al.,2015 <sup>161</sup> | Not reported | Systematic Review 4 | What is the impact of pharmaceutical interventions on antiretroviral therapy adherence and virologic suppression? | Pharmacotherapeutic Follow-up; Health Education | HIV viral load; Adherence | Pharmaceutical interventions can contribute to improved adherence and virologic suppression, especially in populations with low adherence and high adherence vulnerability | Cochrane Risk of Bias Tool |
| RUEDA, S. et. al.,2006 <sup>162</sup> |  | Systematic Review 19 | Supportive interventions and patient education improve adherence to high-efficacy antiretroviral therapy (HAART) for HIV Long-Term Survivors? | Pharmacotherapeutic Follow-up; Health education | Adherence | Interventions focused on practical medication management skills and individual support improve adherence to HAART. | Cochrane criteria and Declaration Consort |
| SABERI,P.et. al.,2012 <sup>163</sup> | Not reported | Systematic Review 32 | What is the impact of clinical pharmacists on multidisciplinary teams in the treatment of HIV/AIDS, especially in adherence to treatment and viral suppression? | Pharmacotherapeutic Follow-up; Health Education | Adherence; HIV viral load | Involvement of clinical pharmacists in HIV care teams may improve outcomes and contribute to greater adherence to treatment. | Cochrane Approach to Quality Assessment. |
| SILVA, APN; & SENNA JR, Va, 2021 <sup>164</sup> | Not reported | Literature Review | What is the importance of pharmaceutical care in the treatment of children infected with HIV/AIDS? | Pharmacotherapeutic Follow-up; Health Education; Dispensing Medicines | Adherence | Pharmaceutical care is indispensable in pediatric care for HIV/AIDS, ensuring greater adherence to therapy and support for families and caregivers. | - |
| SILVA, P.2022 <sup>165</sup> | Not reported | Integrative Review 5 | What is the importance of the pharmacist in adherence to antiretroviral therapy (ART) in the management of HIV/AIDS care? | Pharmacotherapeutic Follow-up; Health Education | Adherence | The pharmacist is essential to improve adherence to ART, offering direct support and continuous guidance to patients, promoting greater effectiveness in treatment. | - |
| SIMONI, J. M. et.al, 2003 <sup>166</sup> | National Institute of Health of the United States. | Narrative Review 60 | What are the most effective strategies to improve adherence to antiretroviral therapy in HIV Long-Term Survivors? | Pharmacotherapeutic Follow-up; health education; Pharmacotherapy review | CD4+ count; HIV viral load |  | Not Reported |
| TSENG A.et.al., 2013 <sup>167</sup> | Not reported | Narrative Review | What is the role of the pharmacist in the care of patients with HIV/AIDS, and what practices are recommended to optimize clinical outcomes? | Pharmacotherapeutic Follow-up; Health Education; Pharmacotherapy Review | CD4+ count, HIV viral load; Adherence; DRP Resolution | Pharmacist involvement in multidisciplinary teams significantly improves HIV patient outcomes. | Not Reported |

Acronym definitions: CHEERS (Consolidated Health Economic Evaluation Reporting Standard); DRP (Drug-Related problem); GRADE System (Grading Of Recommendations, Assessment, Development, And Evaluation); Rob2(Risk-Of-Bias Tool)

### Study Distribution and Methodological Diversity

Figure 1 depicts the geographical distribution of the 132 primary studies included in this review. The majority were conducted in North America (51.87%; n=69), highlighting this region’s predominant role in the field. Europe contributed 21.04% (n=28), followed by Africa (11.27%; n=15), South America (9%; n=12), Asia (5.2%; n=7), and Oceania (0.75%; n=1). A detailed breakdown of these studies by country, and study design is provided in Appendix 6.

**Figure 1.**
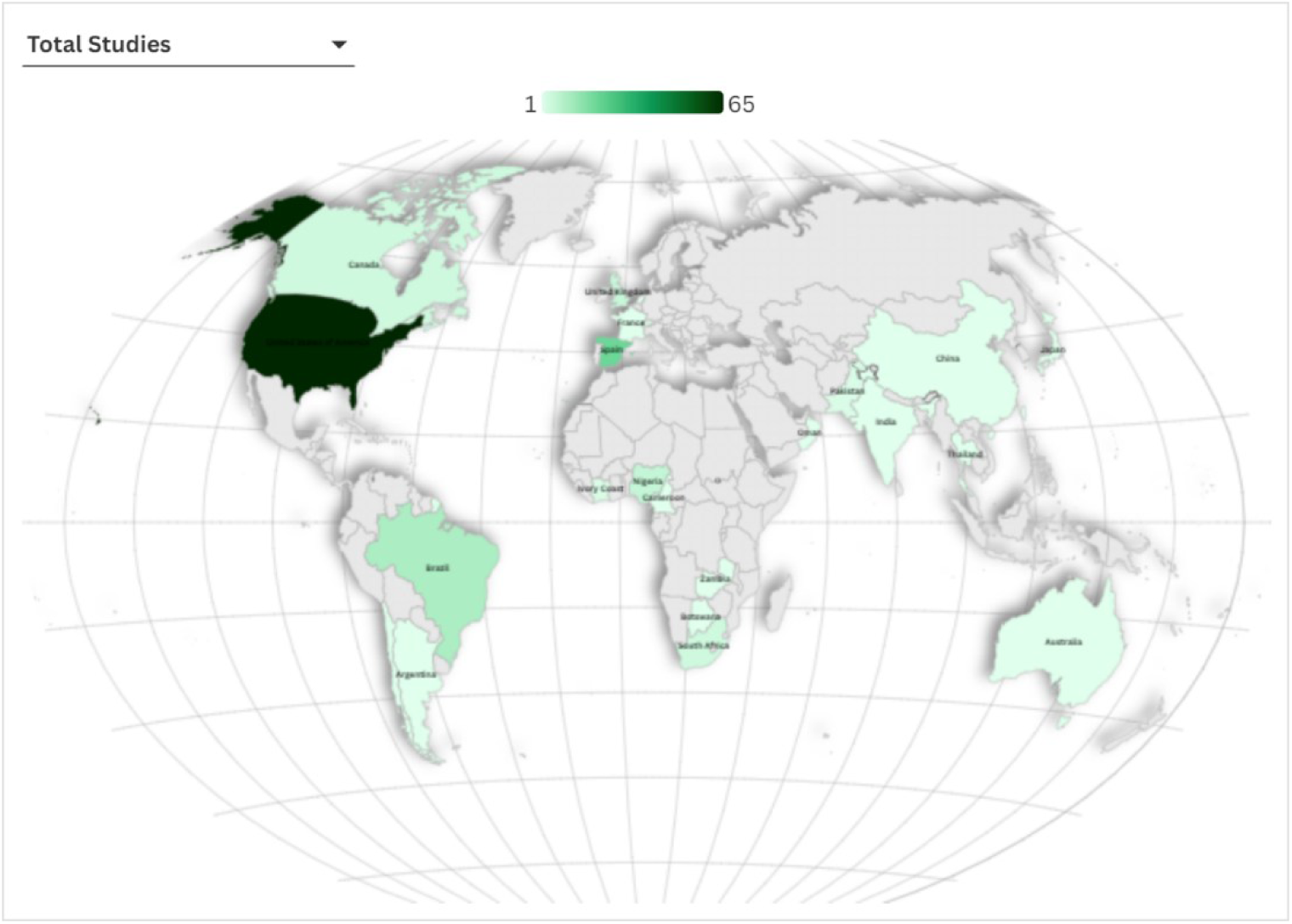
Geographic and study design distribution of primary studies (Heatmap). An interactive map version is available online: https://public.flourish.studio/visualisation/22610603/. Generated in PowerBI.®

This regional disparity in research output and methodological diversity likely stems from differences in economic resources, research infrastructure, and policy support for pharmaceutical services ^168^. Developing countries face challenges such as limited qualified pharmaceutical personnel, reduced access to medicines, and inadequate infrastructure, which hinder the expansion of clinical pharmacy services ^168,169^. Consequently, 73.6% (n=97) of the included studies originated from developed nations, with the United States (48.8%; n=65), Spain (14.25%; n=19), and the United Kingdom (3.7%; n=5) contributing the most. Among developing countries, Brazil was the most represented, accounting for 7.5% (n=10) of the studies.

Figure 2 illustrates the diverse methodological approaches among the 146 included studies, encompassing a substantial sample of 82,325 PLWH. This highlights both the breadth of data and the relevance of findings regarding clinical pharmaceutical services in HIV care.

**Figure 2.**
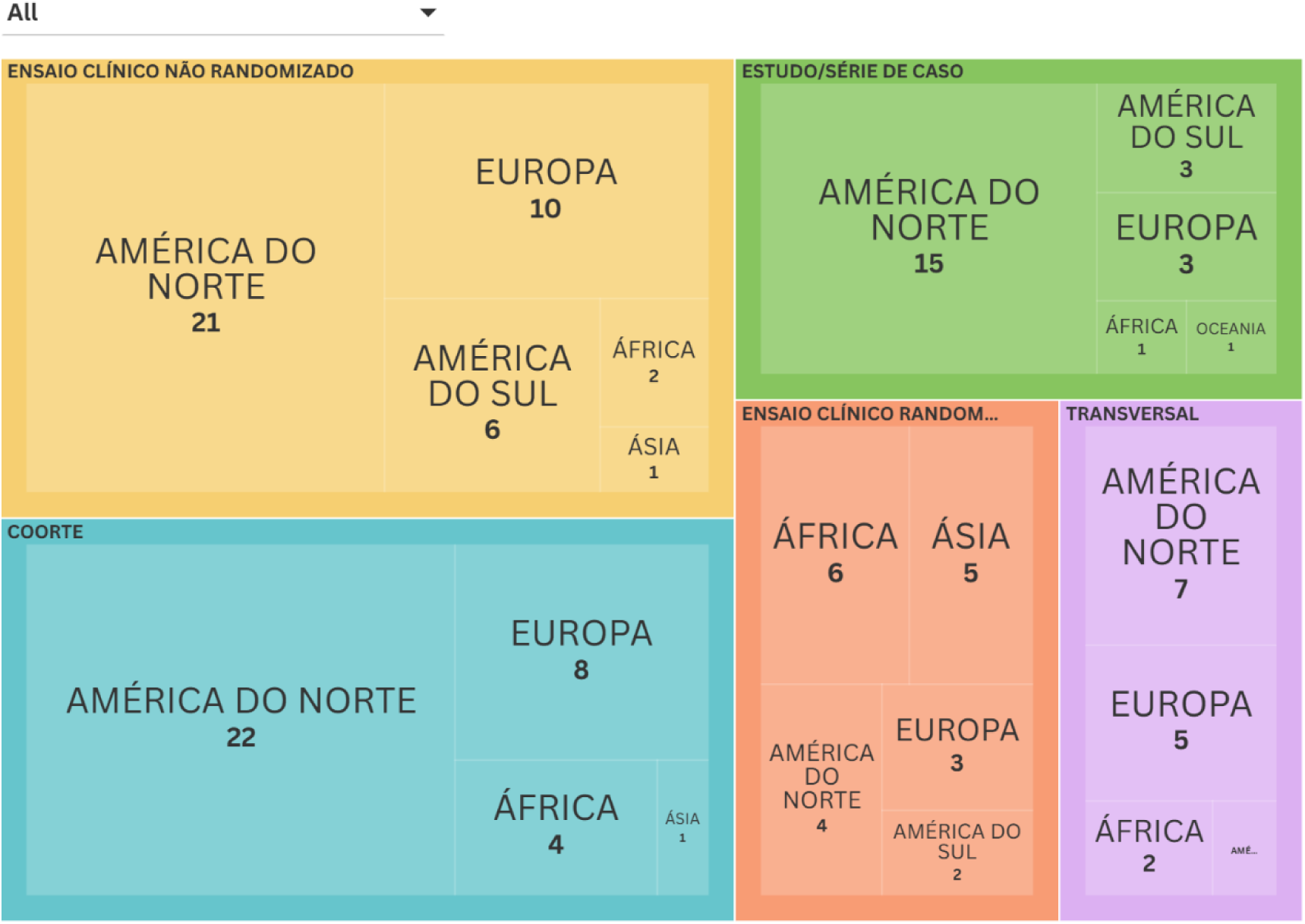
Tree Map illustrating the distribution of primary studies by region and research design An interactive version of this treemap is available online: https://public.flourish.studio/visualisation/22609493 Generated in Flourish Studio®.

Observational studies predominated, representing 48.9% (n=72), with 35 cohort studies, 23 case reports or series, and 14 cross-sectional studies. Interventional studies accounted for 42.8% (n=62), comprising 40 non-randomized controlled trials (NRCTs), most conducted in North America (n=21), and 20 randomized controlled trials (RCTs). Review studies represented 9.6% (n=14), including systematic (n=5), narrative (n=4), integrative (n=2), scoping (n=1), and literature reviews (n=1) (Table 2).

Review studies primarily addressed health education (8.8%; n=13), pharmacotherapeutic follow-up (7.5%; n=11), and pharmacotherapy review (4.1%; n=6), whereas pharmacotherapeutic monitoring, disease management, medication reconciliation, health screening, and medication dispensing were underrepresented (0.7% each; n=1). The most frequently reported outcomes were adherence (5.4%; n=8), HIV viral load (4.8%; n=7), and CD4+ count (3.4%; n=5), indicating a focus on clinical effectiveness. Other outcomes such as resolution of medication-related problems (MRPs), economic impact, and quality of life were less frequently addressed (1.4% each; n=2).

Case reports or series were concentrated in North America (n=15), with smaller contributions from Europe (n=3) and South America (n=3). Cross-sectional studies were less frequent, with representation from North America (n=7), Europe (n=4), and South America (n=1).

This review identified 20 RCTs, with a focus on health education (n=19), pharmacotherapeutic follow-up (n=16), and pharmacotherapy review (n=11), laying the groundwork for future systematic reviews that could provide robust evidence on the effectiveness of clinical pharmaceutical services. Notably, RCTs were most frequent in Africa (n=6), followed by Asia (n=5) and North America (n=4), reflecting the continent’s substantial HIV burden, accounting for approximately 67% of global infections ^170^, and highlighting the need for rigorous evaluation of pharmaceutical interventions to improve clinical outcomes and quality of life.

Despite progress, data on the long-term impact of clinical pharmaceutical services remain limited. Specifically, it is unclear to what extent these services contribute to sustained adherence to antiretroviral therapy (ART), reduction of HIV-related complications, and improved quality of life for PLWH.

Traditional clinical trials, designed primarily to assess intervention efficacy, may not accurately represent real-world clinical practice. Such trials often feature small, selective samples and are conducted in specialized centers, which may overestimate benefits and underestimate risks^170^. This underscores the value of pragmatic trials, which are specifically designed to evaluate intervention effectiveness in broader, real-world settings. This review identified 61 clinical trials, both inpatient and outpatient, encompassing participants of varying sexes and ages, reflecting the diversity of clinical practice. These studies were conducted in real-world contexts, without strict control over external factors, allowing clinical pharmaceutical services to be tailored to individual patient needs rather than following rigid protocols. Moreover, these studies assessed key clinical indicators such as medication adherence, quality of life, hospitalization rates, HIV viral load, and CD4+ count, all directly relevant to clinical decision-making. These characteristics demonstrate a pragmatic approach to evaluating intervention effectiveness under real-world health care conditions.

Regarding publication trends over time, Figure 3 reveals that between 1984 and 2023, research on clinical pharmaceutical services was scarce. The growth observed since 2000 aligns with the consolidation of highly active antiretroviral therapy (HAART) and the need for closer monitoring to ensure treatment adherence and minimize drug interactions and adverse effects^15^. Additionally, the evolution of international guidelines has reinforced the clinical role of pharmacists in managing chronic diseases, including HIV/AIDS, contributing to increased scientific output in this field ^158^. The past two decades have seen a marked increase in the number of studies, with peaks in 2009, 2014, and 2018, each with 11, 11, and 9 publications, respectively.

**Figure 3.**
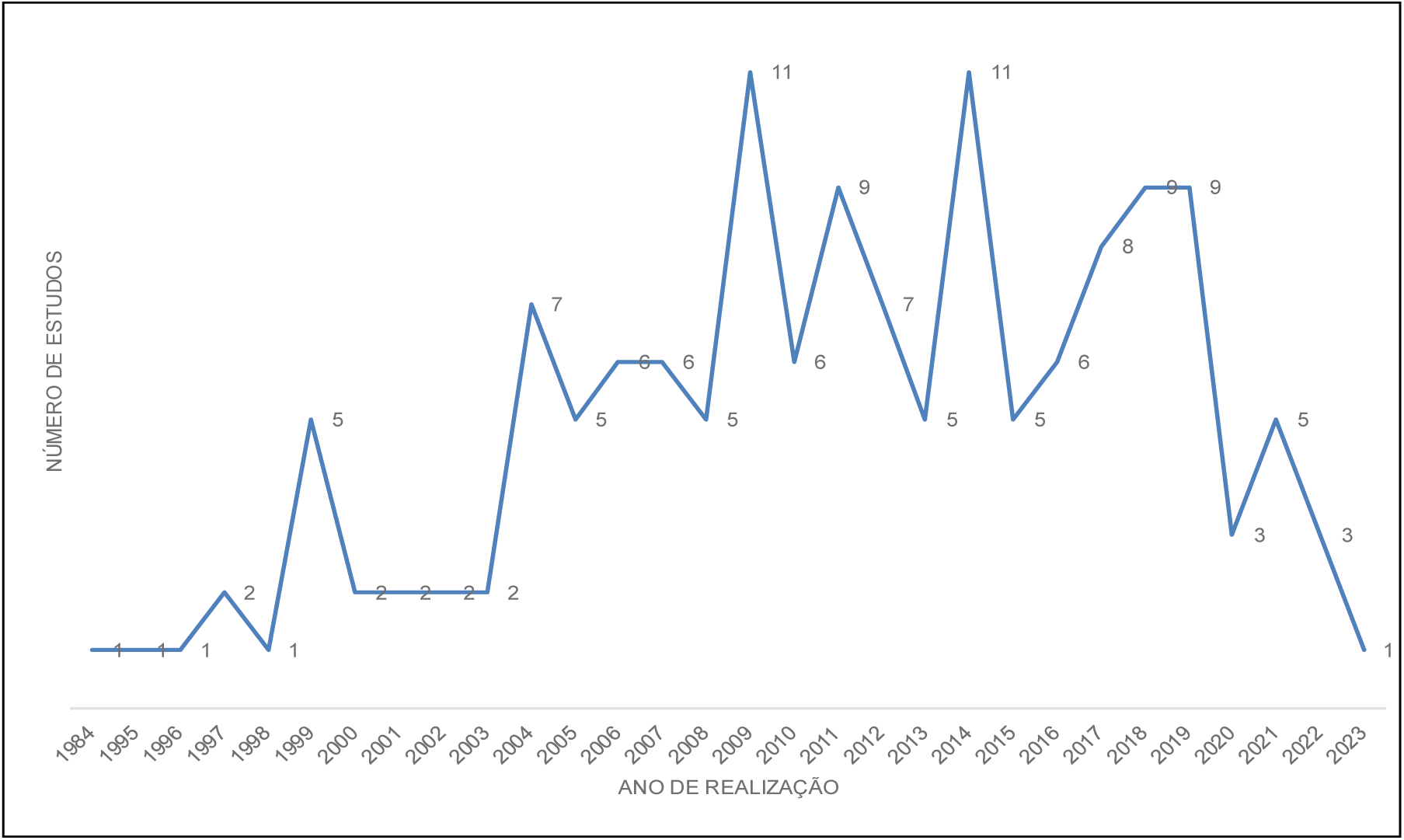
Annual Distribution of Publications on Clinical Pharmaceutical Services (1984–2023). Created in Excell ®

### Characteristics of the study participants

Table 3 summarizes the participant profiles from the 132 primary studies. The analysis revealed a diverse range of individuals involved in PLWH care. Most participants (n=57) were under regular clinical follow-up with consistent medication dispensing, suggesting the effectiveness of structured health services in maintaining ART adherence and reducing disease progression ^172,138^ further demonstrated that strong connections between patients and healthcare providers enhance treatment outcomes.

**Table 3.**
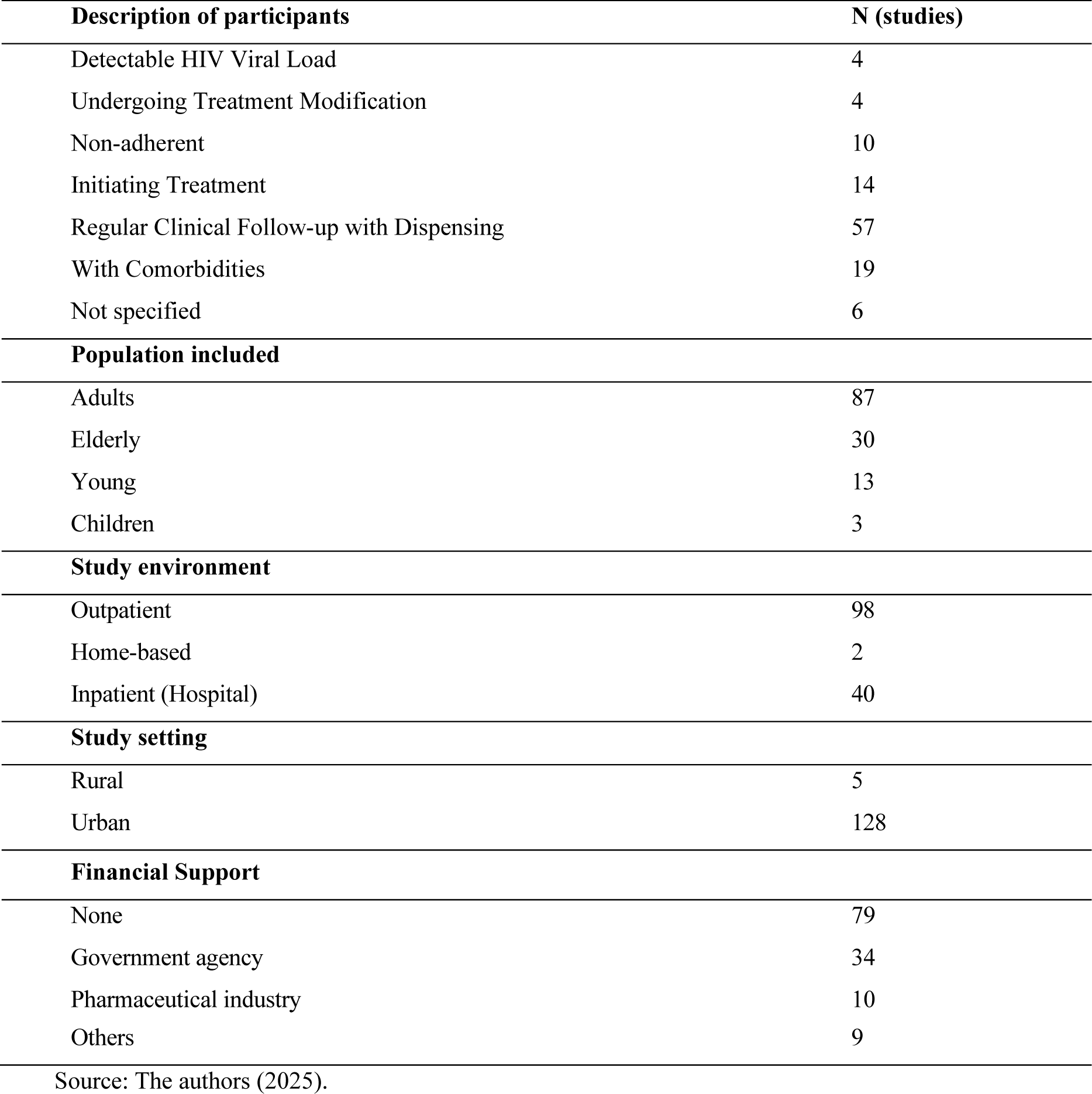
Distribution of primary studies, according to study characteristics.

| **Description of participants** | **N (studies)** |
| --- | --- |
| Detectable HIV Viral Load | 4 |
| Undergoing Treatment Modification | 4 |
| Non-adherent | 10 |
| Initiating Treatment | 14 |
| Regular Clinical Follow-up with Dispensing | 57 |
| With Comorbidities | 19 |
| Not specified | 6 |
| **Population included** |  |
| Adults | 87 |
| Elderly | 30 |
| Young | 13 |
| Children | 3 |
| **Study environment** |  |
| Outpatient | 98 |
| Home-based | 2 |
| Inpatient (Hospital) | 40 |
| **Study setting** |  |
| Rural | 5 |
| Urban | 128 |
| **Financial Support** |  |
| None | 79 |
| Government agency | 34 |
| Pharmaceutical industry | 10 |
| Others | 9 |

Source: The authors (2025).

A significant presence of patients with comorbidities (n=19) reinforces the need for interdisciplinary therapeutic strategies, given the high prevalence of cardiovascular diseases, diabetes, and neurocognitive disorders in this population (31). Early ART initiation (n=14) is also well represented, reflecting its critical role in reducing morbidity, mortality, and disease progression ^174^.

However, the underrepresentation of non-adherent patients (n=10) and those undergoing treatment modifications highlights a critical gap in understanding the factors influencing adherence and retention. This limits the development of effective strategies to improve ART continuity and outcomes.

Additionally, while the majority of studies focused on adults (n=87) and older adults (n=30), reflecting increased life expectancy with ART, younger populations were less represented (n=13), and studies on children were scarce (n=3)^118,175^. This gap may stem from the success of vertical transmission prevention programs, but it underscores the need for focused research on pediatric HIV care.

In summary, despite the breadth of participant profiles, significant gaps remain in understanding the challenges faced by non-adherent patients, those undergoing treatment changes, and pediatric populations, warranting targeted future research.

### Study environments

The analysis of study environments (Table 4) highlights a predominance of investigations conducted in outpatient (n=98) and urban settings (n=128), reflecting greater access to structured health services, often provided by advanced practice pharmacists offering comprehensive care, including medication management, specialist referrals, and access to antiretroviral therapy and combined prevention strategies such as PrEP and PEP ^176^. Conversely, rural areas were underrepresented, with only five studies, a gap likely attributable to the scarcity of healthcare professionals in these regions, limiting access to comprehensive HIV services ^177^.

Qualitative studies reinforce these findings: in rural Australia, barriers such as limited professional engagement, stigma, and logistical challenges were identified ^178^. In Africa, obstacles included transportation difficulties, provider attitudes, stigma, geographic barriers, and challenging working conditions ^179^. These factors undermine shared care effectiveness and disrupt continuity of care for people living with HIV.

Additionally, studies conducted in inpatient settings (n=40) were less frequent, and research in home-based care was rare (n=2)^11,153^, emphasizing the need to expand investigations into home and rural care contexts to better capture the experiences of PLWH.

Regarding research funding 74 studies did not report funding sources, while only 59 studies acknowledged financial support, primarily from government agencies (n=34) and the pharmaceutical industry (n=10). The lack of funding information may reflect challenges in securing research grants, potentially affecting the scope and depth of these investigations.

In summary, these findings underscore the necessity of expanding research into rural areas and household settings to comprehensively capture the diversity of experiences of individuals living with HIV/AIDS, thereby highlighting a significant data gap in contexts characterized by heightened structural vulnerability.

### Pharmaceutical Clinical Services

#### Services Studied

Figure 4 illustrates the timeline of the appearance of clinical pharmaceutical services (CPS) in the literature from 1986 onwards, highlighting the cumulative number of studies for each CPS category.

**Figure 4.**
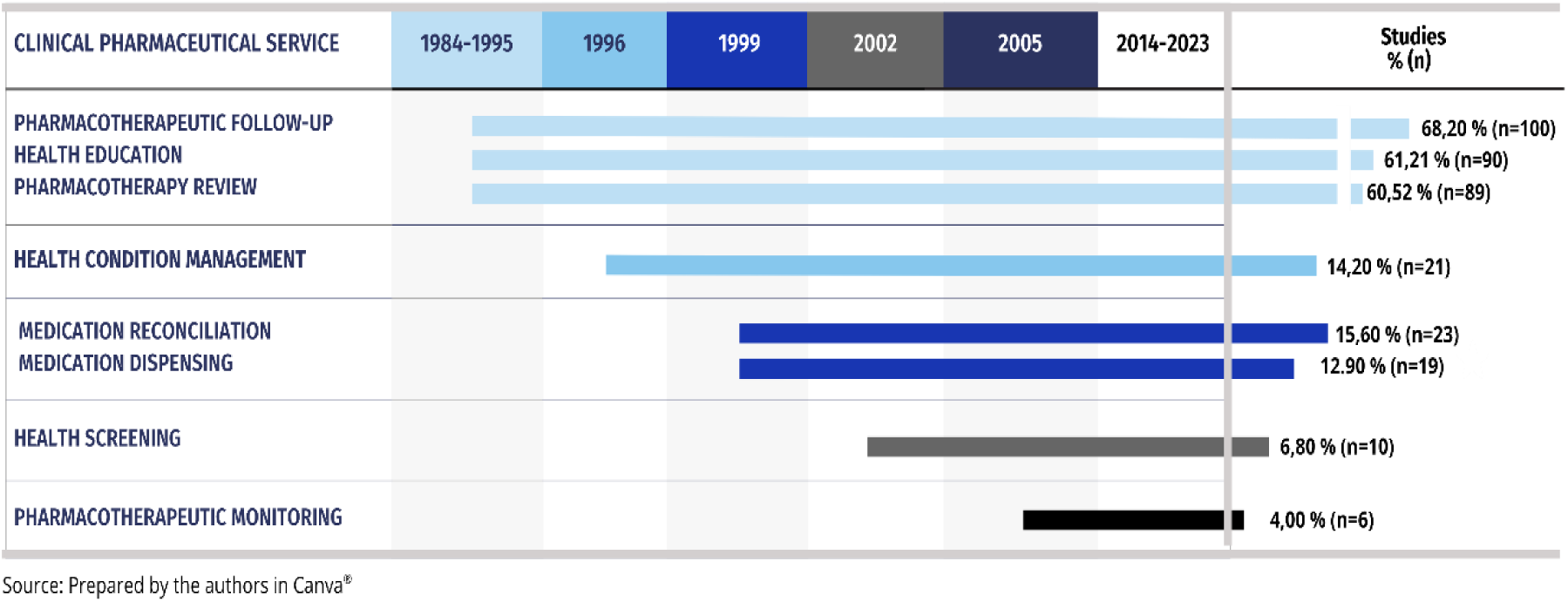
Temporal emergence of clinical pharmaceutical services in studies, and cumulative study count by service type.

The development of clinical pharmacy services for people living with HIV (PLWH) has progressed notably from the 1980s to the present, reflecting advancements in care delivery, medication management, and pharmacist integration into healthcare teams. In the early years, pharmacists’ roles were limited, largely centered on antiretroviral (ARV) therapy development. However, innovations in pharmaceutical care have since been crucial for addressing the complex needs of an aging HIV population and improving outcomes ^42^.

Between 1984 and 1995, there were only a few studies focused on pharmacotherapeutic follow-up (n=2), health education (n=1), and pharmacotherapy review (n=2), likely due to the limited therapeutic options at the time. The introduction of ART in the 1990s dramatically improved life expectancy, increasing hospitalizations for noncommunicable diseases and intensifying the need for comprehensive medication management. Pharmacists’ roles shifted from product-oriented to patient-centered models, emphasizing drug therapy management, comorbidity care, and medication reconciliation ^42,180^.

A milestone occurred in 1996 with the first publication on health condition management services (n=1) in PLWH, marking pharmacists’ integration into multidisciplinary care. From 1999 onward, studies reported on medication reconciliation (n=1) and medication dispensing services (n=1). Health screening was first examined in 2002 (n=1), and therapeutic monitoring in 2005 (n=2). Since then, no new clinical pharmaceutical services have been explored.

Additionally, Figure 4 shows that the most frequently studied clinical pharmaceutical service is pharmacotherapy review (68.02%; n=100), highlighting efforts to minimize drug interactions and optimize regimens for PLWH, particularly for those with comorbidities ^73,163^. Furthermore, studies have also focused on reducing medication-related problems ^181^.

The second most frequently studied clinical pharmaceutical service was health education, reported in 61.21% (n=90) of studies, followed closely by pharmacotherapeutic follow-up in 60.52% (n=89). Services such as medication reconciliation (15.6%; n=23), health condition management (14.2%; n=21), and medication dispensing (12.9%; n=19) were less prevalent but highlight the expansion of pharmaceutical care beyond ARV dispensing, despite the latter’s continued prominence.

More limited attention was given to clinical health screening services (6.8%; n=10) and therapeutic drug monitoring (TDM) (4.0%; n=6) ^46, 151, 83, 29, 96, 89^. The underrepresentation of TDM may be attributed to the high material and technical demands required for its implementation, despite its demonstrated accuracy and effectiveness in supporting therapeutic success and treatment adherence ^182^.

### Instruments used in CPS

Figure 5 presents the analysis of instruments employed in clinical pharmaceutical services. Registration and documentation tools were the most frequently reported, used in 61 studies, underscoring the importance of structured documentation for ensuring traceability, quality of care. Given that clinical pharmaceutical services are complex interventions, all processes must be well defined and standardized to enable reproducibility and outcomes evaluation ^183^.

**Figure 5.**
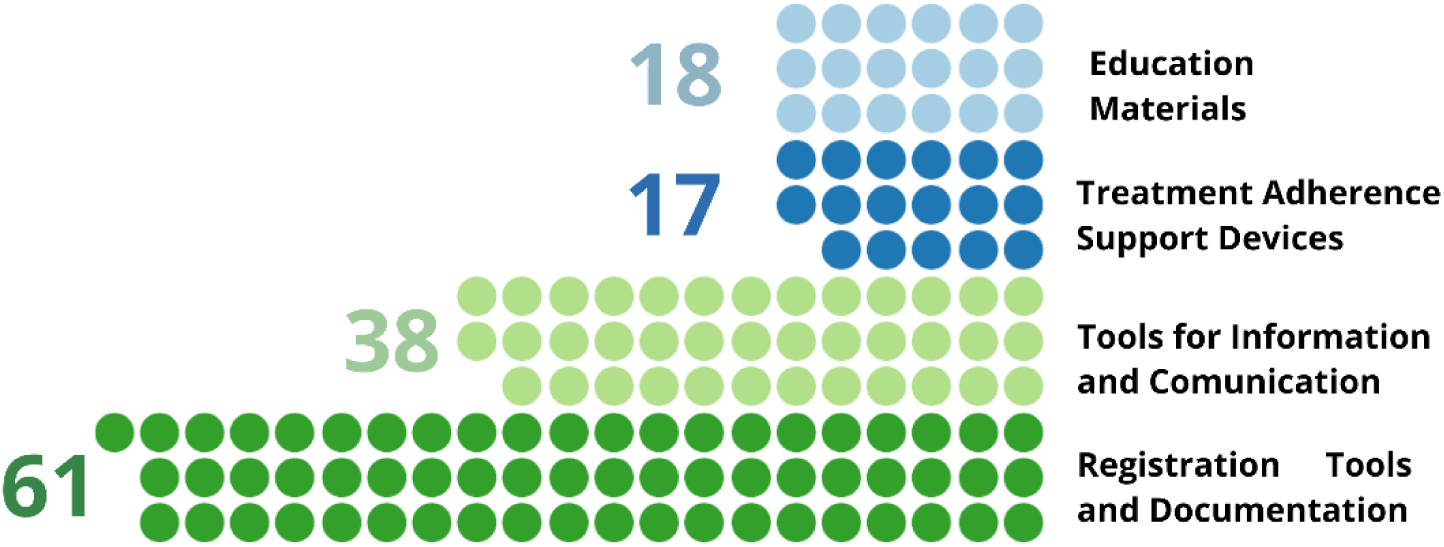
Distribution of instruments cited in the studies, and used in the pharmaceutical clinical services. Generated using Flourish Studio ®

Information and communication technologies appeared in 38 studies, reflecting their growing role in facilitating professional–patient interactions. Treatment adherence support tools (n=17) and patient education materials (n=18) were less commonly reported but underscore efforts to enhance correct drug use. These findings highlight the diversity of instruments used in clinical practice and their relevance to the effectiveness of pharmaceutical services ^184^.

### Team Organization

The role of pharmacists in the care of PLWH has advanced considerably. For instance, in Brazil, since 2023, pharmacists have been authorized not only to dispense antiretrovirals (ARVs) but also to prescribe pre-exposure prophylaxis (PrEP) and tuberculosis prophylaxis as part of combined prevention strategies ^185^.

Despite these advances, challenges persist, including the need for continuous professional development and stronger integration of pharmacists into multidisciplinary teams. The evolution of clinical pharmaceutical services is ongoing, aligning with changes in HIV treatment and highlighting the value of collaborative care models ^42^.

Figure 6 illustrates pharmacist’s participation in various team configurations across the studies analyzed. Most activities (n=95) were conducted solely by pharmacists, reflecting a growing reliance on pharmacist-led care models—particularly in response to physician shortages and the increasing number of PLWH with comorbidities. These pharmacist-led services have demonstrated high user acceptance and positive feedback ^176^.

**Figure 6.**
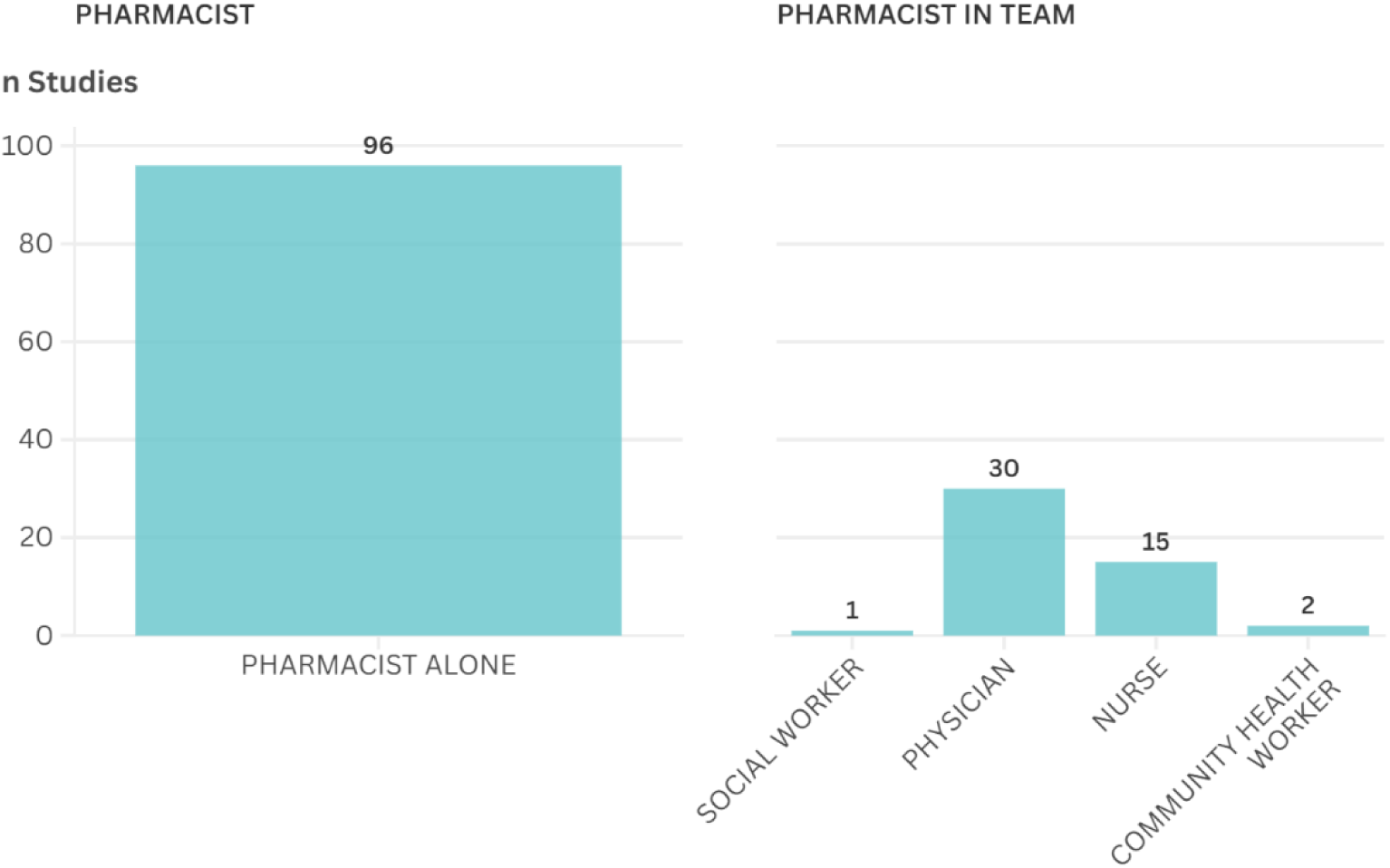
Pharmacist participation in clinical services teams in the included studies. Generated using Flourish Studio®.

Collaborative efforts were observed with physicians in 30 studies, with nurses in 15, and with community health agents in only 2 ^69,132^. Only one study reported collaboration with a social worker. Such integration into multidisciplinary teams, supported by evolving international guidelines, has reinforced the clinical role of pharmacists in managing chronic conditions like HIV/AIDS, enhancing ART adherence, managing drug interactions, and optimizing therapy ^186^.

### Outcomes studied

Figure 7 highlights the most frequently studied outcomes in the included primary studies. Medication-related problem solving (MRPs) was the predominant focus (n=78), followed by treatment adherence (n=72), HIV viral load reduction (n=44), and CD4 count improvement (n=32), outcomes known to improve significantly with pharmaceutical interventions ^15^. These findings demonstrate a research priority on immune response and disease control in PLWH.

**Figure 7.**
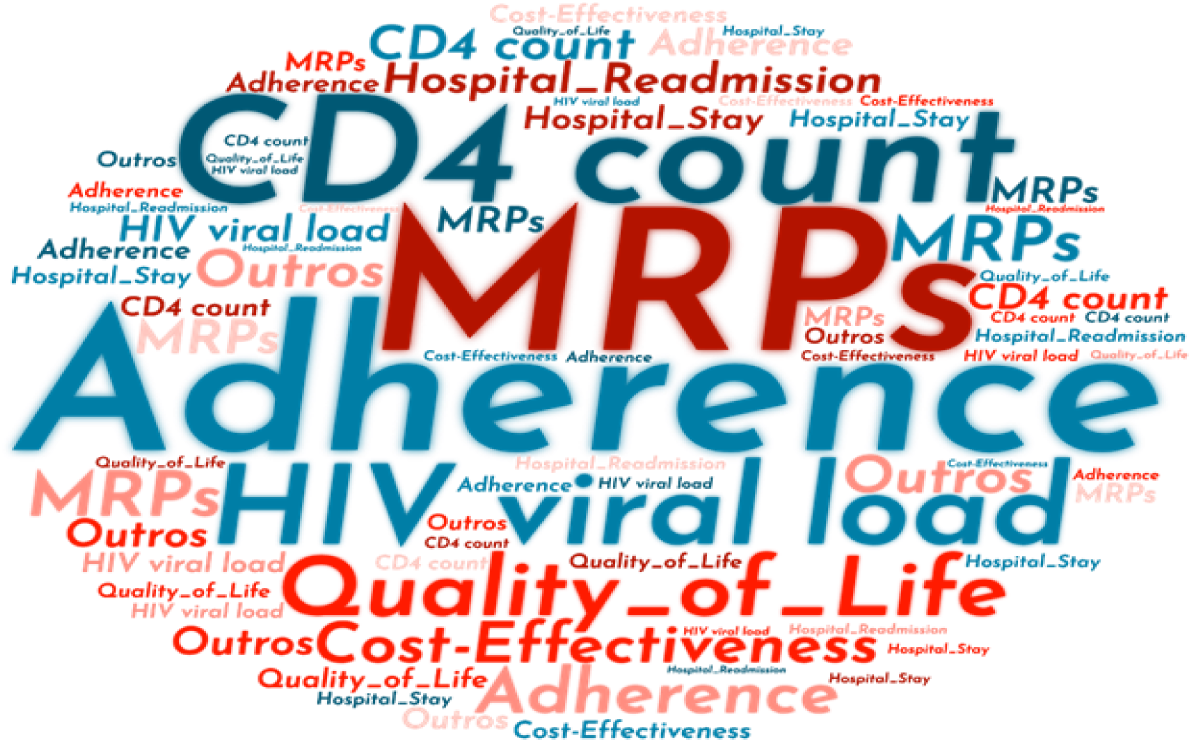
Word cloud illustrating the outcomes investigated in the primary studies included in this review. Generated with Wordclouds.com

Quality of life was assessed in 19 studies, reflecting an increasing focus on patient well-being. However, economic outcomes were reported in only 13 studies, and impacts on hospital management, such as reduced readmission rates, were rare (n=4), highlighting significant gaps in the evaluation of broader impacts of pharmaceutical services.

An integrative review of 22 articles identified that trust in healthcare providers, including pharmacists, and spirituality were the domains with the highest quality-of-life scores for people living with HIV, while environment and confidentiality concerns were associated with the lowest scores ^187^. These results emphasize the need for pharmaceutical care to address quality of life, which directly influences clinical and laboratory outcomes ^1^.

Despite these advances, substantial gaps persist, particularly in ensuring equitable access to pharmaceutical services globally. Research has highlighted the need for studies on geographical, economic, and social barriers, especially in low-income settings, and for strategies to expand service coverage in vulnerable communities, essential for improving HIV treatment outcomes ^188^.

## STRENGTHS AND LIMITATIONS

A major strength of this scoping review lies in its comprehensive and systematic literature search, which encompassed four international and multidisciplinary scientific databases, as well as gray literature through ProQuest, a broad international repository, and LA Referencia, which covers publications from Spain and Latin America. This approach ensured extensive regional representation and enhanced the robustness of the findings. Additionally, strict adherence to established guidelines for conducting and reporting scoping reviews, alongside rigorous data synthesis methods, further reinforces the credibility and comprehensiveness of this work.

A key limitation was the inability to retrieve 7 studies for full-text analysis during Phase 2, despite exhaustive efforts, including direct author contact, searches in local and remote libraries, and assistance from librarians. The absence of these studies may have influenced the comprehensiveness of the findings, potentially restricting the identification of relevant evidence and the generalizability of the results.

## CONCLUSION AND SUGGESTION FOR FUTURE STUDIES

To the best of our knowledge, this scoping review represents the first global mapping and description of clinical pharmaceutical services (CPS) in the care of people living with HIV (PLWH), as of March 2025. It highlights the breadth of investigations involving 82,325 patients and offers valuable insights into the current landscape of CPS for this population.

The analysis underscores the pivotal role of CPS in promoting adherence to antiretroviral therapy, ensuring pharmacotherapy follow-up, and reducing medication-related problems. Among the most frequently reported services, pharmacotherapy review, health education, and pharmacotherapeutic follow-up emerged as central interventions, all demonstrating a positive impact on PLWH quality of life and treatment success.

Given the recent globally expansion of CPS, coupled with ongoing challenges in controlling the AIDS epidemic, this review not only maps the current state of research but also identifies critical gaps, offering guidance for future studies, whether primary research, systematic reviews, or overviews. Key areas for further investigation include:

a. The development and application of clinical instruments and decentralized strategies (e.g., telepharmacy and remote monitoring), as well as operational challenges in pharmacy practice.
b. Therapeutic drug monitoring, given the limited focus on pharmacokinetics in PLWH, representing a significant opportunity for advancing clinical pharmacy.
c. Research on pediatric PHIV populations, which remains scarce, especially regarding adherence, drug interactions, and long-term effects of ART.
d. The implementation and evaluation of CPS in rural and home-based settings, addressing accessibility challenges and strategies to expand service coverage.
e. Expanded research on health screening and adverse drug reaction monitoring.
f. Comprehensive overviews on CPS for health education, pharmacotherapeutic follow-up, and pharmacotherapy review, capitalizing on the existing volume of review studies.

The findings of this scoping review are expected to serve as a foundational reference for future research and as a catalyst for the refinement of care policies and practices. Ultimately, this work aims to strengthen the delivery of more efficient, accessible, and patient-centered pharmaceutical care tailored to the unique needs of HIV Long-Term Survivors.

## Author Contributions

All authors contributed substantially to the development of this systematic review in accordance with international authorship criteria. MDAV Rosa and ACS Lucas conceived the study, developed the protocol, and supervised the methodological framework. ZSJ Bessa conducted the literature search. MDAV Rosa and IAV Neves performed the study selection. MDAV Rosa and RC Badin performed the data extraction. MDAV Rosa performed the data synthesis and the interpretation of the results. MDAV Rosa drafted the manuscript and ACS Lucas and PDO Almeida critically reviewed the manuscript. All authors read and approved the final version. Each author agrees to be accountable for the accuracy and integrity of any part of the work.

## Acknowledgments

The authors would like to thank Dr. Marcelo Campese for their punctual yet valuable contributions. Although not meeting authorship criteria, their expert feedback during the methodological discussions added depth to this review.

## Ethical Considerations

This study is a secondary analysis of publicly available data and did not involve human participants or identifiable personal data. Therefore, ethics approval was not required.

## Data availability statement

The datasets generated during the current study are not publicly but may be obtained from the corresponding author upon reasonable request.

## APPENDIX 1 LIST OF SEARCH SYNTAXES

APPENDIX 1 - LIST OF SEARCH SYNTAXES.
| BASES | SEARCH EQUATION |
| --- | --- |
| LILACS | (atención farmacéutica OR servicios farmacéuticos OR servicio de farmácia OR farmácia clínica OR atenção farmacêutica OR assistência farmacêutica OR uso racional de medicamentos OR uso de medicamentos OR uso de drogas uso de medicamentos OR serviços farmacêuticos OR serviço de farmácia hospitalar OR serviço de farmácia clínica OR) OR (pharmaceutical services OR pharmaceutic services OR pharmaceutic service OR pharmacy services OR pharmacy service OR pharmaceutical care OR community pharmacy services OR pharmacist service OR pharmacy practice OR clinical pharmacy OR clinical pharmacist OR pharmacies OR pharmacist OR pharmacy OR pharmacist- patient relationship OR pharmaceutical assistance) AND (hiv OR aids OR hiv OR hiv OR long-term survivors of HIV OR long-term survivors of AIDS OR long-term survivors of AIDS OR long-term survivors HIV-1 OR HIV survivors OR women who live with HIV OR men who live with HIV OR survivors with HIV OR survivors in wide HIV-1 OR AIDS survivors OR HIV-1 survivors OR long-term survivors with HIV-1 OR long-term survivors with HIV-1 OR long-term survivors with HIV-1 OR HIV-1 survivors OR HIV-1 survivors OR plazo OR mujeres que viven con el vih OR hombres que vivencon el vih OR survivors a largo plazo de vih OR survivors a largo plazo del vih-1 OR survivors de sida a largo plazo) OR (hiv OR human immunodeficiency virus OR human immune deficiency virus OR aids OR acquired immune deficiency syndromevirus OR acquired immunodeficiency syndrome virus OR hiv long term survivors OR long-term hiv survivors OR hiv-1 long-term survivors OR hiv 1 long term survivors OR hiv-1 long-term survivorOR aids long- term survivors OR aids long term survivors OR aids long-term survivor OR hiv non progressors OR hiv non-progressor) AND (db:("LILACS")) |
| Medline/PUBMED | ("pharmacy services"[Title/Abstract] OR "pharmacy service"[Title/Abstract] OR "pharmaceutical care"[Title/Abstract] OR "community pharmacy services"[Title/Abstract] OR "pharmacistservice"[Title/Abstract] OR "pharmacy practice"[Title/Abstract] OR "clinical pharmacy"[Title/Abstract] OR "clinical pharmacist"[Title/Abstract] OR "pharmacies"[Title/Abstract] OR "pharmacist"[Title/Abstract] OR "pharmacy"[Title/Abstract] OR "pharmacist-patient relationship"[Title/Abstract] OR " pharmaceutical assistance"[Title/Abstract]) AND ("HIV"[Title/Abstract] OR "human immunodeficiency virus"[Title/Abstract] OR "human immune deficiency virus"[Title/Abstract] OR "AIDS"[Title/Abstract] OR "acquired immune deficiency syndrome virus"[Title/Abstract] OR "acquired immunodeficiency syndrome virus"[Title/Abstract] OR "hiv longterm survivors"[Title/Abstract] OR "hiv long term survivors"[Title/Abstract] OR "hiv 1 long term survivors"[Title/Abstract] OR "hiv 1 long term survivors"[Title/Abstract] OR "hiv 1 long term survivors"[Title/Abstract] OR "aids long term survivors"[Title/Abstract] OR "aids long term survivors"[Title/Abstract] OR "AIDS long-term survivor"[Title/Abstract] OR "HIV non progressors"[Title/Abstract] OR "HIV non- progressor"[Title/Abstract]) |
| Web Of Science (WOS) | TS=("pharmaceutical services" OR "pharmaceutic services" OR "pharmaceutic service" OR "pharmacy services" OR "pharmacy service" OR "pharmaceutical care" OR "community pharmacy services" OR "pharmacist service" OR "pharmacy practice" OR "clinical pharmacy" OR "clinical pharmacist" OR "pharmacies" OR "pharmacist" OR "pharmacy" OR "pharmacist-patient relationship" OR "pharmaceutical assistance") AND TS=("HIV" OR "human immunodeficiency virus" OR "human immune deficiency virus" OR "AIDS" OR "acquired immune deficiency syndrome virus" OR "acquired immunodeficiency syndrome virus"OR "HIV long term survivors" OR "HIV long-term survivors" OR "HIV-1 long-term survivors" OR "HIV 1 long term survivors" OR "HIV-1 long-term survivor" OR "AIDS long-term survivors" OR "AIDS long-term survivors" OR "HIV non-progressors" OR "HIV non-progressors") |
| SCOPUS | ( TITLE-ABS-KEY ({pharmaceutical services} OR {pharmaceutic services} OR {pharmaceutic service} OR {pharmacy services} OR {pharmacy service} OR {pharmaceutical care} OR {community pharmacy services} OR {pharmacist service} OR {pharmacy practice} OR {clinical pharmacy} OR {clinical pharmacist} OR {pharmacies} OR {pharmacist} OR {pharmacy} OR {pharmacist- patient relationship} OR {pharmaceutical assistance} ) AND TITLE-ABS-KEY ( {HIV} OR {human immunodeficiency virus} OR {human immune deficiency virus} OR {AIDS} OR {acquired immune deficiency syndrome virus} |
|  | OR {acquired immunodeficiency syndrome virus} OR {HIV long term survivors}OR {HIV long-term survivors} OR {HIV-1 long-term survivors} OR {HIV 1 long term survivors} OR {HIV-1 long-term survivor} OR {AIDS long-term survivors} OR {AIDS long term survivors} OR {AIDS long-term survivor} OR {HIV non progressors} OR {HIV non-progressor} ) ) AND ( LIMIT-TO ( DOCTYPE , "ar" ) OR LIMIT-TO ( DOCTYPE , "re" ) ) |
| PROQUEST<br>(via WOS) | TS=("pharmaceutical services" OR "pharmaceutic services" OR "pharmaceutic service" OR "pharmacy services" OR "pharmacy service" OR "pharmaceutical care" OR "community pharmacy services" OR "pharmacist service" OR "pharmacy practice" OR "clinical pharmacy" OR "clinical pharmacist" OR "pharmacies" OR "pharmacist" OR "pharmacy" OR "pharmacist-patient relationship" OR "pharmaceutical assistance") AND TS=("HIV" OR "human immunodeficiency virus" OR "human immune deficiency virus" OR "AIDS" OR "acquired immune deficiency syndrome virus" OR "acquired immunodeficiency syndrome virus" OR "HIV long-term survivors" OR "HIV long-term survivors" OR "HIV-1 long-term survivors" OR "HIV 1 long term survivors" OR "HIV-1 long- term survivor" OR "AIDS long-term survivors" or "AIDS long-term survivors" OR "HIV non-progressors" OR "HIV non-progressors" OR "HIV non-progressors") |
| LA REFERENCIA | Buscar: (Todos los Campos:pharmaceutical services OR pharmaceuti services OR pharmaceutic service OR pharmacy services OR pharmacy service OR pharmaceutical care OR community pharmacy services OR pharmacist service OR pharmacy practice OR clinical pharmacy OR clinical pharmacist OR pharmacies OR pharmacist OR pharmacy OR pharmacist- patient relationship OR pharmaceutical assistance O Todos los Campos:HIV OR human immunodeficiency virus OR human immune deficiency virus OR AIDS OR acquired immune deficiency syndrome virus OR acquired immunodeficiency syndrome virus OR HIV long term survivors OR Long-term HIV survivors OR HIV-1 long-term survivors OR HIV 1 long term survivors OR HIV-1 long-term survivor OR AIDS long-term survivor OR AIDS long-term survivors OR AIDS long-term survivor OR HIV noprogressors OR HIV non-progressor) |

Source: Prepared by the authors, 2024

## APPENDIX 2 LIST OF NON-RETRIEVED STUDIES

1. Consultant pharmacists improve patient care in HIV/AIDS group home. *Med Interface*. 1995;8(1):107,110.
2. Pharmacist counseling, use of compliance tool: Does it improve outcomes in an HIV population? *Formulary*. 1998;33(11):1144.
3. Pharmacy project assists with meds adherence. Structured pharmd involvement crucial. *AIDS Alert*. 2002;17(11):144–146.
4. Geletko SM, Segarra M, Copeland DA, Teague AC. Pharmaceutical care for hospitalized HIV-infected patients compared to infectious diseases consult patients without HIV infection. *J Infect Dis Pharmacother*. 1996;2(1):45–57. doi:10.1300/J100v02n01\_04
5. Ibarra BO, Gabilondo ZI, Mora AO. HIV patient selection for pharmaceutical follow-up. *Aten Farm*. 2009;11(2):95–100.
6. Marshall N, Harvey S, Naous N, et al. Pharmacist interventions on home delivery prescriptions in three London HIV outpatient clinics. *HIV Med*. 2013;14(Suppl 2):16.
7. Warnock AC, Rimland D. The provision of pharmaceutical care in a Veterans’ Affairs Medical Center outpatient HIV clinic. *Hosp Pharm*. 1994;29(2):114–116,119–120.

## APPENDIX 3 LIST OF EXCLUDED STUDIES AND REASONS

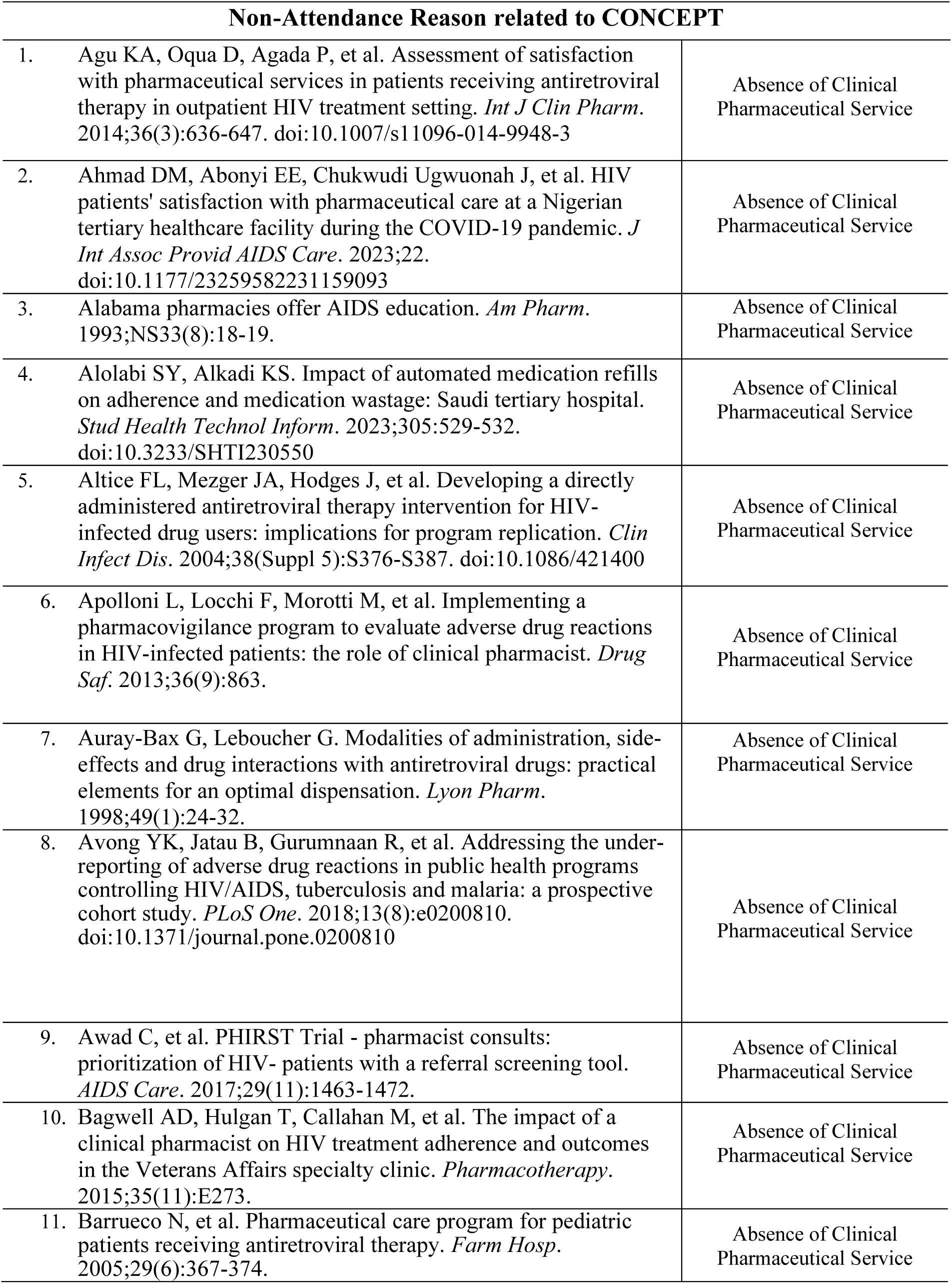
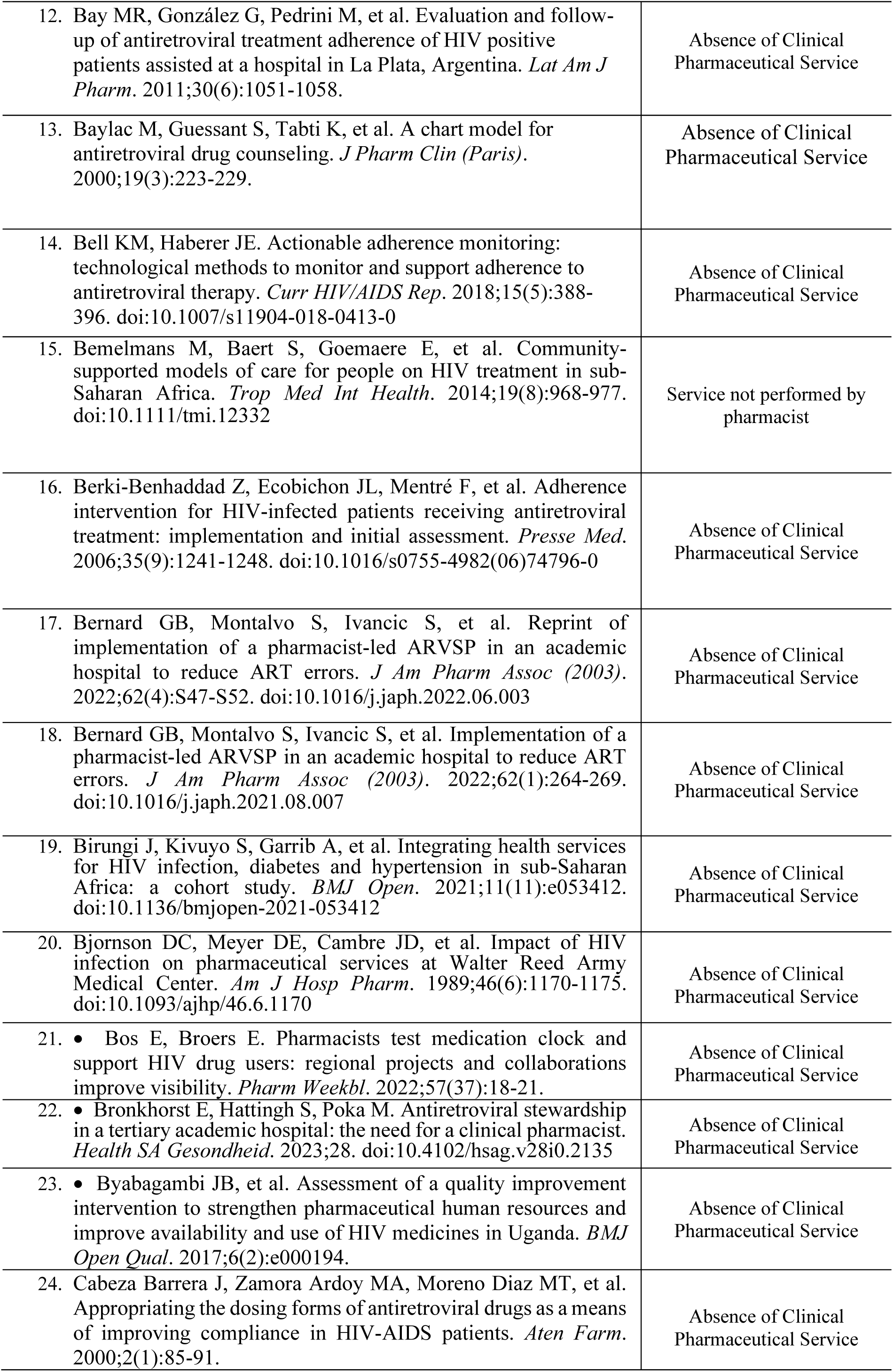
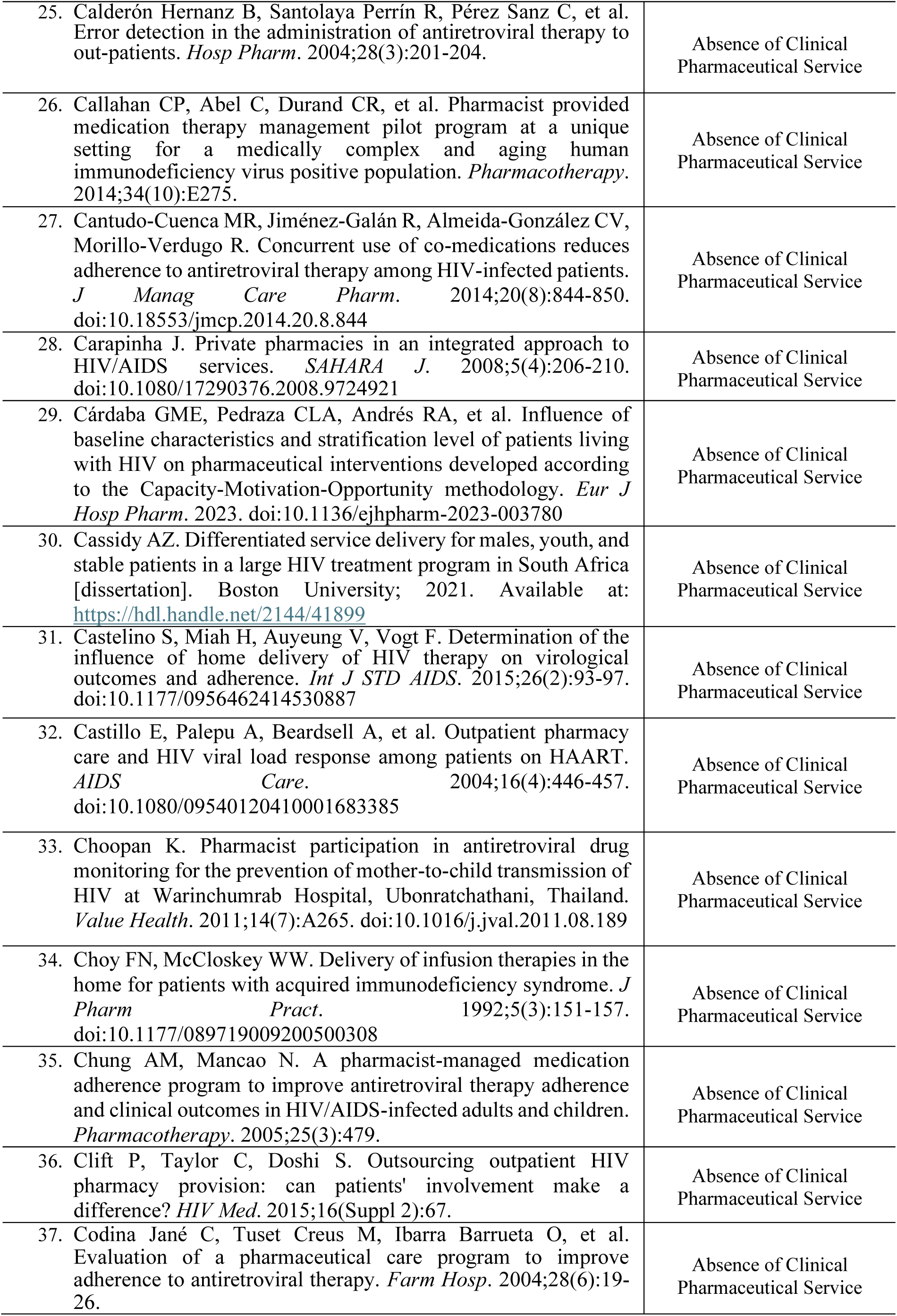
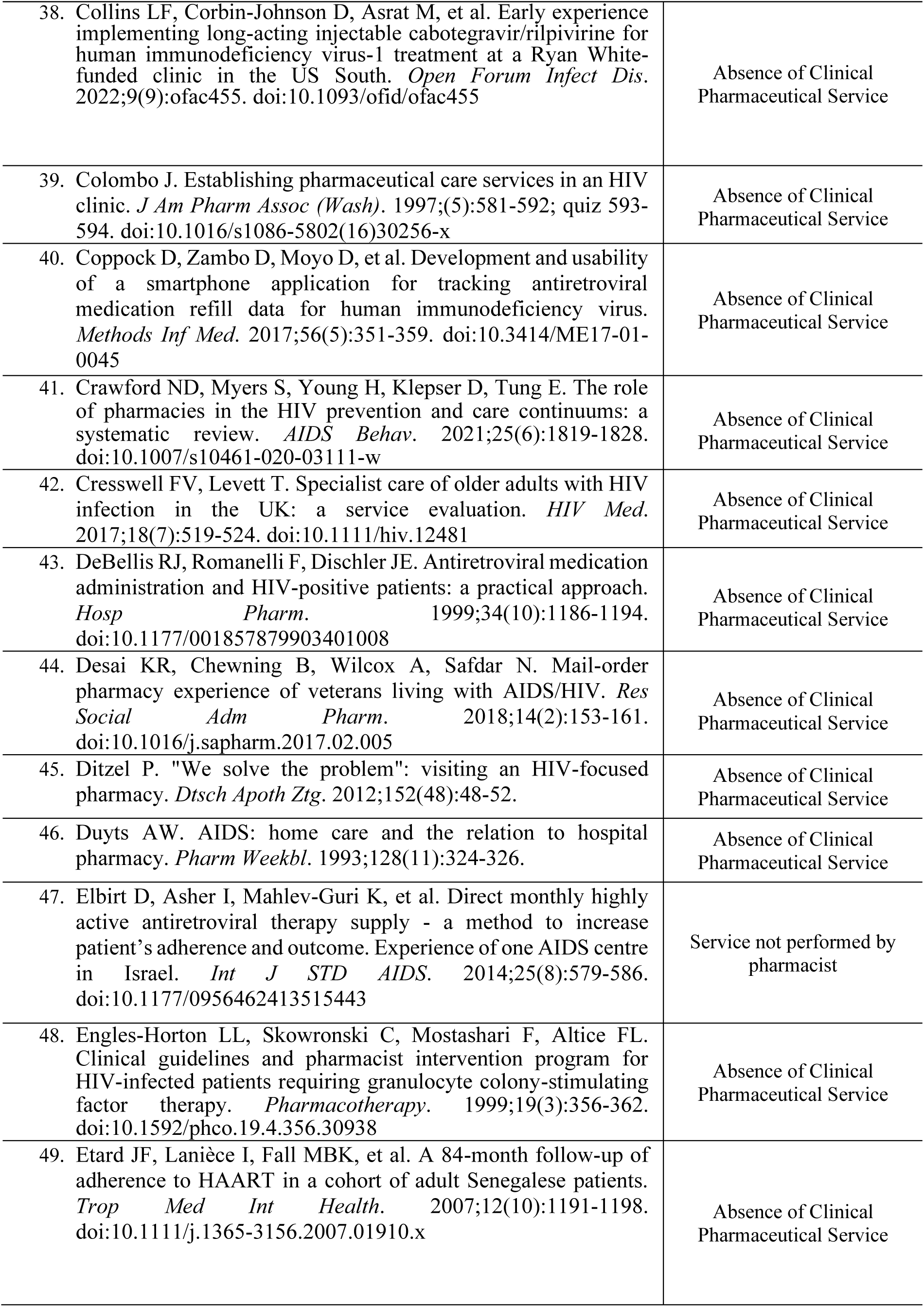
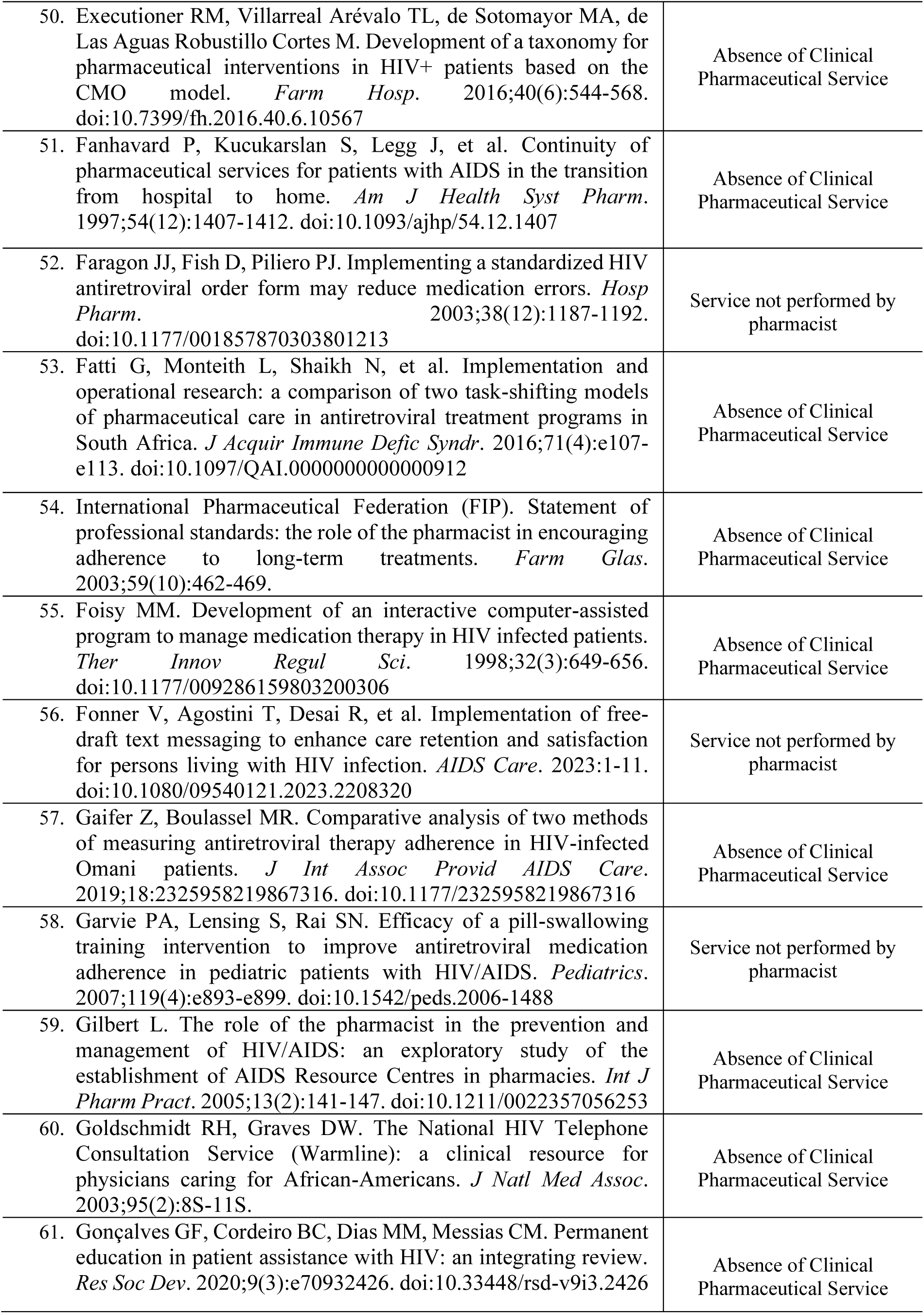
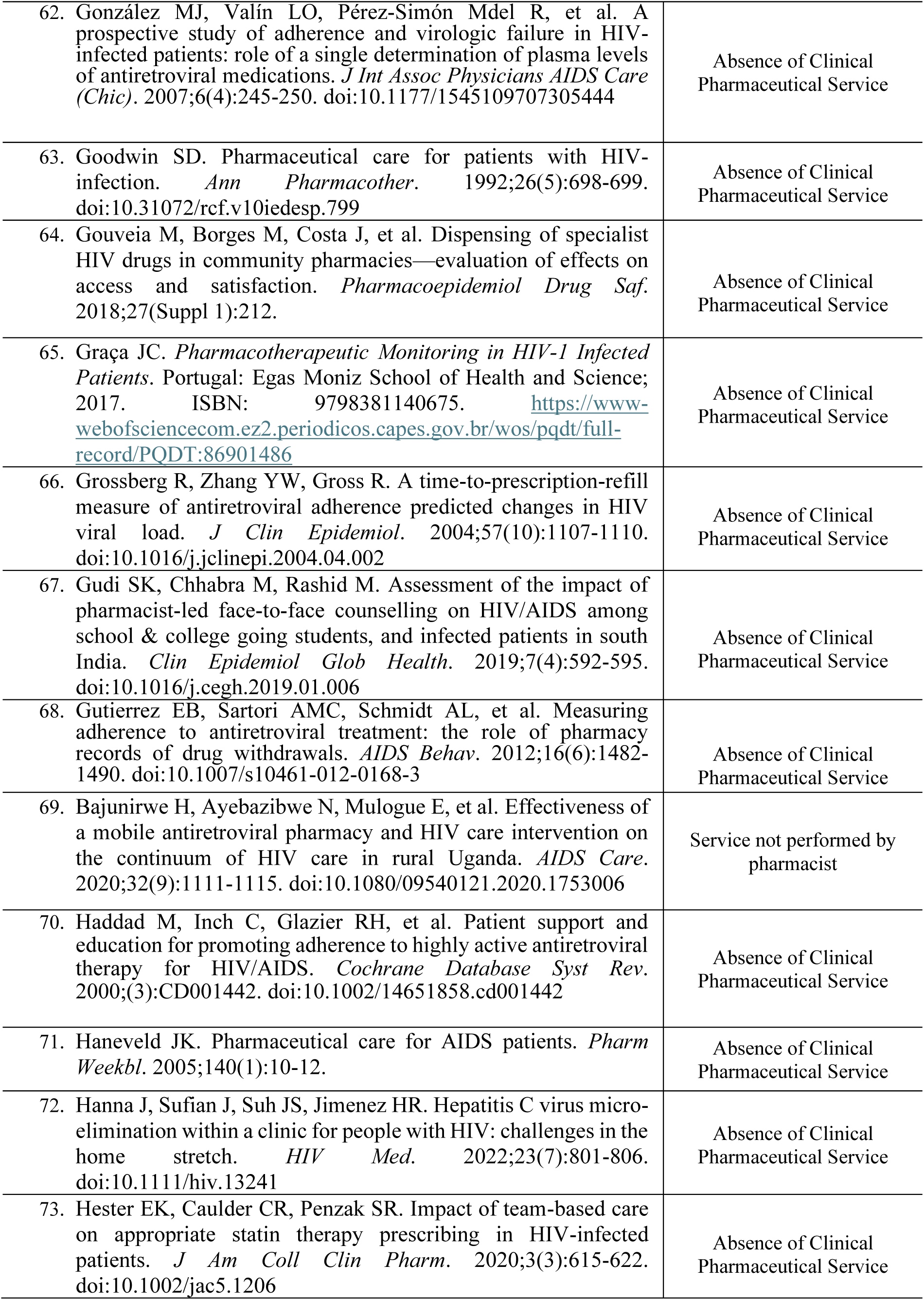
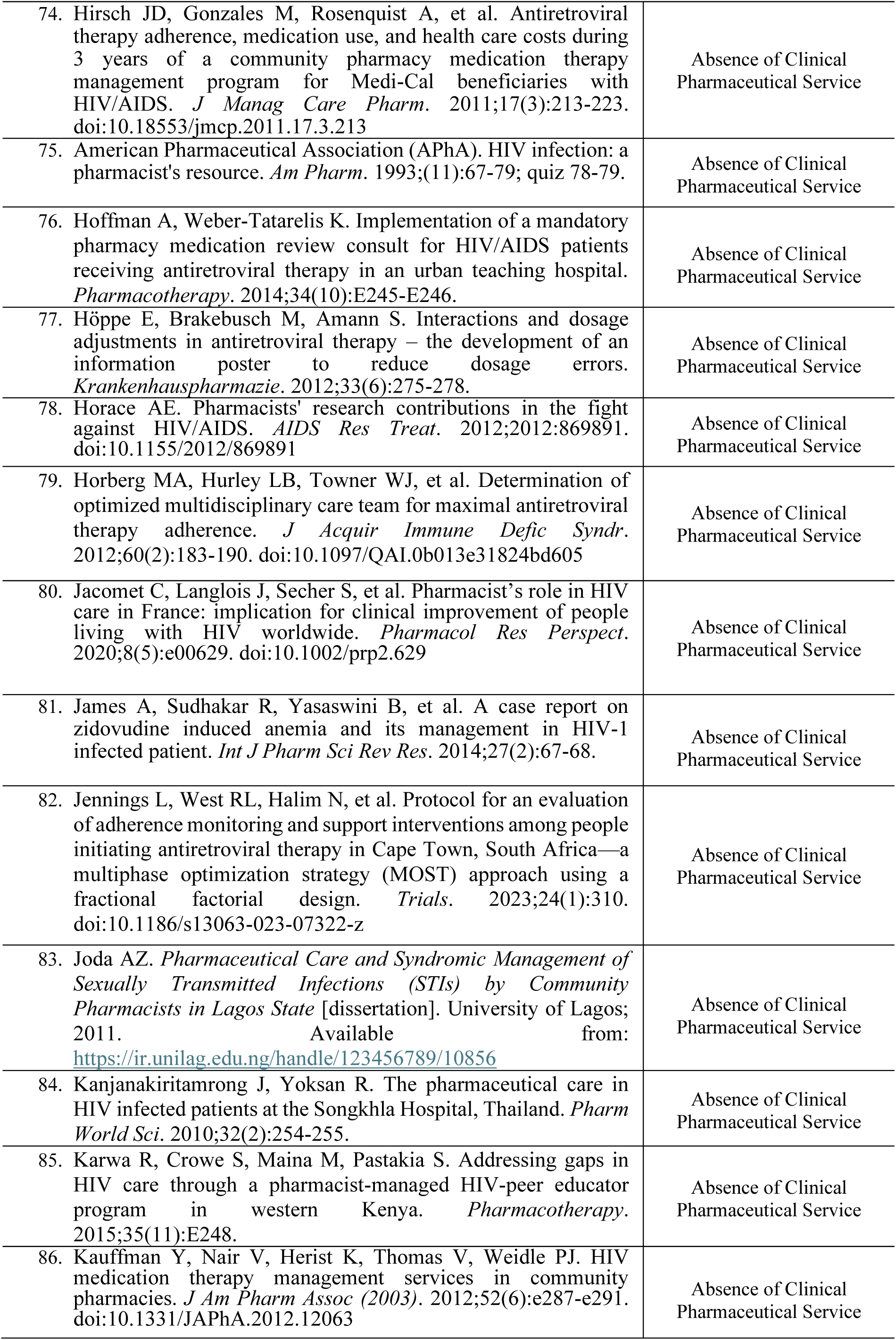
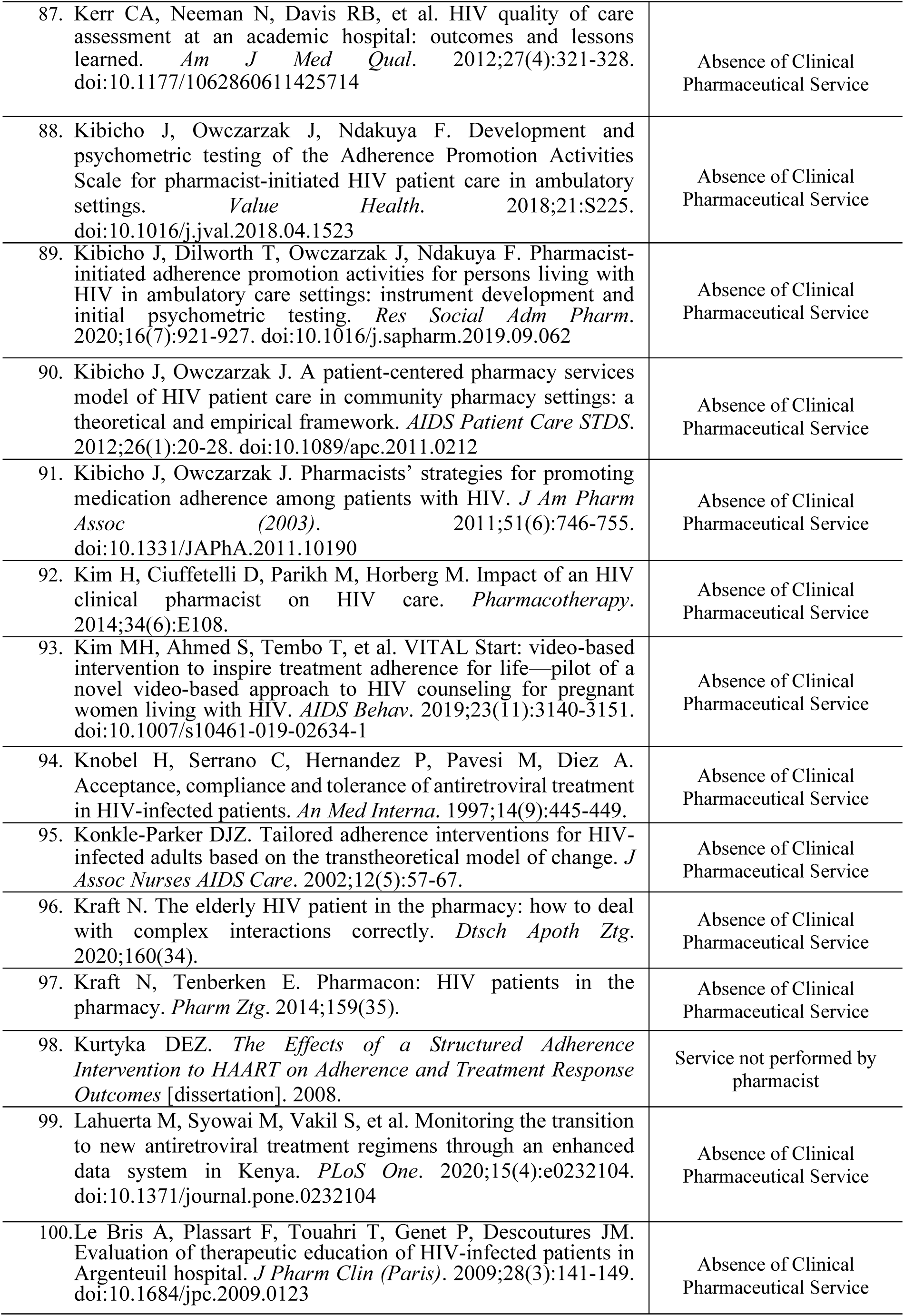
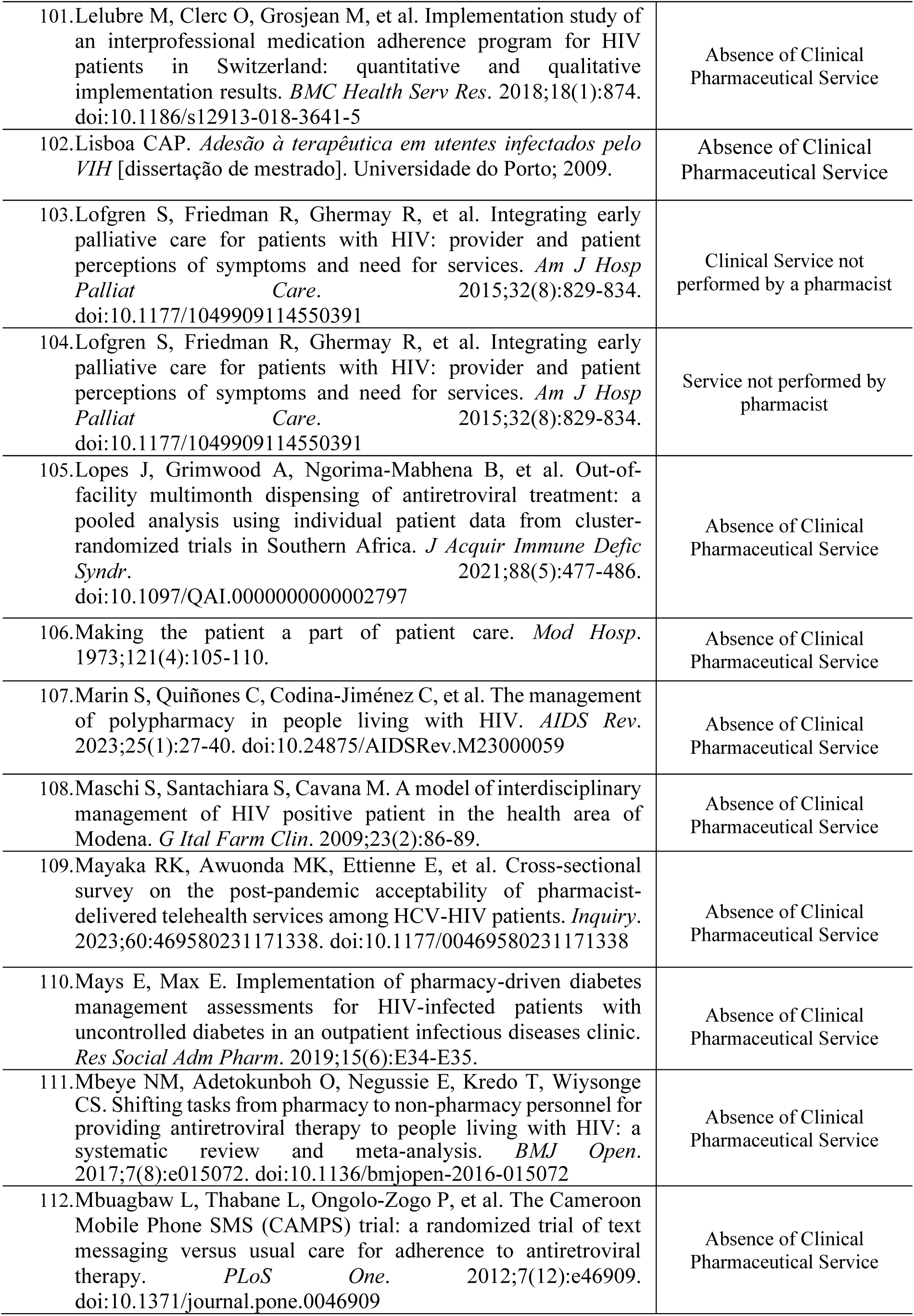
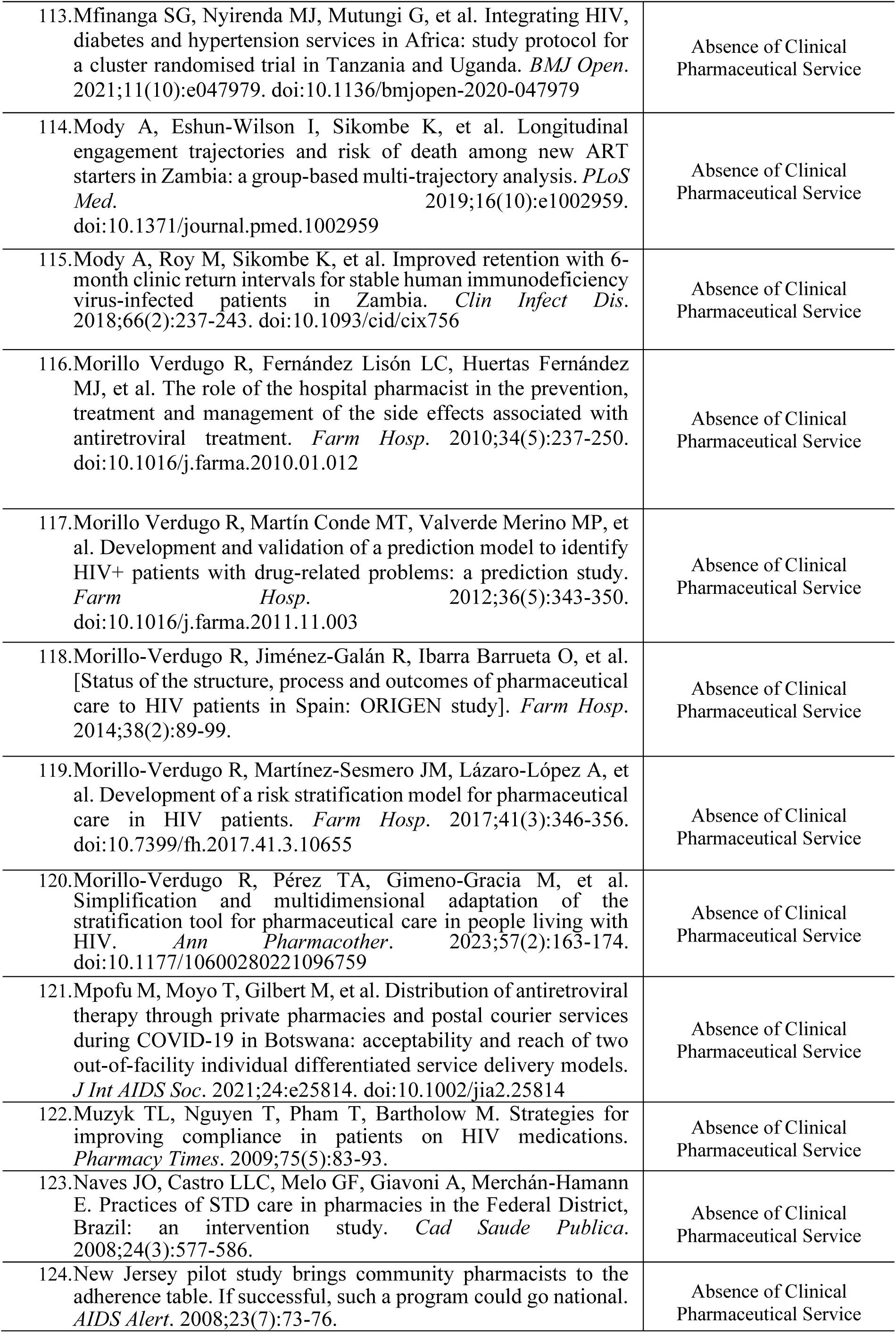
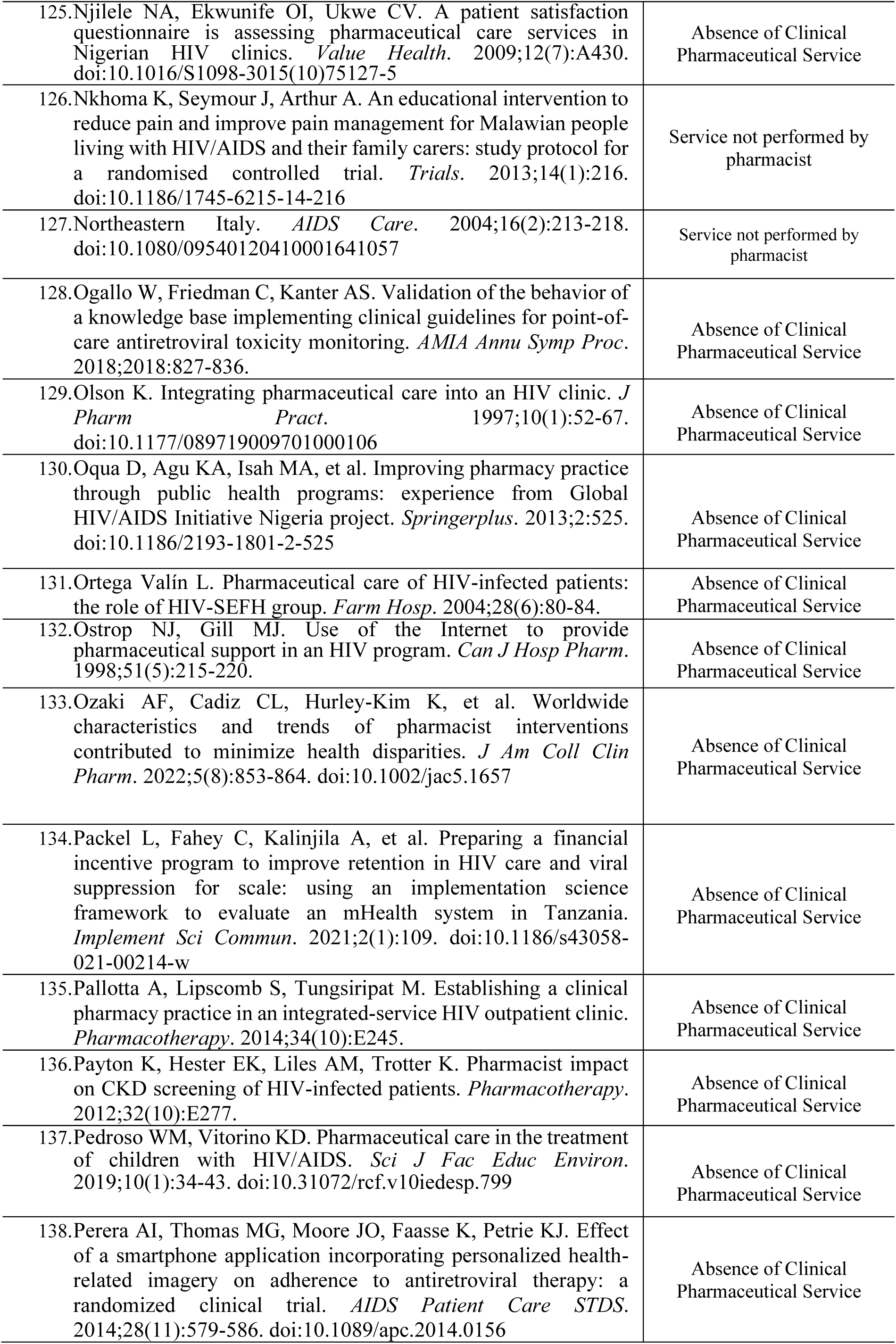
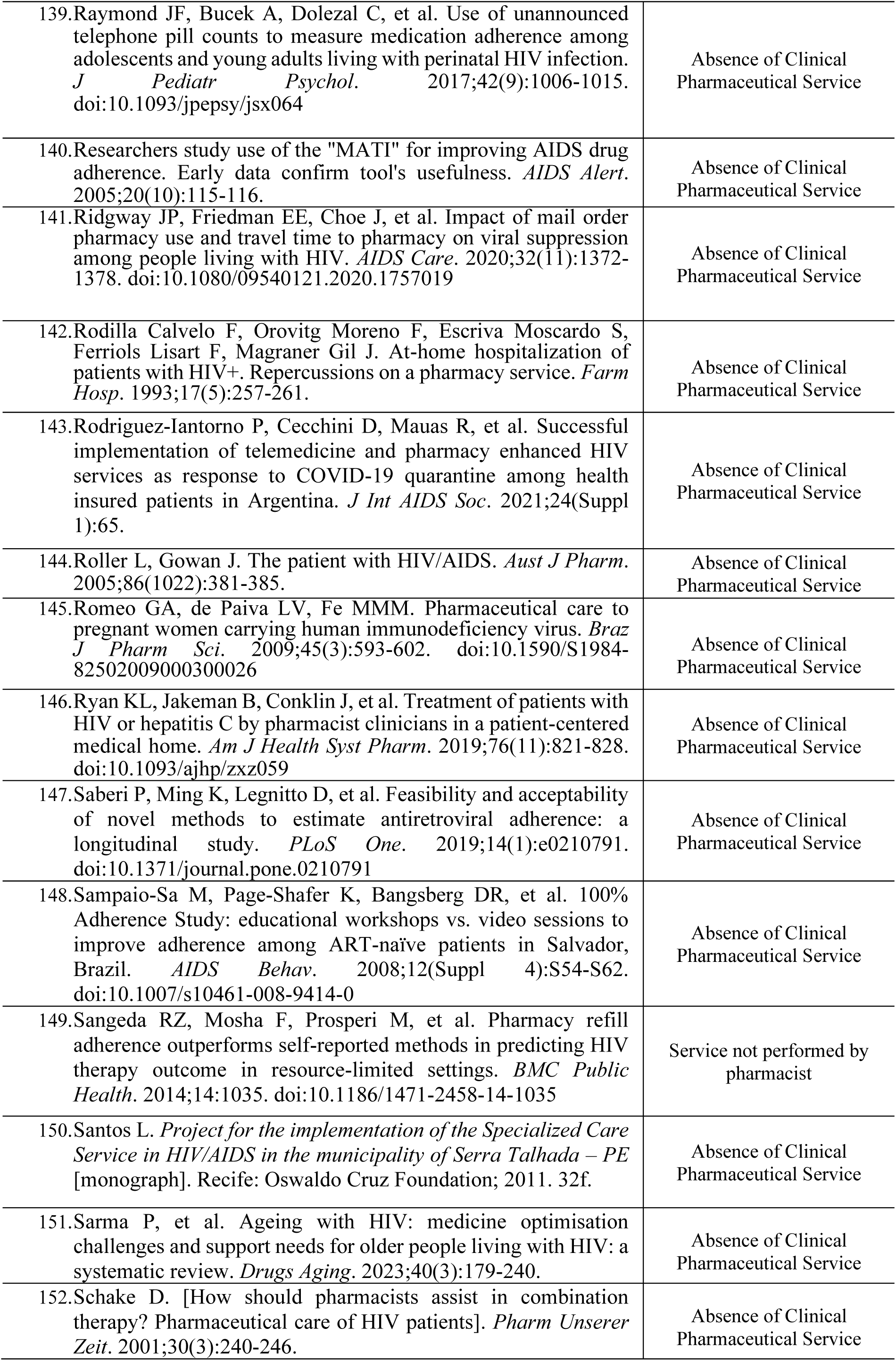
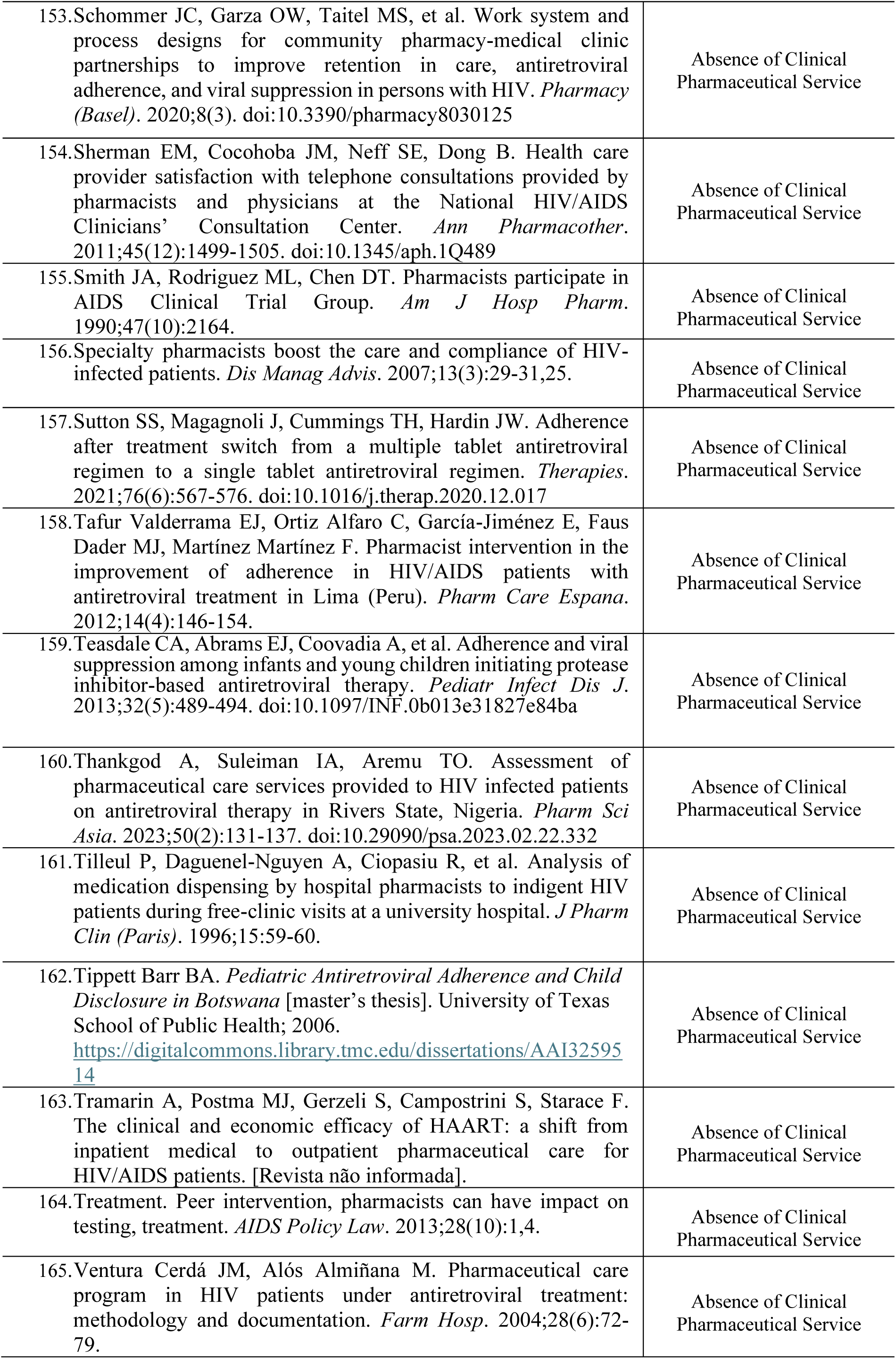
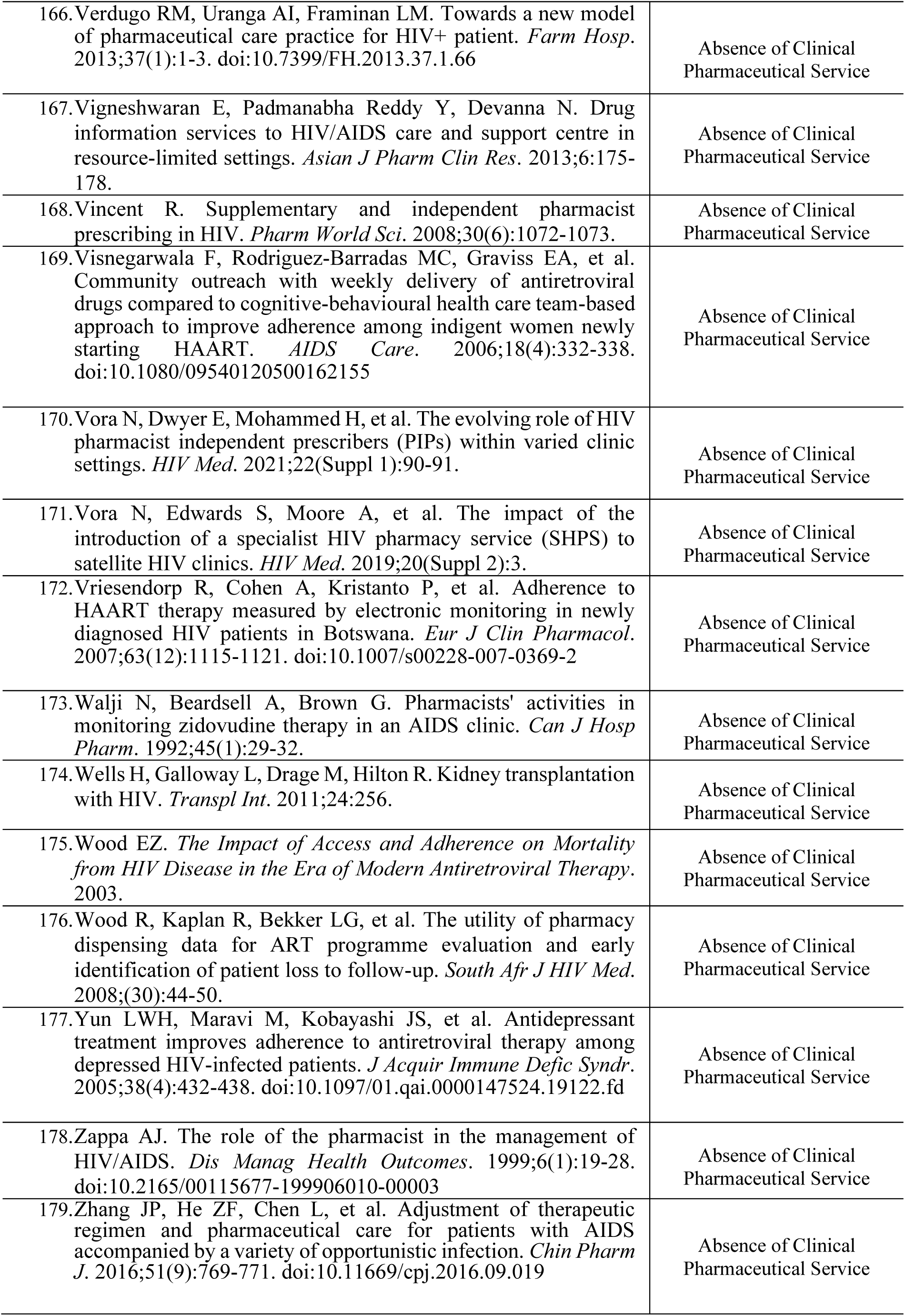
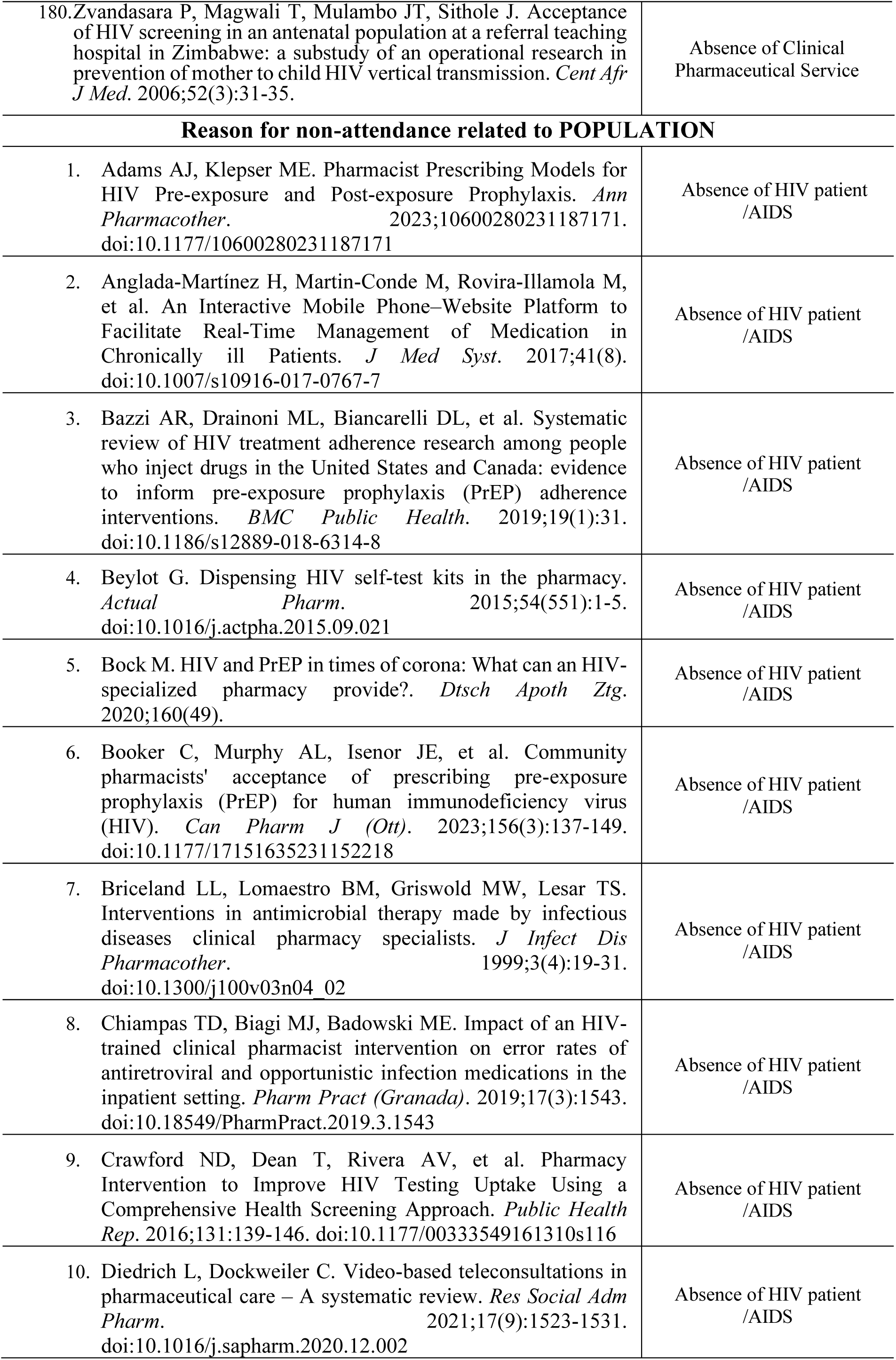
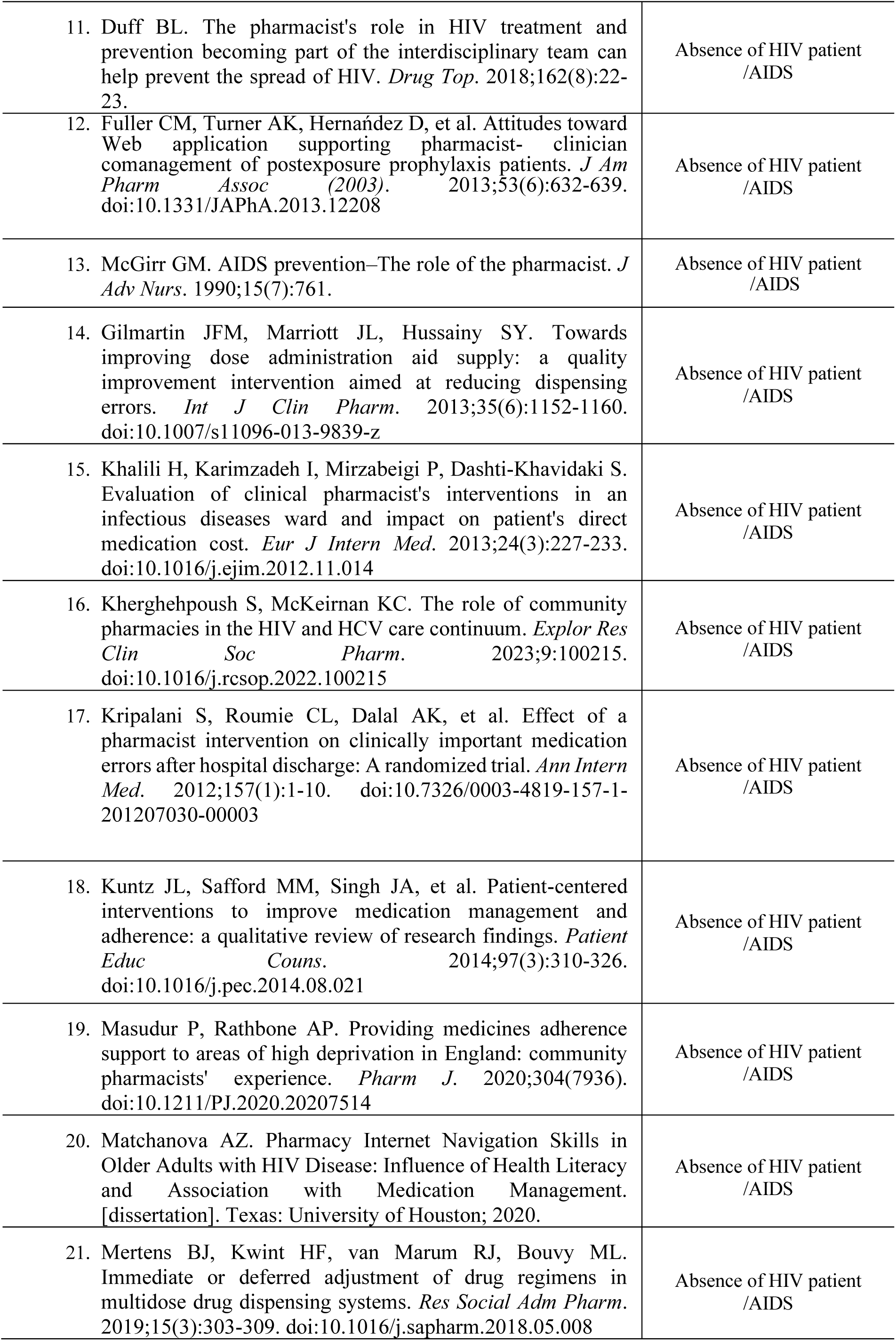
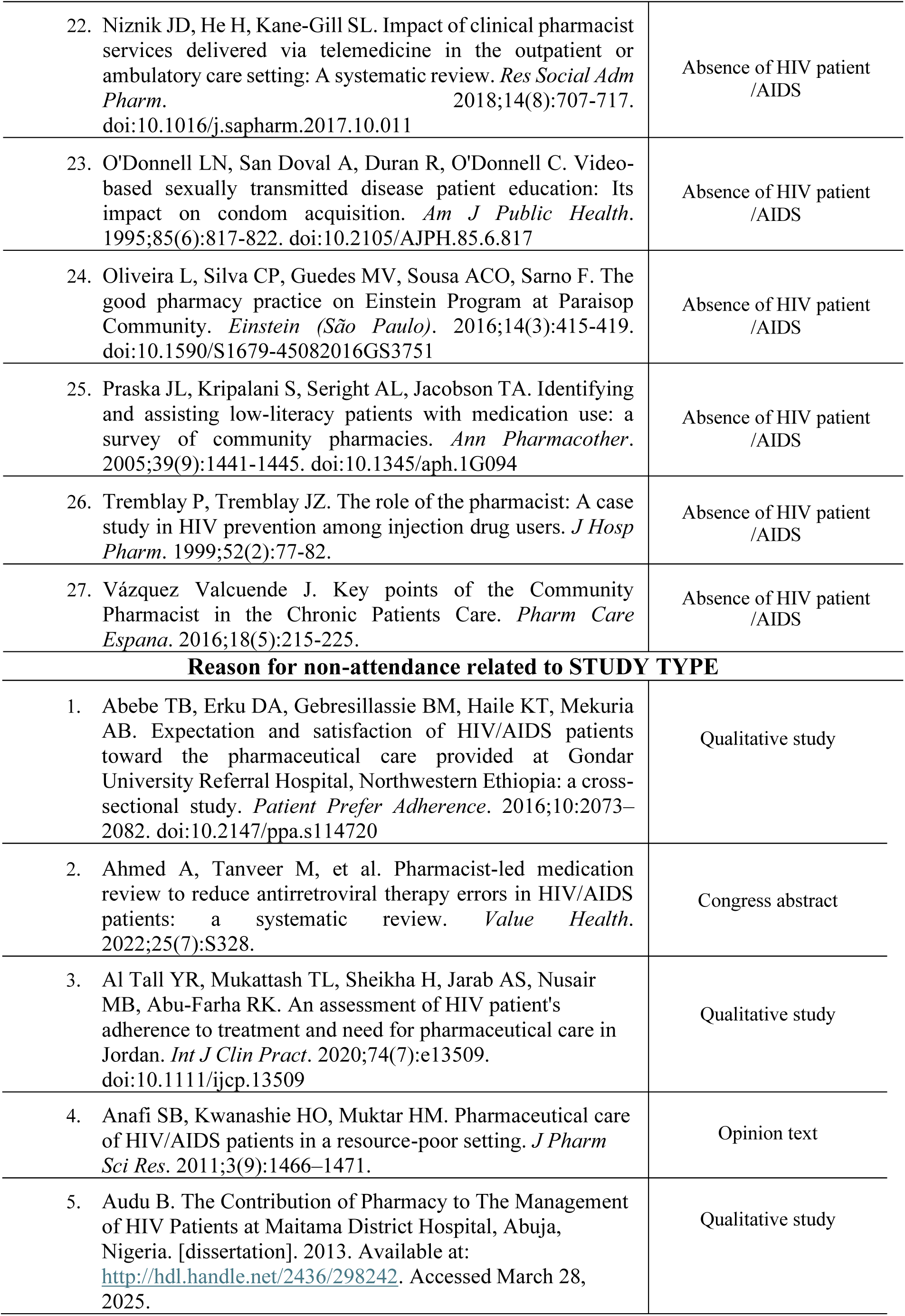
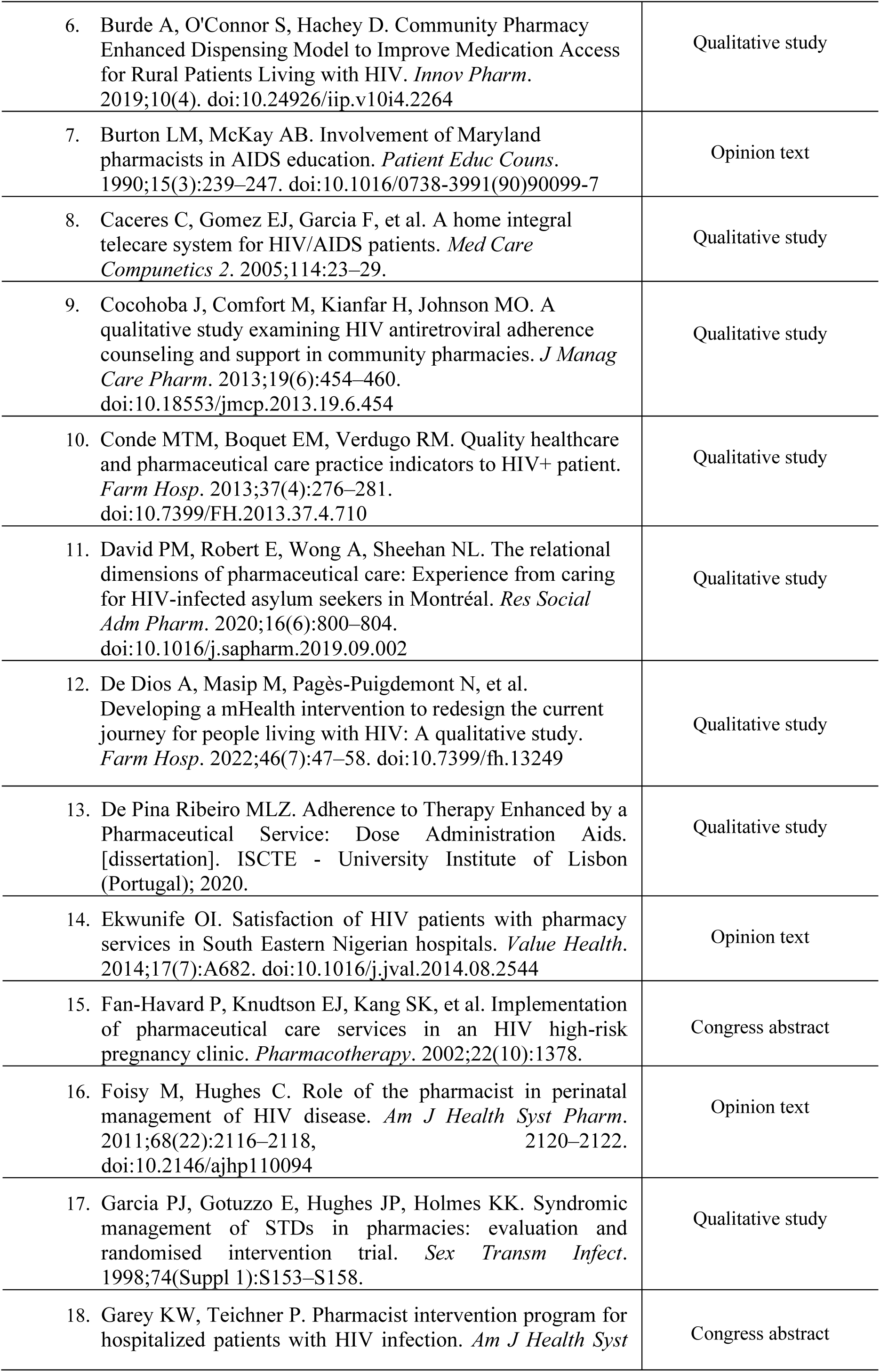
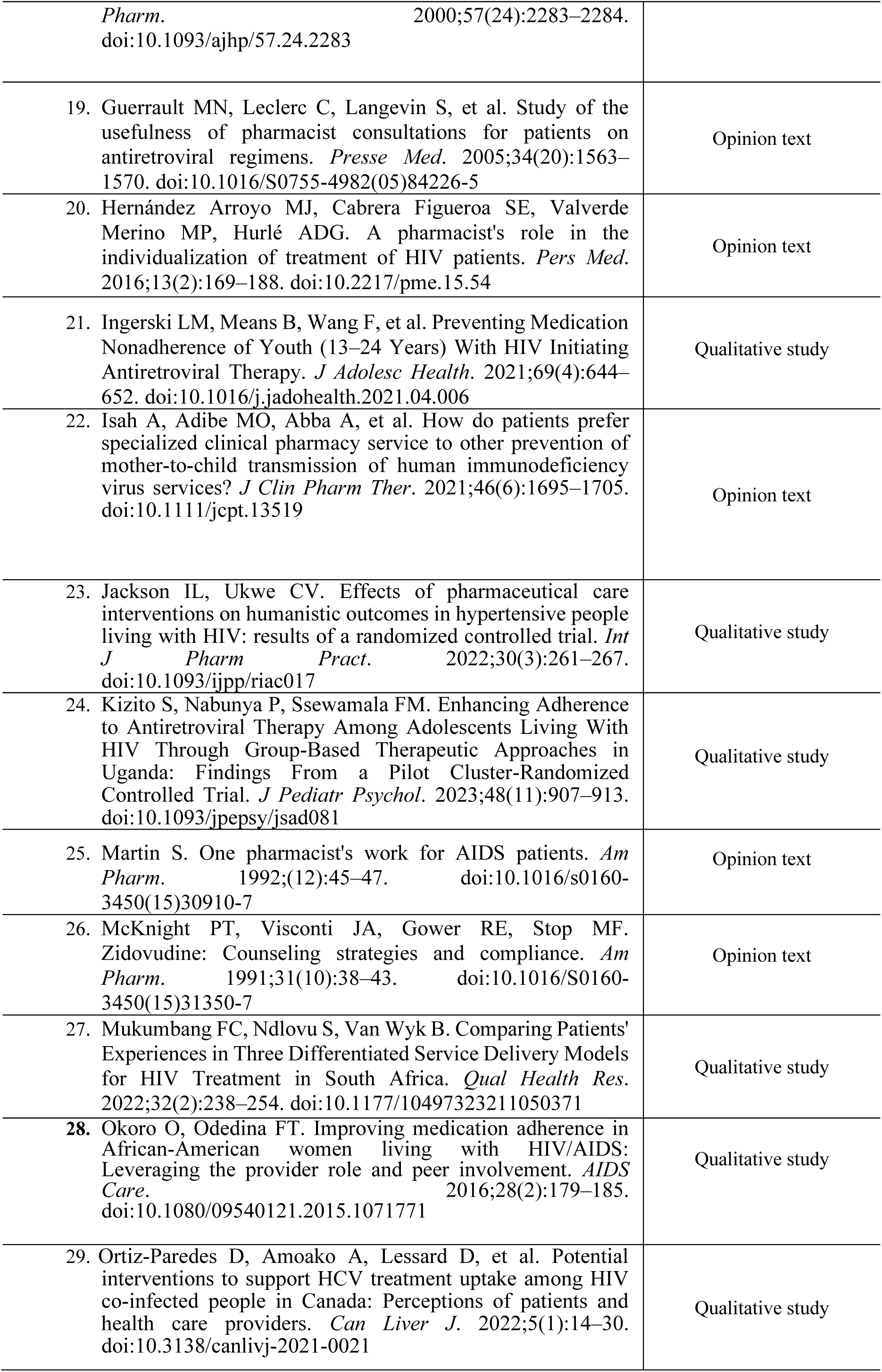
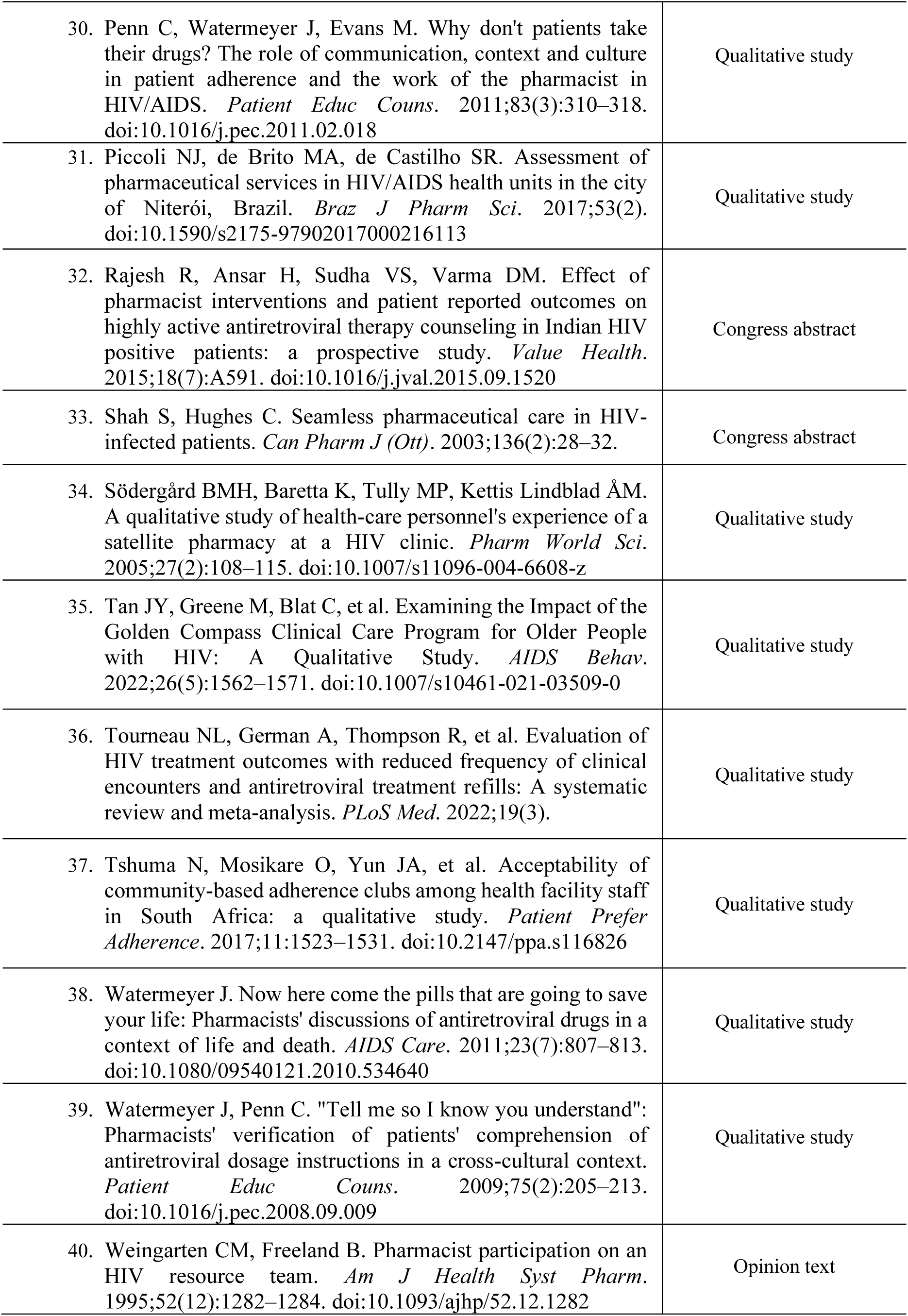
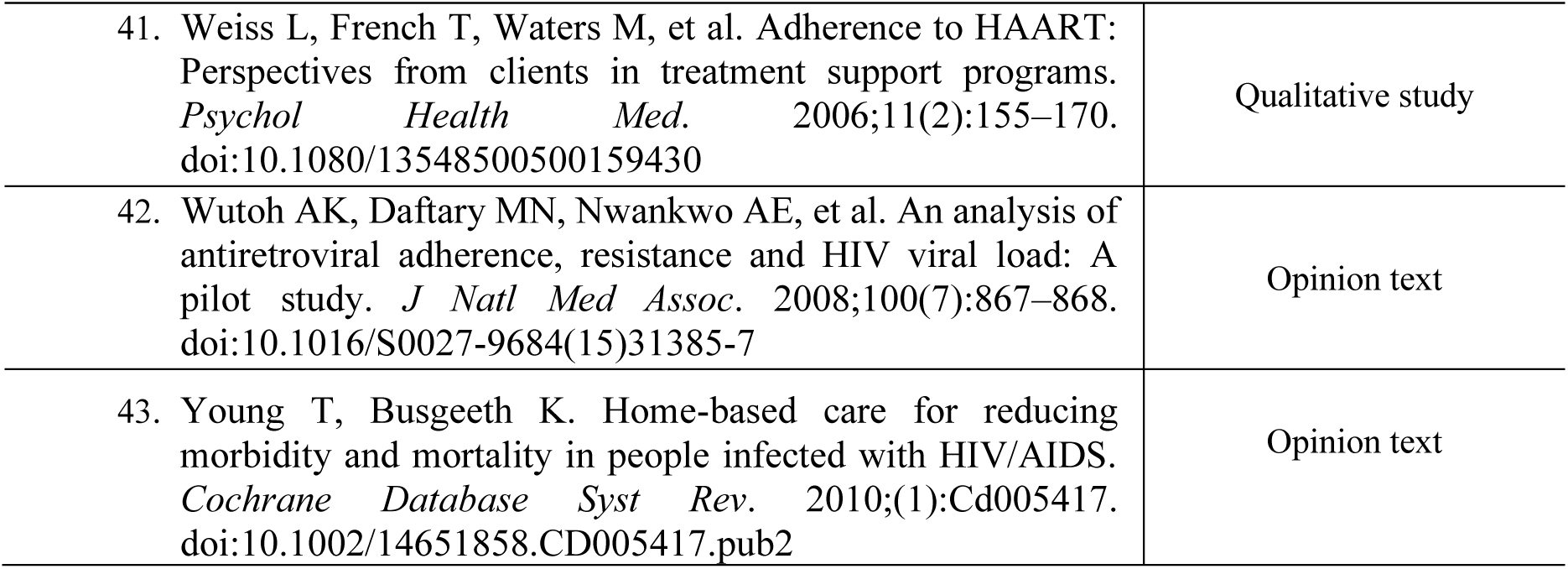

## APPENDIX 4 PRISMA FLOWCHART OF THE SELECTION OF STUDIES

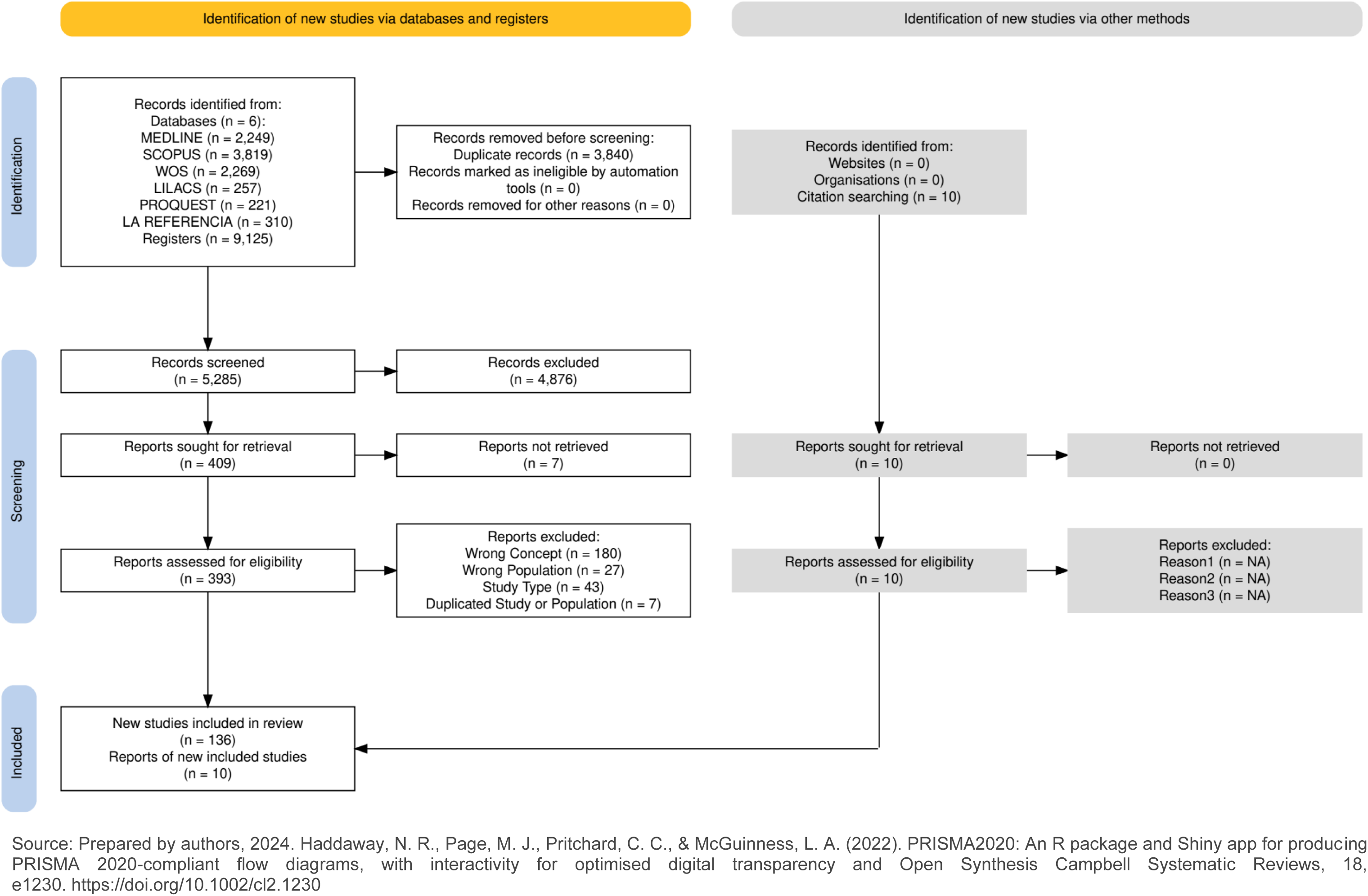

## APPENDIX 5 DATA EXTRACTION GUIDE

APPENDIX 5 – DATA EXTRACTION GUIDE
| Category | Items |
| --- | --- |
| **1- Bibliographic information** | Title:<br>Author<br>Year of publication<br>Country of study |
| **2 - Information regarding the study** | Financing<br>Year of the Environment study<br>Method<br>Study design<br>Sample size and characteristics<br>Duration of the survey<br>Participant description<br>Objective Results<br>Conclusions<br>Study limitations |
| **3 – Clinical pharmaceutical service** | **Type of clinical pharmaceutical service:**<br>Pharmacotherapeutic follow-up<br>Reconciliation of medicines<br>Dispensing of medicines<br>Health education<br>Health condition management<br>Self-limited health problem management<br>Pharmacotherapeutic monitoring<br>Health screening<br>Pharmacotherapy review<br>**Service follow-up details**<br>**Who performs the intervention**<br>**Team composition**<br>**Results**<br>CD4<br>HIV viral load<br>Quality of life<br>Adherence |

## APPENDIX 6 PRIMARY STUDIES, ACCORDING TO PLACE OF CONDUCT AND STUDY DESIGN

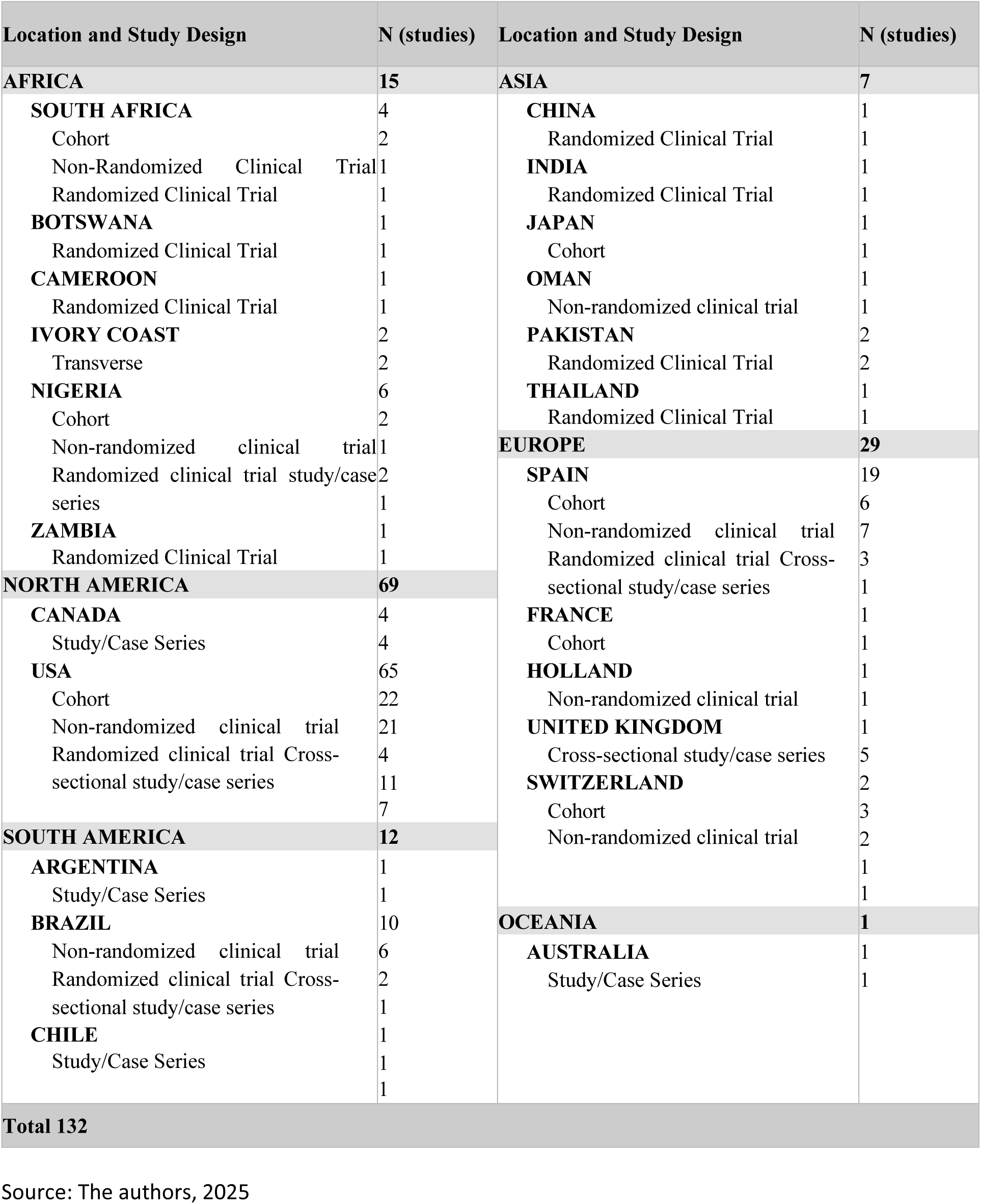

